# Usefulness of ecological mobility and socio-economic indicators in SARS-CoV-2 infection modelling: a French case study

**DOI:** 10.1101/2024.05.05.24306895

**Authors:** Nicolas Romain-Scelle, Benjamin Riche, Thomas Benet, Muriel Rabilloud

## Abstract

**Introduction:** Following its emergence in January 2020, SARS-CoV-2 diffusion occurred for a year with only non-pharmaceutical interventions (NPIs) available as mitigation tools. We aimed to assess the predictive capability of census-based indicators on the infection risk by SARS-CoV-2 in the French Auvergne-Rhône-Alpes region to assist NPIs allocation at the neighbourhood level.

**Methods:** We aggregated all counts of biologically confirmed cases of SARS-CoV-2 infection at the neighbourhood level between May 2020 and February 2021. 10 census-based ecological covariates were evaluated as predictors of case incidence using a Poisson regression with conditional autoregressive (CAR) spatial effects. Benefits of CAR effects and covariates on model fit were evaluated using pseudo-R² and Moran’s I statistics.

**Results:** 438,992 infection cases over 5,410 neighbourhoods among 7,917,997 inhabitants were analysed. The association between covariates and case incidence was inconstant across time and space. Spatial correlation was estimated at high levels. Spatial CAR effects were necessary to improve on the pseudo-R^2^ and the Moran’s I statistics compared to the null model (intercept only).

**Conclusion:** The ecological covariates assessed were insufficient to adequately model the distribution of cases at the neighbourhood level. Excess incidence was found mainly in metropolitan areas before the epidemic wave peak.

## Introduction

The emergence of the SARS-CoV-2 virus and the COVID-19 disease in early 2020 has stressed the necessity of readiness in pandemic response, as no pharmaceutical measure or pre-existent immunity against the pathogen can be expected in the onset of an epidemic. Among all immediately available public health resources available to manage an emergent infectious disease outbreak, contact tracing have proven effective in mitigating reproduction rates, with varying successes depending on exhaustiveness, reactivity and resource allocation [1]. However, contact tracing is a labour-intensive activity, with limited scalability in short delays, as individuals cannot be quickly tasked from one job to another if prior training is required. As such, there is a necessity for policy makers to plan an early control in the case of emergent disease with pandemic potential, as contact tracing can be easily overwhelmed as the number of cases increase [2].

The first wave of the pandemic in France occurred between the first imported case in January 24^th^ 2020 and a near complete recess of viral circulation with a minimum infection incidence rate during the first week of June at 3,169 confirmed cases per 100,000 person-week. This inaugural wave impacted French regions varyingly: the north-east, and the Paris region of France were particularly marred with a high number of cases (up to 74 new hospital admissions per 100,000 person-week) compared to a more moderate impact in the north, south-east and centre regions, and a low impact on the Atlantic coast and associated in-land areas (10/100,000 person-week in the Brittany region).

Using the weekly infection cases incidence rate, a second wave period can be determined around the peak, which occurred the last week of October 2020 (335,204 cases in France). The incidence rate started to increase by the beginning of June 2020, and decreased back to a moderately high level of 72,758 cases in the first week of December 2020 before a slow increase to 128,276 weekly cases by the first week of March 2021. This second wave was also marked by unequal regional impacts, with the south-eastern part of France experiencing the highest incidence rates in infection cases and hospital admissions (49 hospital admissions/100,000 person-week in the Auvergne-Rhône-Alpes region). All epidemiologic indicators are provided by the French Public Health Agency (Santé publique France) and available online [3]

Prognostic factors of COVID-19 disease, morbidity and mortality are an extensively studied topic [4]. Supplementary improvements could be obtained from a better understanding of the risk factors associated with the infection itself, as this understanding would allow for preventive public health actions aimed toward at-risk individuals and populations, such as more intense contact-tracing effort and dedicated procurement of masks.

It is established that infectious diseases do not affect all equally. Beyond the potential existence of genetic risk factors, socio-economic determinants are known predictors of incidence of infection. In the viral world of infectious disease, influenza infections are associated with lower literacy levels, unemployment rates, and home ownership as shown by retrospective works conducted in US cities in 1918-1919 [5–8].

Several socio-economic factors are already proven or highly suspected of being associated with the risk of infection by SARS-CoV-2. A literature review from Khanijahani et al. (2021)[9] has shown that a low education level, poverty or deprivation, household overcrowding and low income are associated with an increase of SARS-CoV-2 infection incidence rate, however with some diverging results. Studies done specifically in France at the individual level has shown that the presence of a child attending school is a risk factor [10]. The same study found the education level as a risk factor, but with a higher risk of infection among individuals with both low and high education level.

As such, the present study aims at identifying relevant socio-economic predictors of an excess risk of infection by SARS-CoV-2 at a population level in order to optimise the use of public health interventions, guiding them toward more at-risk populations, therefore more effectively mitigate emergent infectious diseases.

## Materials and methods

We conducted a retrospective, ecological analysis of the count of incident cases of infection by SARS-CoV-2 within the Auvergne-Rhône-Alpes French region, to assess the association between infection risk and socio-economic indicators from census data. The study region accounted for 12% of the total metropolitan French population, with 8.1 million inhabitants in 2022. Our study was conducted for a time period between May 13^th^, 2020 and February 14^th^, 2021, covering most of the second wave. Aggregated counts of confirmed cases of infection were used as an outcome.

### National information system for infection cases

Infection cases data was extracted from the dedicated national infection case information system (SI-DEP). This system was deployed in France from May 13^th^ 2020 in order to centralize all tests and results of biological analysis searching for SARS-CoV-2 infection, independently of technique used. Upload of tests results is mandatory for all the French medical analysis laboratories, making the system exhaustive. For each test carried out, the system collects the test result and the following information regarding the patient: age, sex, main postal address. The data are anonymized and aggregated before the extraction for analysis, providing a count of people tested and infected by spatial unit. Due to both technical limitations at the time of extraction and a changing definition of the count of individual tested over the study period, only the counts of confirmed cases were available for this study.

### Case definition

A case was defined by the confirmed infection of an individual by the SARS-CoV-2 virus by specific RT-PCR test or antigenic test. In case of repeatedly positive biological analyses, an individual was considered as a new case if the repeated test was positive more than 60 days after the previous one to exclude test positivity associated to viral material persistence at the sampling site.

### Spatial unit

The IRIS (*Ilôt de Regroupement pour l’Information Statistique*), an infra-municipal spatial unit, was used for this ecological study as the aggregation scale for the case count of SARS-CoV-2 infection and the covariates. Each mainland French town *(“commune”*) counting 15,000 residents or more is split between multiples IRIS, with towns between 5,000 and 15,000 inhabitants being split if necessary. The composition target for each IRIS is a resident population of 1,500 and a coherence of habitation type.

### Temporal unit

This study was conducted over four time periods, between May 13^th^ 2020 and February 14^th^ 2021, defined empirically to capture four phases of the second wave corresponding respectively to extremely low case count following the first wave and lockdown, rapid case count growth, incidence peak and decrease, stabilization at an elevated level of weekly incidence. Data aggregation over time was necessary to limit the number of spatial units with case counts censored to prevent patients’ identification. The four periods were defined as follow:

- May 13^th^ to July 26^th^ 2020 (weeks 20 to 30), period P1, said “Low incidence”,
- July 27^th^ to October 25^th^ 2020 (weeks 31 to 43), period P2, said “Growth”,
- October 26^th^ to December 13^th^ 2020 (weeks 44 to 50), period P3, said “Peak and decrease”,
- December 14^th^ 2020 to February 14^th^ 2021 (weeks 2020-51 to 2021-06), period P4, said “Stabilization”

### Inclusion and exclusion criteria

All 5,410 IRIS belonging to the Auvergne-Rhône-Alpes region of mainland France were included. Fifteen IRIS for which the number of inhabitants was 0 or near 0 were excluded of the analysis (industrial/commercial areas with marginal habitations). All IRIS for which census-based indicators were unavailable were excluded from the analysis (341 units).

### Socio-economic, mobility and population density indicators

The socio-economic indicators potentially associated with the distribution of infection cases were the proportion of migrants, the proportion of unemployed individuals, the proportion of single person homes, the proportion of households without a child, the proportion of car ownership, the proportion of individuals without a high school diploma, and the proportion of overpopulated homes (French National Institute of Statistics and Economic Studies, INSEE). Two other indicators were included to account for population mobility in volume and nature, i.e. the proportion of individuals working outside of their residency town, and the proportion of individuals travelling to work by car. Their distributions were described in Table 1, defined and represented using maps in Supplementary Material S1. To control for the urbanisation degree of the spatial units, a combined indicator in four levels measuring population density and urbanization, was included in all the models. A detailed definition of this indicator is provided in Table 1. This indicator was grouped into two levels for the purpose of the present analysis (Very high-High vs. Low-Very low). All mentioned indicators were obtained from the 2017 population census data. The socio-economic indicators were available at the IRIS level. The mobility and population density were available at the town level. All indicators are censored at the 1^st^ and 99^th^ centiles to control for extreme data issuing from low population areas.

**Table 1:** Description of the demographic and socio-economic characteristics of the IRIS (inframunicipal spatial units) of the French region Auvergne-Rhône-Alpes.

| Characteristics | N<sup>a</sup> = 5,069 |
| --- | --- |
| Number of individuals at risk, median (IQR) | 1,149 (444, 2,403) |
| Population density index<sup>b</sup> (4 levels) n (%) |  |
| Very low | 863 (17) |
| Low | 2,249 (44) |
| High | 1,053 (21) |
| Very high | 904 (18) |
| % of immigrants, median (IQR) | 4 (2, 9) |
| % of individuals unemployed, median (IQR) | 9.8 (7.3, 13.6) |
| % of single person homes, median (IQR) | 31 (25, 39) |
| % of individuals working outside their residency town, median (IQR) | 59 (37, 72) |
| % of individuals travelling to work by car, median (IQR) | 83 (73, 89) |
| % of families without a child, median (IQR) | 50 (45, 57) |
| % of car ownership, median (IQR) | 93 (87, 96) |
| % of low education, median (IQR) | 55 (48, 62) |
| % of overpopulated homes, median (IQR) | 1.60 (0.00, 3.48) |

<sup>a</sup> number of spatial units with available data
<sup>b</sup> The density index is defined with respect to the proportion of the population of a town living
in an urban centre (50,000 people living within a contiguous area with a population density above
1,500 ind/km<sup>2</sup> or more), an urban cluster (5,000 people, 300 ind/km<sup>2</sup> or more), or an intermediary
rural aggregate (300 people, 25 ind/km<sup>2</sup> or more): 50% or more of the population is within a urban
centre: Very high ; 50% or more of the population lives within a urban centre or a urban cluster: High ;
50% or more of the population lives outside a centre, cluster or aggregate: Very low ; Otherwise: Low

### Statistical analysis

To quantify the association between the socio-economic indicators and case incidence, four models described in Table 2 were carried out for each period. The first model (M1), serving as reference, was a generalized linear model with the counts of cases following a Poisson distribution, estimating the mean incidence rate across the region (intercept only). A second GLM model was carried out (M2), with the introduction of the studied covariates. A conditional autoregressive (CAR) random effect was added in the third (M3) and fourth models (M4), to account for the spatial autocorrelation of the case counts. The CAR random effect proposed by Leroux and MacNab [11,12] was used based on its capacity to provide estimates of all parameters with minimum bias in scenarios of both high and low spatial correlation compared to other CAR effects [13].

**Table 2:** Statistical models used to analyse the count of COVID-19 cases per IRIS (infra-communal spatial units) of the French region Auvergne-Rhône-Alpes during the second epidemic wave of COVID-19, with their respective labels and detailed composition

| Label | Model<sup>a</sup> | Description |
| --- | --- | --- |
| M1: Null | $Y_k \sim \text{Poisson}(\lambda_k) ; \ln(\lambda) = \mu$ | Intercept only: $\mu$ |
| M2: Covariates only | $Y_k \sim \text{Poisson}(\lambda_k) ; \ln(\lambda) = \mu + X\beta$ | X: vector of covariates, $\beta$: vector of regression parameters |
| M3: Spatial effect only | $Y_k \sim \text{Poisson}(\lambda_k) ; \ln(\lambda) = \mu + \phi$ | Intercept and spatial effect ($\phi$) |
| M4: Full | $Y_k \sim \text{Poisson}(\lambda_k) ; \ln(\lambda) = \mu + X\beta + \phi$ | Covariates and spatial effect |

<sup>a</sup> $Y_k$ : count of cases in IRIS k

The CAR random effect follows a multinomial distribution, with two parameters to estimate, i.e. a spatial correlation coefficient p measuring the strength of the dependence among neighbouring spatial units, and a parameter r^2^ weighting the variance of the spatial effect component around its expectancy.

Parameters’ estimate was obtained using a Bayesian approach, with weakly informative priors. All *β* parameters related to a continuous covariate and corresponding rate ratios are given for one standard deviation increase of the covariate. Incidence rate ratios (IRR), defined as the ratio between the predicted incidence rate within one spatial unit and the mean predicted incidence rate across the study region, were presented using maps. Values above 1 therefore indicate an excess incidence.

The neighbourhood structure between spatial units was defined on a travel-time-based metric. All IRIS within 60 minutes of each other were considered neighbours, with a weight defined as the inverse of the computed travel time separating each other. Normalization of the weight matrix imply a mean variance for the random effect of 1 when *τ*^2^ is equal to 1.

Each model was evaluated by computing the Moran index of the residuals to assess whether the model was successful in accounting for the spatial autocorrelation. To assess goodness-of-fit, the observed case count in each IRIS was compared to the posterior distribution of the predicted count. A good fit was defined by an observed count in the interval defined by the 5^th^ and 95^th^ centiles of the posterior distribution. We used the pseudo-R² proposed by McFadden [14] to measure and compare, for each period, the variance explained by the models.

A sensitivity analysis was conducted to assess the validity of modelling over the entire study region. We fitted the full model in each of the 12 *Départements* (French administrative subdivision) composing the study region.

Detailed models’ specification, address of the violation of the case independence hypothesis, weight matrix computation, and estimation procedure are reported in Supplementary Material 2. All analysis were conducted using R software (4.1.2). Travel times were computed using the r5r package [15] to interface with the Conveyal R5 routing engine [16]. Geographic data was extracted from the OpenStreetMap dataset. The model parameters were estimated using the package CARBayes [17].

## Results

The study region is composed of 5,410 IRIS, with 5,069 for which all variables were available, and 7,917,997 individuals at risk (Table 1). The total number of cases observed during the study period was 562,376, of which 438,992 cases could be attributed to one IRIS and used in this study (78%).

Unless stated otherwise, all results regarding parameters estimates are extracted from the full model (M4). Across all periods, all socio-economic indicators were significantly associated with the rate of confirmed cases of SARS-CoV-2 infection (Table 3). Those associations were not statistically significant for all periods, nor were they homogeneous. The effect of the proportion of low education level was dependent of the observation period: during the Growth period, it was associated with an incidence rate reduction of 10% for an increase of 11.6% of the proportion of low education level (rate ratio (RR): 0.90, 95% credibility interval (CI): [0.88;0.93]), but during the Peak and decrease, and Stabilization periods the estimated RR was above 1 (respectively 1.08, 95% CI: [1.05;1.10] and 1.06, 95% CI:[1.04;1.09]). The estimated RR for unemployment, statistically significant only during the Peak and decrease period, was under 1, corresponding to a decrease of 4% of the incidence rate for an increase of 5.7% of the proportion of unemployed individuals (RR: 0.96, 95% CI: [0.93;0.99]). The proportion of migrants was the only indicator found to be associated with the risk of infection during the Low incidence period, corresponding to an increase of 15% of the incidence rate for an increase of 7.2% of the proportion of migrants (RR: 1.15, 95% CI: [1.04;1.29]). The increase of the proportion of individuals living alone and of car ownership were both associated with a significant incidence rate reduction during the Growth period and a significant increase during the Peak and decrease period.

**Table 3:** Effect of the demographic and socio-economic characteristics of the IRIS (infra-municipal spatial units) of the French region Auvergne-Rhône-Alpes on the rate of SARS-CoV-2 infection (rate ratio estimates) during the four periods of the second epidemic wave: results of Poisson models with inclusion of covariates and a conditional autoregressive (CAR) random effect (models M4)

| Variable | Period |  |  |  |
| --- | --- | --- | --- | --- |
|  | Low incidence | Growth | Peak and decrease | Stabilization |
| Mean predicted weekly incidence<sup>a</sup> | 0.01<br>[0.00;0.01] | 0.68<br>[0.66;0.70] | 2.47<br>[2.41;2.54] | 1.28<br>[1.24;1.31] |
| Very High or High population density index (ref. Low or Very low) | 1.24<br>[0.99;1.56] | **1.29**<br>**[1.20;1.39]** | 1.05<br>[0.98;1.11] | **1.09**<br>**[1.02;1.16]** |
| **Socio-economic indicators** (for 1 standard deviation increase) |  |  |  |  |
| % of low education (SD: 11.6%) | 0.96<br>[0.87;1.05] | **0.90**<br>**[0.88;0.93]** | **1.08**<br>**[1.05;1.10]** | **1.06**<br>**[1.04;1.09]** |
| % of single person homes (SD: 10.6%) | 0.98<br>[0.88;1.09] | **0.93**<br>**[0.90;0.97]** | **1.07**<br>**[1.04;1.10]** | 0.98<br>[0.95;1.01] |
| % of car ownership (SD: 10.4%) | 1.08<br>[0.92;1.27] | **0.89**<br>**[0.85;0.93]** | 1.01<br>[0.97;1.05] | 1.00<br>[0.96;1.04] |
| % of immigrants (SD: 7.2%) | **1.15**<br>**[1.04;1.29]** | **1.06**<br>**[1.02;1.10]** | **1.04**<br>**[1.01;1.08]** | 1.01<br>[0.98;1.05] |
| % of individuals unemployed (SD: 5.7%) | 1.11<br>[0.98;1.25] | 1.01<br>[0.98;1.05] | **0.96**<br>**[0.93;0.99]** | 1.02<br>[0.98;1.05] |
| % of individuals working outside their residency town (SD: 20.6%) | 1.09<br>[0.99;1.19] | **1.04**<br>**[1.02;1.07]** | 1.01<br>[0.99;1.04] | **1.04**<br>**[1.02;1.07]** |
| % of individuals travelling to work by car (SD: 15.9%) | 0.86<br>[0.73;1.01] | 0.99<br>[0.94;1.03] | **1.06**<br>**[1.02;1.10]** | 0.99<br>[0.95;1.02] |
| % of families without a child (SD: 9.9%) | 0.95<br>[0.85;1.05] | **0.94**<br>**[0.91;0.97]** | **0.96**<br>**[0.93;0.99]** | 0.99<br>[0.97;1.02] |
| % of overpopulated homes (SD: 2.8%) | **1.15**<br>**[1.03;1.28]** | **0.96**<br>**[0.94;0.99]** | **0.97**<br>**[0.95;0.99]** | 0.98<br>[0.95;1.01] |
| **Spatial effect parameters** |  |  |  |  |
| Spatial correlation coefficient ( $\rho \in (0,1)$ ) | 0.485<br>[0.300;0.656] | 0.761<br>[0.680;0.832] | 0.888<br>[0.835;0.931] | 0.603<br>[0.529;0.669] |
| Variance weighting parameter ( $\tau^2 \in \mathbb{R}_+$ ) | 1.342<br>[1.150;1.564] | 0.340<br>[0.319;0.363] | 0.256<br>[0.239;0.274] | 0.284<br>[0.268;0.302] |

*\*The point estimate and the 95% credible interval is given for each model parameter and each study period. Apart from the base risk and the spatial effect parameters, estimates significantly different than their neutral values are emboldened.*
<sup>a</sup> Incidence rate of SARS-CoV-2 infection in an IRIS of 1,000 inhabitants, low population density and mean covariates level (/1,000 person-week).

Among mobility-related indicators, the proportion or individuals working outside their residency town was found positively associated with an increased risk of infection, significantly so during the Growth and Stabilization periods (RR: 1.04, 95% CI: [1.02;1.07] in both cases). The proportion of individuals travelling to work by car was positively associated with the incidence rate during the Peak and decrease period (RR: 1.06, 95% CI: [1.02;1.10]), but appeared negatively associated with the incidence rate during the other periods (not significantly).

A very high or high population density index was found to be associated with an increase of the incidence rate in comparison to a low or very low index, although statistical significance was reached only for the Growth and Stabilization periods (respectively RR: 1.29, 95% CI: [1.20;1.39] and RR: 1.09, 95% CI: [1.02;1.16]).

In the full model the spatial correlation coefficients were estimated at 0.49 and 0.60 for the Low incidence and Stabilization periods respectively, and 0.76 and 0.89 for the Growth and Peak and decrease periods, indicating a varying strength of dependence of the incidence rates between neighbours across periods. The variance weighting parameter was low for the Growth, Peak and decrease and Stabilization periods, between 0.26 and 0.34, but was found significantly higher during the Low incidence period (1.34, 95% CI: [1.15;1.56]).

Models’ residuals for the Low incidence and Stabilization periods were not spatially correlated, per their respective Moran indices (maximum: 0.08) (Table 4). For the Growth period, both the inclusion of a spatial effect and the inclusion of covariates were able to mitigate the residual autocorrelation indicating a spatial structure of the covariate distribution. The explained variance for the Growth period was also noticeably improved by the introduction of covariates (M2), with the pseudo-R² reaching 0.15. For the Peak and decrease and Stabilization periods, the proportion of variance explained improved significantly only with the addition of a spatial effect to the model, with no relevant difference between M3 and M4. During the Low incidence period, the pseudo-R² showed marginal improvement between M3 and M4, showing a small benefit from the covariates addition. Model fitting was satisfactory for M3 and M4 only during the last three periods (Supplementary Material S3).

**Table 4:** Moran indices and McFadden pseudo-R², for each model fitted on the complete dataset, by epidemic period.

| Period | Model | McFadden R<sup>2</sup><sup>a</sup> | Moran I<sup>b</sup> |
| --- | --- | --- | --- |
| Low incidence | Null |  | 0,048 |
|  | Covariates only | 0,044 | 0,017 |
|  | Spatial effect only | 0,416 | 0,076 |
|  | Full | 0,441 | 0,027 |
| Growth | Null |  | 0,410 |
|  | Covariates only | 0,150 | 0,139 |
|  | Spatial effect only | 0,688 | 0,088 |
|  | Full | 0,687 | 0,044 |
| Peak and decrease | Null |  | 0,230 |
|  | Covariates only | 0,057 | 0,186 |
|  | Spatial effect only | 0,680 | 0,045 |
|  | Full | 0,680 | 0,037 |
| Stabilization | Null |  | 0,052 |
|  | Covariates only | 0,029 | 0,042 |
|  | Spatial effect only | 0,565 | 0,042 |
|  | Full | 0,567 | 0,026 |

<sup>a</sup> *Pseudo-R<sup>2</sup> are computed using the corresponding null model as reference.*
<sup>b</sup> *All Moran indices are significant (at a threshold of 0.05).*

Cartography of the predicted IRRs by the full model (M4) for each period is provided in Figure 1 (high resolution version with metropolitan areas foci available in Supplementary Material S4). The “Low incidence” period was marked by a sharp distinction between urban centres exhibiting excess incidence and the rest of the region (Figure 1A). The “Growth” period displayed IRRs above 1 within and around urban areas mostly, showing a development of the second wave in a limited number of populated areas (Figure 1B). The maps for the “Peak and decrease” and “Stabilization” periods expressed a progressive diffusion of the elevated IRRs across the study region, with the last period showing a dispersed IRR spatial distribution (Figures 1C and 1D). A longitudinal gradient was observed in the first three periods, with IRRs under 1 preferentially in the western part of the study region and IRRs over 1 in the eastern part.

**Figure 1:**
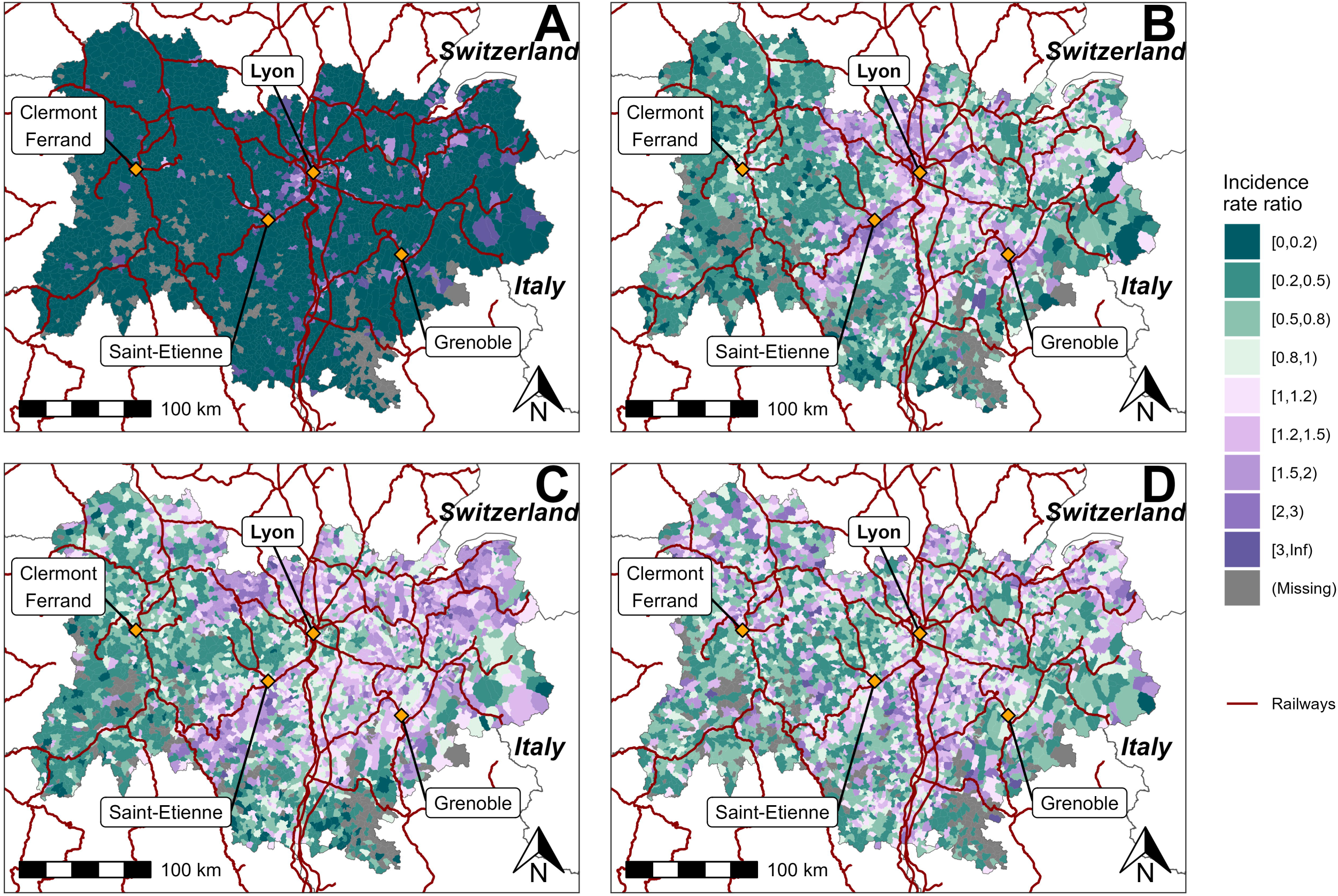
predicted incidence rate ratio of SARS-CoV-2 infection for the IRIS (infra-municipal spatial unit) of the French region Auvergne-Rhône-Alpes during the second epidemic wave: model with covariates and spatial effect (M4). From A to D: Low incidence, Growth, Peak and decrease, and Stabilization periods.

The sensitivity analysis revealed varying direction of association between the covariates and infection incidence across spatial subunits (Supplementary Material S5), and lower than average estimates of the spatial correlation in the Alps (northeastern part of the study region).

## Discussion

This research found several associations between ecological factors and infection risk. Population concentration was found to be the most preeminent ecological characteristic to explain the case distribution, with a higher population density associated with an increased risk of infection, even more so during the build-up to the second wave peak in November 2020.

The results found regarding the socio-economic and mobility indicators are less clear, with only three indicators maintaining a relatively constant and significant effect across study periods: the proportion of immigrants, the proportion of individuals working outside their residency town, and the proportion of families without a child. The protective effect of families without a child may be related to the family size (i.e. number of family sources) or the lack of Covid-19, thus limiting household transmissions [18]. Our results are concordant with other findings that identified higher risk of Covid-19 infection among immigrants [19]. The remaining indicators were either significant for a single period, or with changing directions relative to the risk of infection.

The association between education level and infection risk at an ecological level suggests a less at-risk behaviour within units with a higher proportion of high education individuals during the Growth period. This may be associated with the spatial distribution of elevated IRRs during that time, as higher education levels tend to be observed in urban areas. This could also be associated with a higher mobility and social activities of more educated individuals, especially during the summer and long vacation periods in a similar fashion as what Plümper and Neumayer [20] reported during the first COVID-19 wave in Germany.

A behavioural mechanism may also be at play regarding unemployment rate. This indicator is considered as a risk factor for worst health outcomes and is a component of several deprivation indexes [21,22]. However, in our study the unemployment rate was found to be associated to a lower risk of infection during the Peak and decrease period. The unemployment may be acting as a proxy for social isolation in this work more than a deprivation measure, as discussed by Scarpone et al. [23]. It is in contrast with the role of single-person households, associated with an excess of risk solely during the Peak and decrease period, when this indicator is expected to be a protective one, since it captures the absence of household contamination risks.

The changing estimated role of home overcrowding is surprising, as this indicator is firmly associated with deprivation and worst health outcomes [9,24–27], and is a proxy of promiscuity. As such, the excess risk found for the Low incidence period is coherent with previous literature, and suggest the interest of directing prevention and infection control resources toward those highly vulnerable households in the early phase of an epidemic wave.

In this study, a limited association between the socio-economic indicators and the rate of SARS-CoV-2 cases was found at the ecological level. Although several indicators were found to be associated with the incidence rate at the IRIS level, it is dubious that any form of inference can be conducted regarding the risk at the individual level. Previous studies on COVID-19 and other infectious diseases clearly suggests that the socio-economic level of an individual is associated with the risk of infection [6,7,10,28,29]. Those results are in line with the more general association between socio-economic position and multiple health outcomes such as all-cause mortality or prevalence of chronic diseases [30–32].

The present findings suggest that one must proceed with care in interpreting the results of ecological studies involving an infectious disease and socio-economic indicators when working with small-area units (around 1,000 inhabitants). As shown in the present study, models which did not account for the spatial autocorrelation of incidence rates failed to sufficiently explain the observed distribution of cases. In contrast, the overall strong spatial correlation associated with a low variance weighting that was found suggests that during the second wave of the COVID-19 pandemic in the study region, the stronger determinant of incidence rate in an IRIS was the incidence rates in its close neighbours, measurably above all the socio-economic indicators studied in this research. From a planning perspective, this result indicates the necessity of a cartographic approach to the allocation of infection control resources or implementation of measures to limit contacts (i.e. curfew) in an emergent pandemic situation more than a socio-economic-based approach at the population level. The present literature remains scarce regarding small-area modelling of infectious disease, limiting our ability to compare the present results appropriately. We identified a handful of authors reporting on small-area analysis about COVID-19 incidence, but without providing a quantitative estimation of the explanatory power of the analysis [33–35]. One study conducted in New York found a significant association between testing for infection and socio-economic position with an estimated R² around 0.3, but with spatial units ranging in population between 10^3^ and 10^5^ [36], compromising comparability with our work.

Our study has several limitations. First, for reasons of personal data protection, we were unable to conduct an analysis standardized for age and sex. More than 30% of cases observed during the entire study period would have been censored. Estimates of age-related risk of infection suggests a upward trend in risk with increasing age [37], indicating a potential excess of cases in units where the elderly are overrepresented, and the opposite in areas inhabited by a younger population. With regards to the sex-related risk, other studies found variable results, with a tendency toward a higher risk of positivity among males compared to females [38,39].

Our second caveat is the absence of availability of the counts of individuals tested for SARS-CoV-2 infection. Indeed, assuming a homogeneous testing rate across all spatial unit is a strong hypothesis [40]. Moreover, similar work on French data has established a variation of the testing rate according to socio-economic level, with a decrease in testing with the increase in deprivation [35]. It is therefore reasonable to suspect the counts of infection cases to be underestimated in more deprived areas, which could lead to bias the estimates of the association between incidence rates and socio-economic indicators.

In conclusion, although this study identified several socio-economic indicators associated with the risk of SARS-CoV-2 infection, they did not prove sufficient to properly explain the spread of the virus during the second wave of the pandemic in one French region. Response to an emerging disease with human to human transmission should be organised around proper cartographic data to quickly identify excesses of cases in low incidence phases and to mitigate the impact of untargeted civil liberties restrictions.

## Statements & Declarations

## Funding

The authors declare that no funds, grants, or other support were received during the preparation of this manuscript.

## Competing interests

The authors have no relevant financial or non-financial interests to disclose.

## Author Contributions

All authors contributed to the study conception and design. Material preparation, data collection and analysis were performed by Nicolas ROMAIN-SCELLE. The first draft of the manuscript was written by Nicolas ROMAIN-SCELLE and all authors commented on previous versions of the manuscript. All authors read and approved the final manuscript.

## Ethics approval

We used only aggregated and statistically censored data for the purpose of this analysis. No ethic committee was solicited for the purpose of this study.

## Data accessibility

The infection cases data that support the findings of this study are available from the French Health Data Hub within the SI-DEP database. Restrictions apply to the availability of these data, which were used under licence for this study. Data may be requested by the following procedure: https://www.demarches-simplifiees.fr/commencer/soumission-d-un-projet-de-recherche-etude-ou-evalu.

The census data that support the findings of this study are openly available on the INSEE website at the following links: https://www.insee.fr/fr/statistiques/4799268, https://www.insee.fr/fr/statistiques/4799323, https://www.insee.fr/fr/statistiques/4799252, https://www.insee.fr/fr/statistiques/4799305.

The administrative layout data that support the findings of this study are openly available on the INSEE website at the following link: https://www.insee.fr/fr/information/2017499

The geographical data that support the findings of this study are openly available on the Geofabrik website at the following links: https://download.geofabrik.de/europe/france.html

Codes and specific datasets (with the exception of the infection cases dataset) are available on request to the corresponding author.

## Acknoledgments

We thank the DATA direction at Santé publique France for providing us with the *ad hoc* dataset, and Dr. Christine Saura from Santé publique France for her imput during the elaboration of this study.

## Notes

### Competing Interest Statement

The authors have declared no competing interest.

