## Supplementary material for "Usefulness of ecological mobility and socio-economic indicators in SARS-CoV-2 infection modelling: a French case study": S1 - Covariates description.pdf

### Covariates definitions and description

Table 1: covariates labels and descriptions

| Covariate | Shorthand | Formula | Definitions | Map |
|---|---|---|---|---|
| Number of individuals at risk | Population | *Population* | | Figure 1 |
| Population density index (4 levels) | Density | *Population.density.index* | Cf. Table 1 | Figure 2 |
| % of immigrants | % migrants | Prop.migrant.indiv | Proportion of non French nationals over all inhabitants | Figure 3 |
| % of individuals unemployed | % unemployed | Prop.unemployed.indiv | Proportion of unemployed individuals over all residents between 15 and 64 | Figure 4 |
| % of single person homes | % single | Prop.indiv.living.alone | Proportion of homes with a single inhabitant over all homes | Figure 5 |
| % of individuals working outside their residency town | % out of resid. town | Prop.indiv.working.outside.residency.town | Proportion of individuals working outside of their residency town over all residents between 15 and 64 | Figure 6 |
| % of individuals travelling to work by car | % work by car | Prop.indiv.going.work.by.car | Proportion of individuals travelling to work by car over all residents between 15 and 64 | Figure 7 |
| % of families without a child | % no child | Prop.families.without.child | Proportion of families without a child over all families | Figure 8 |
| % of car ownership | % own a car | Prop.car.ownership | Proportion of homes possessing at least one car over all homes | Figure 9 |
| % of low education | % low educ. | Prop.indiv.under.high.school.diploma | Proportion of individuals not in education without a high school diploma over all individuals not in education | Figure 10 |
| % of overpopulated homes | % overpop. home | Prop.overcrowded.homes | Proportion of homes with more than one inhabitant for each room (excluding the bathroom) over all homes | Figure 11 |

Back to top

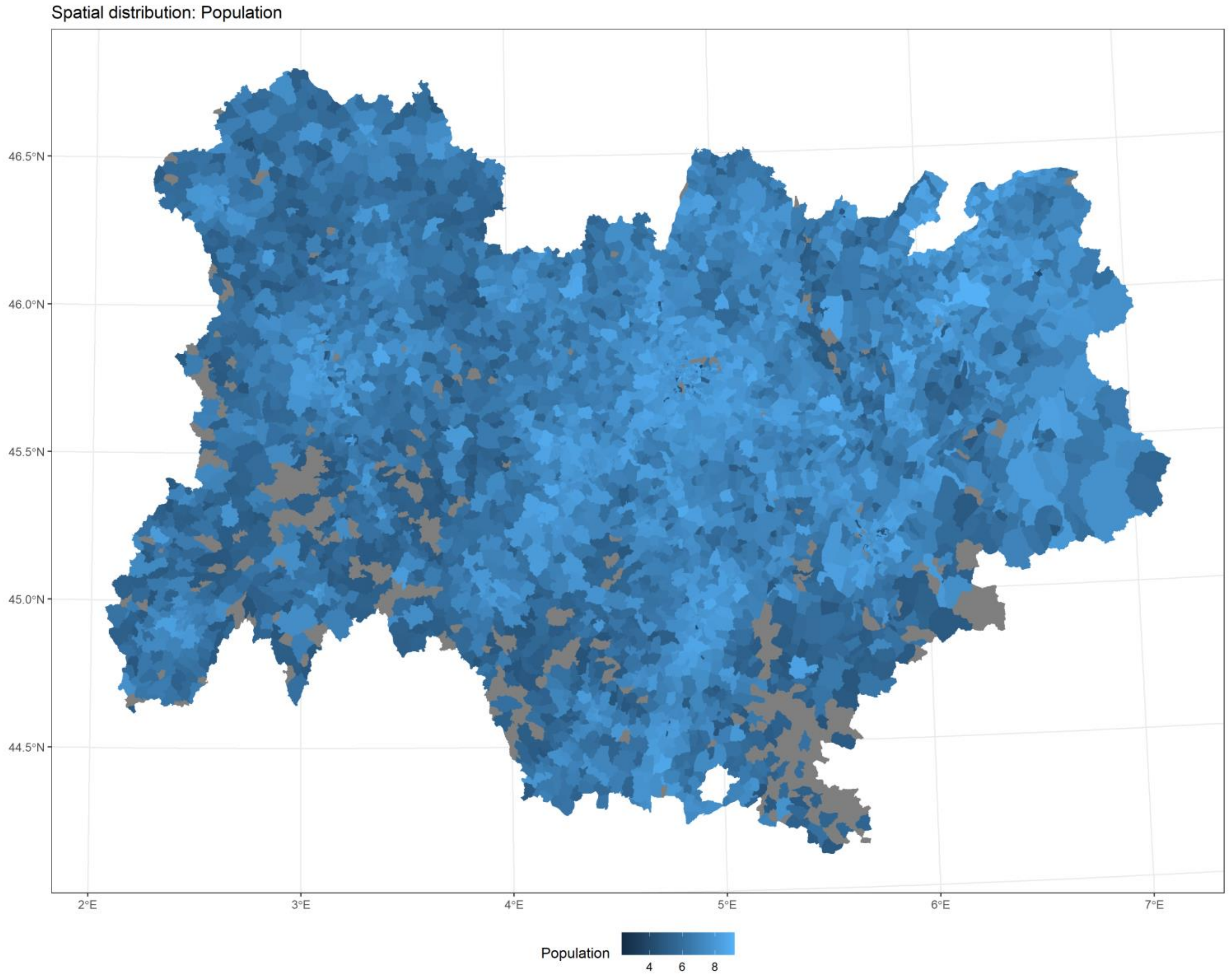


*Figure 1*

[Back to top](#)

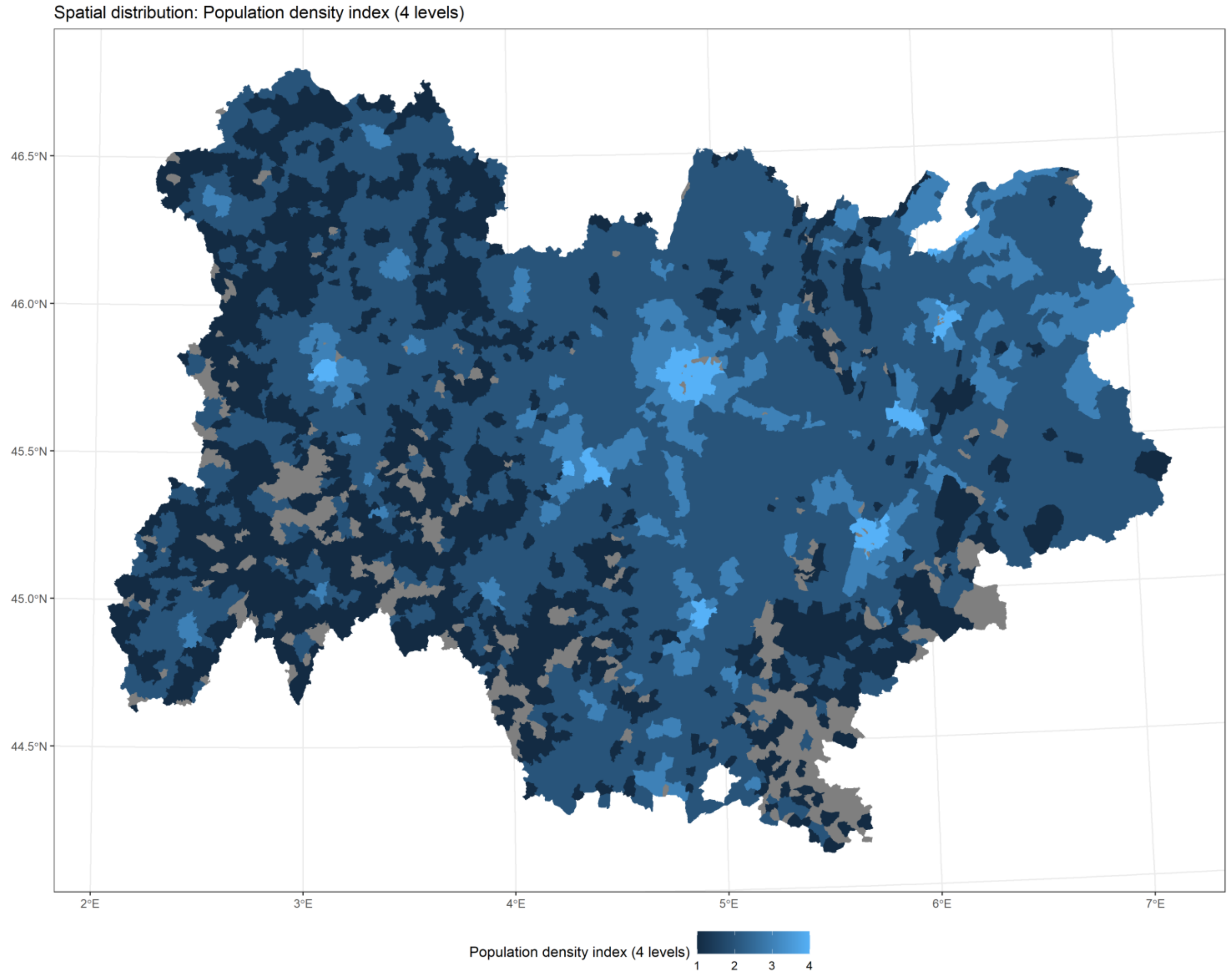


*Figure 2*

Back to top

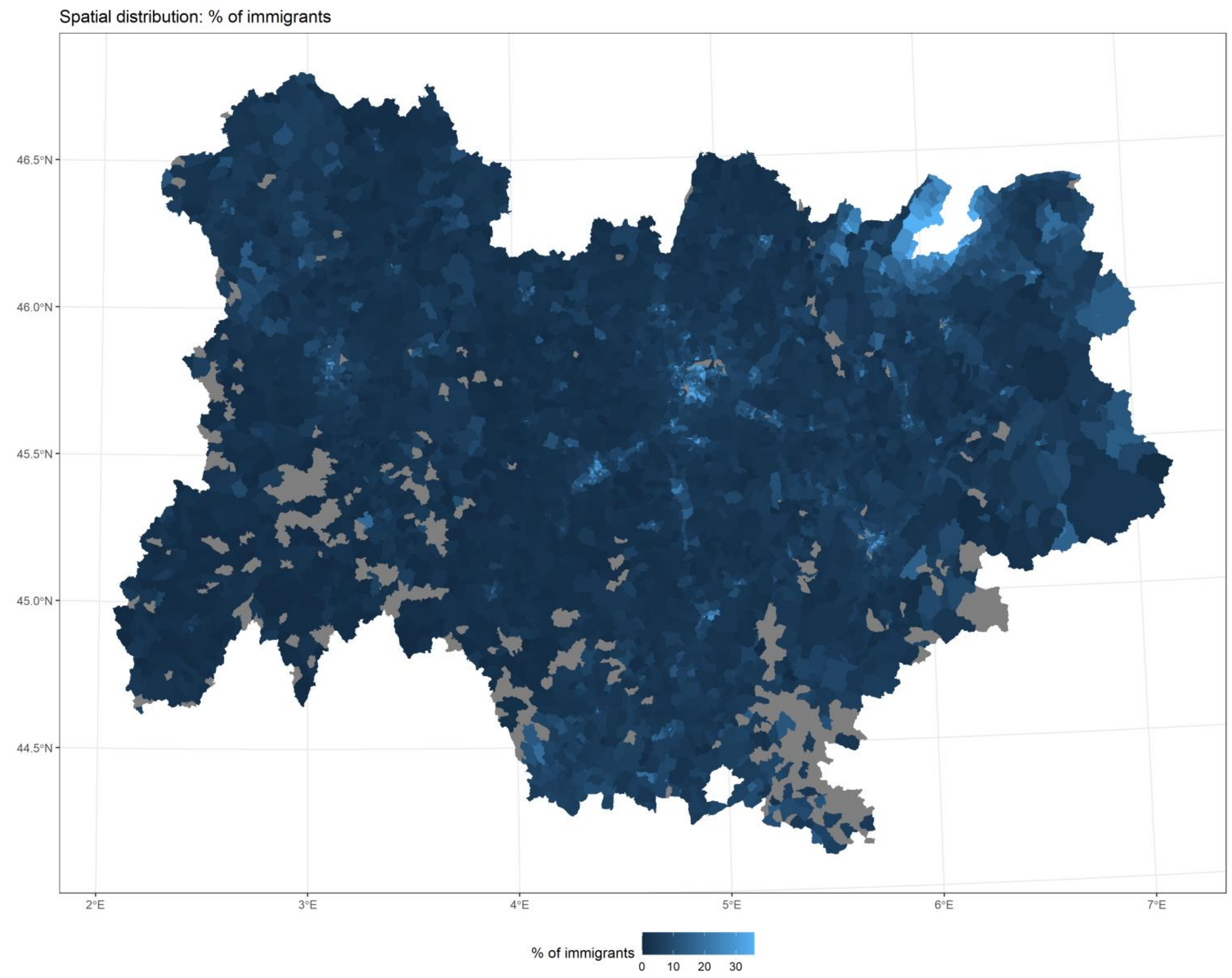


*Figure 3*

Back to top

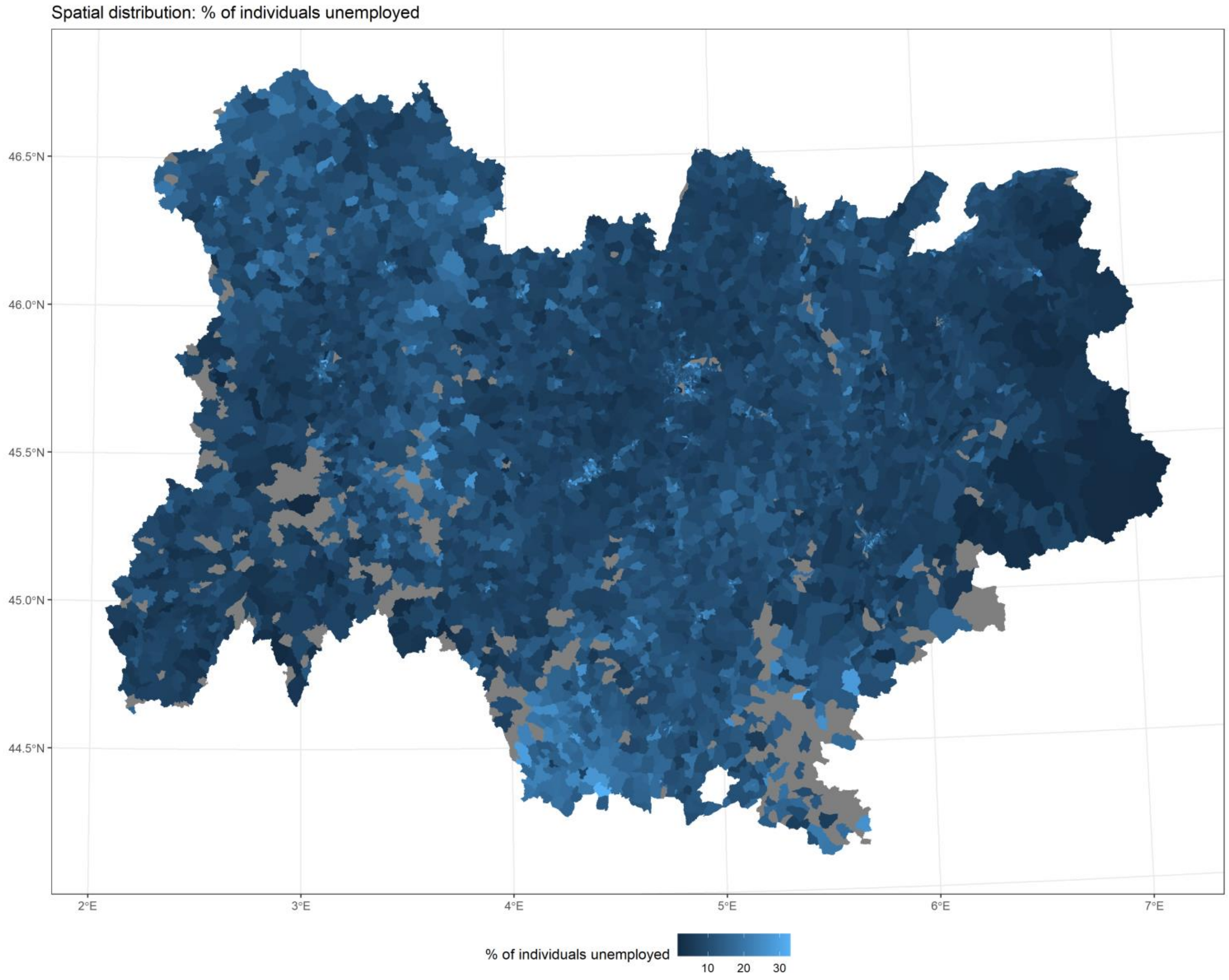


*Figure 4*

[Back to top](#)

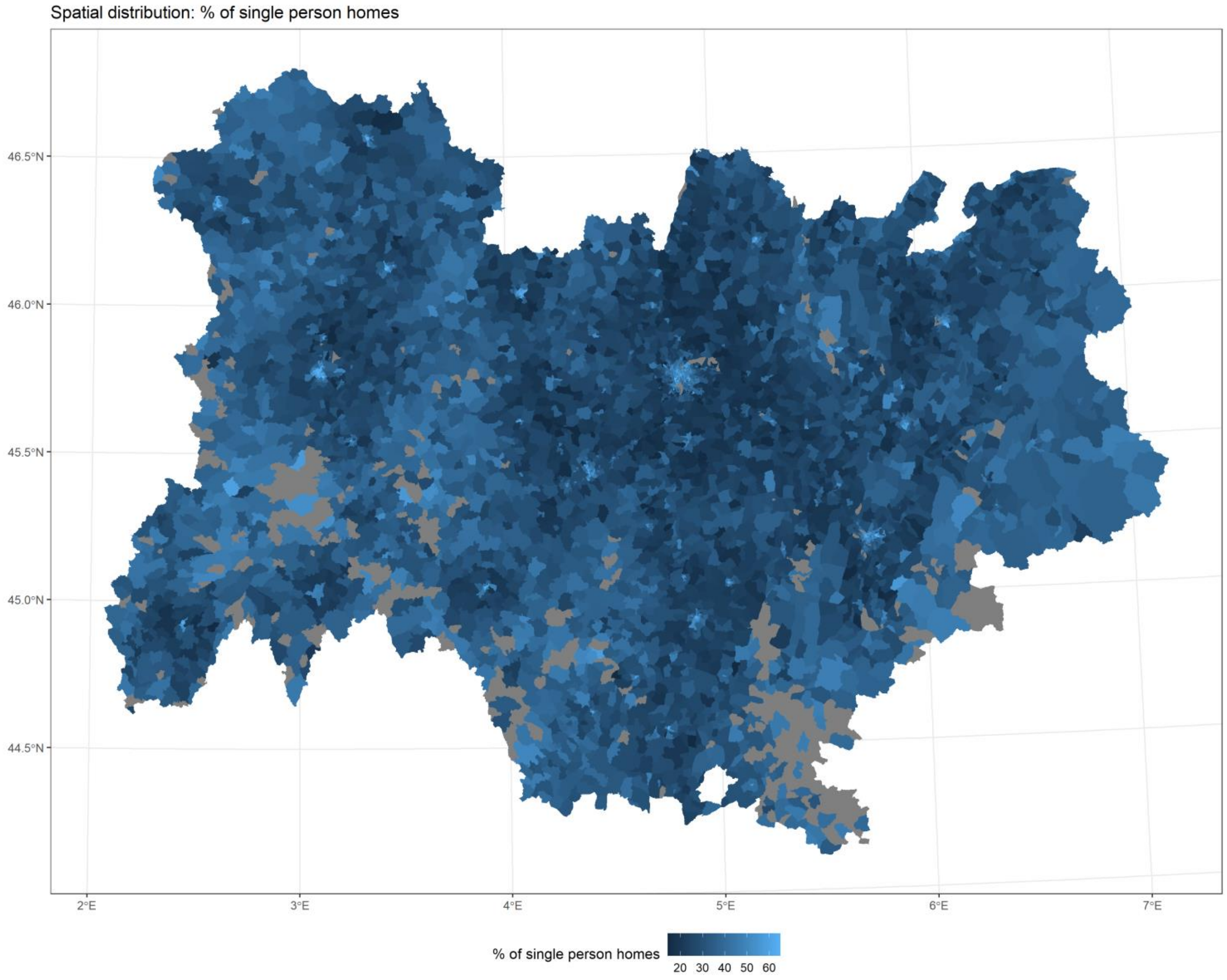


*Figure 5*

[Back to top](#)

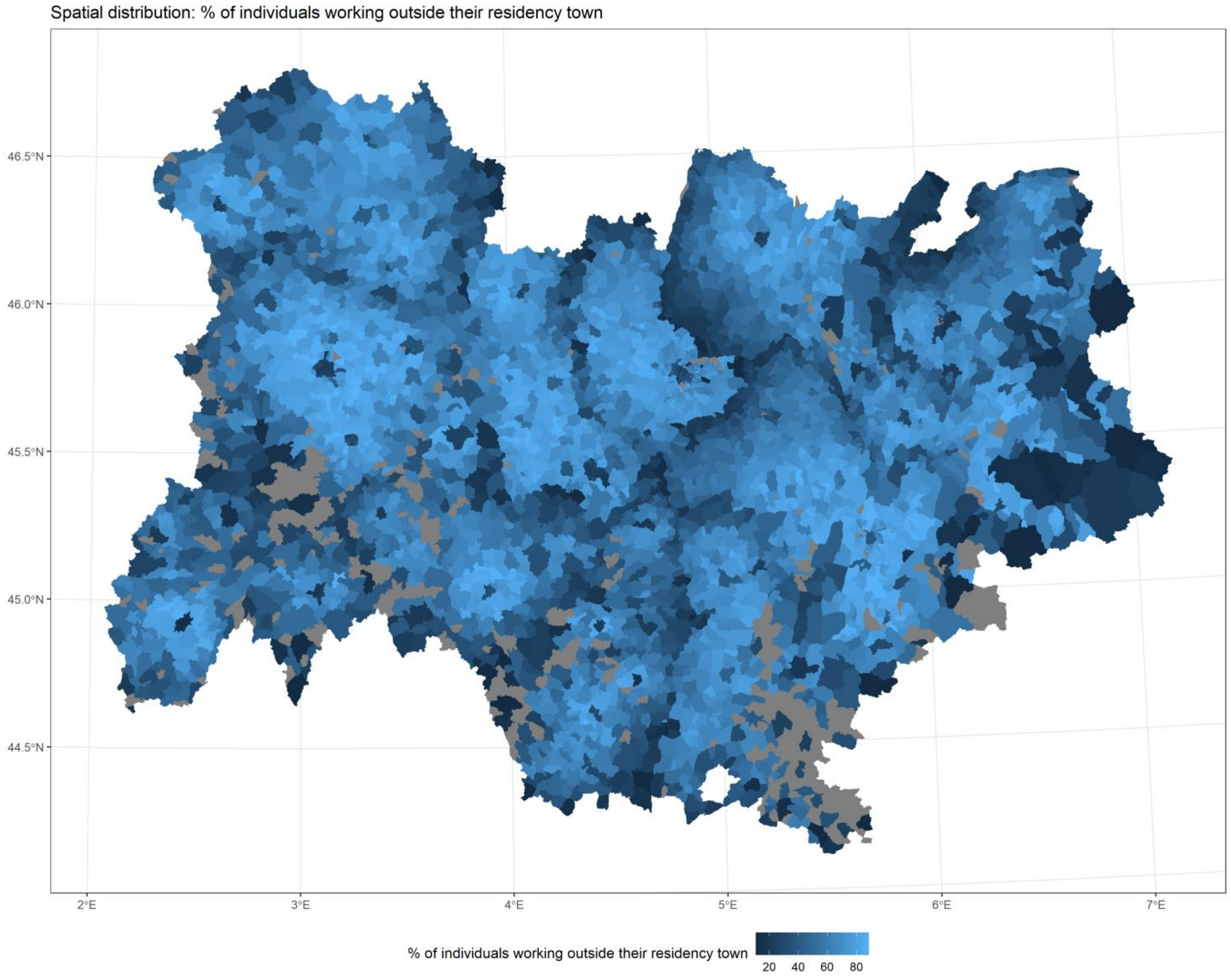


*Figure 6*

[Back to top](#)

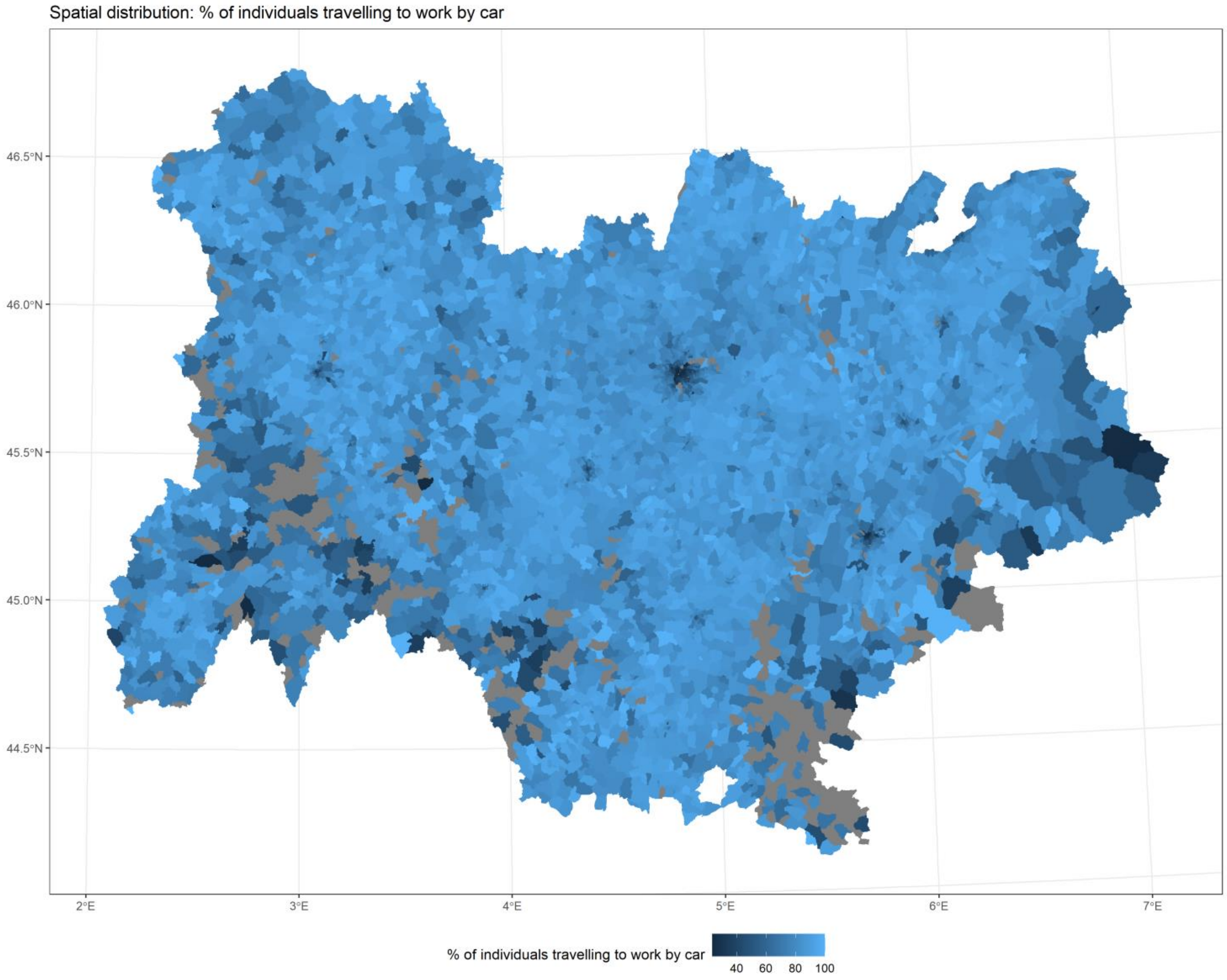


*Figure 7*

[Back to top](#)

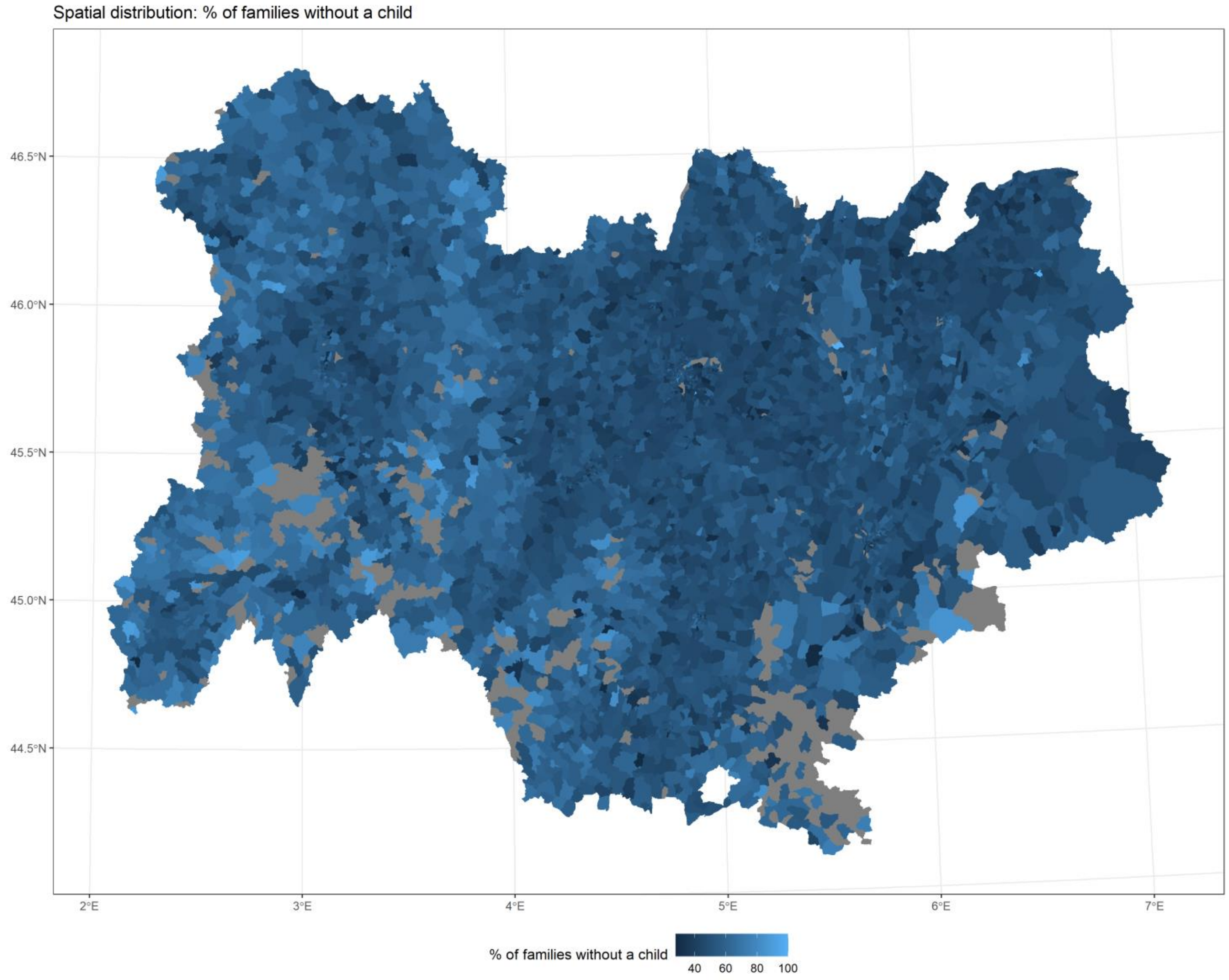


*Figure 8*

Back to top

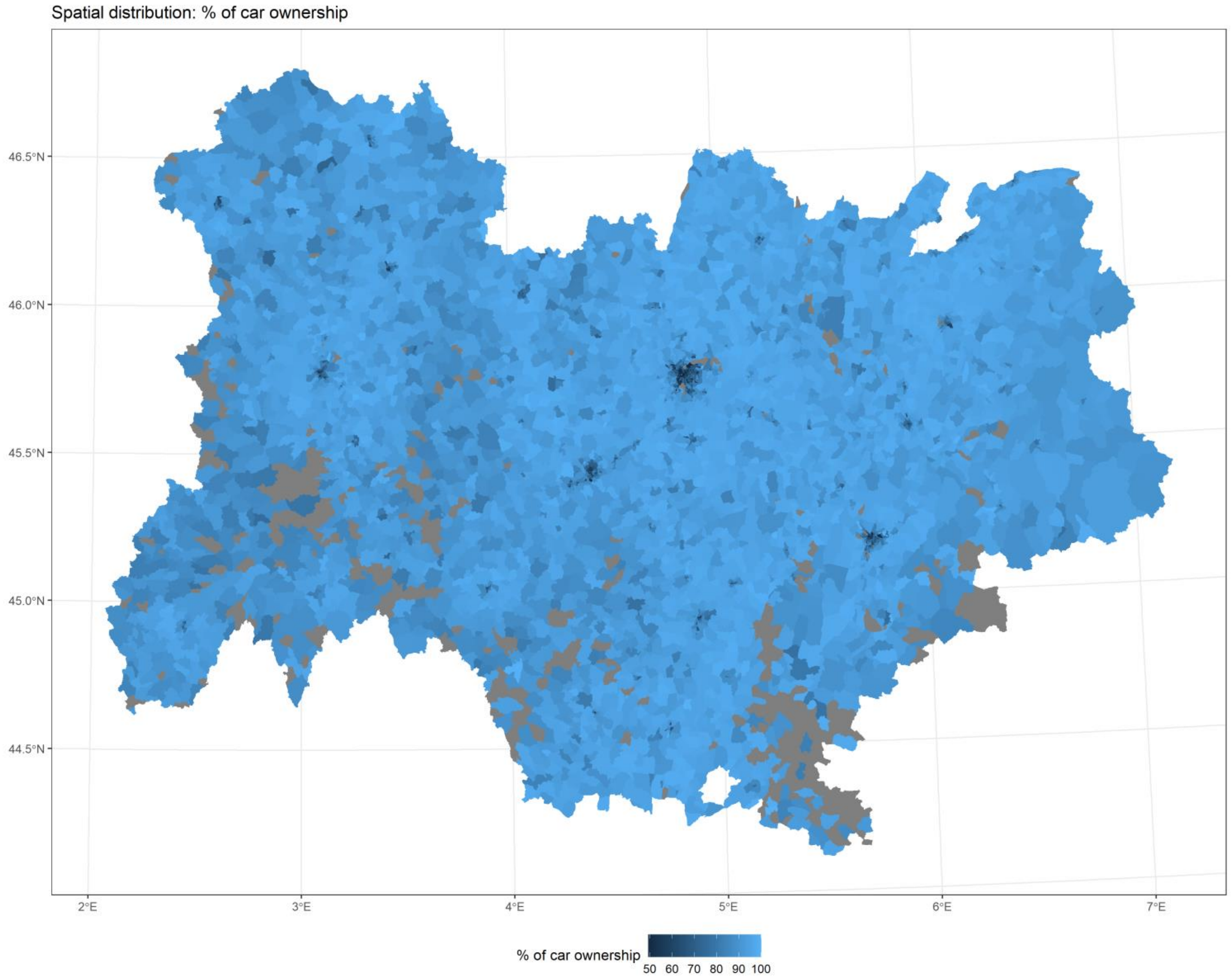


*Figure 9*

[Back to top](#)

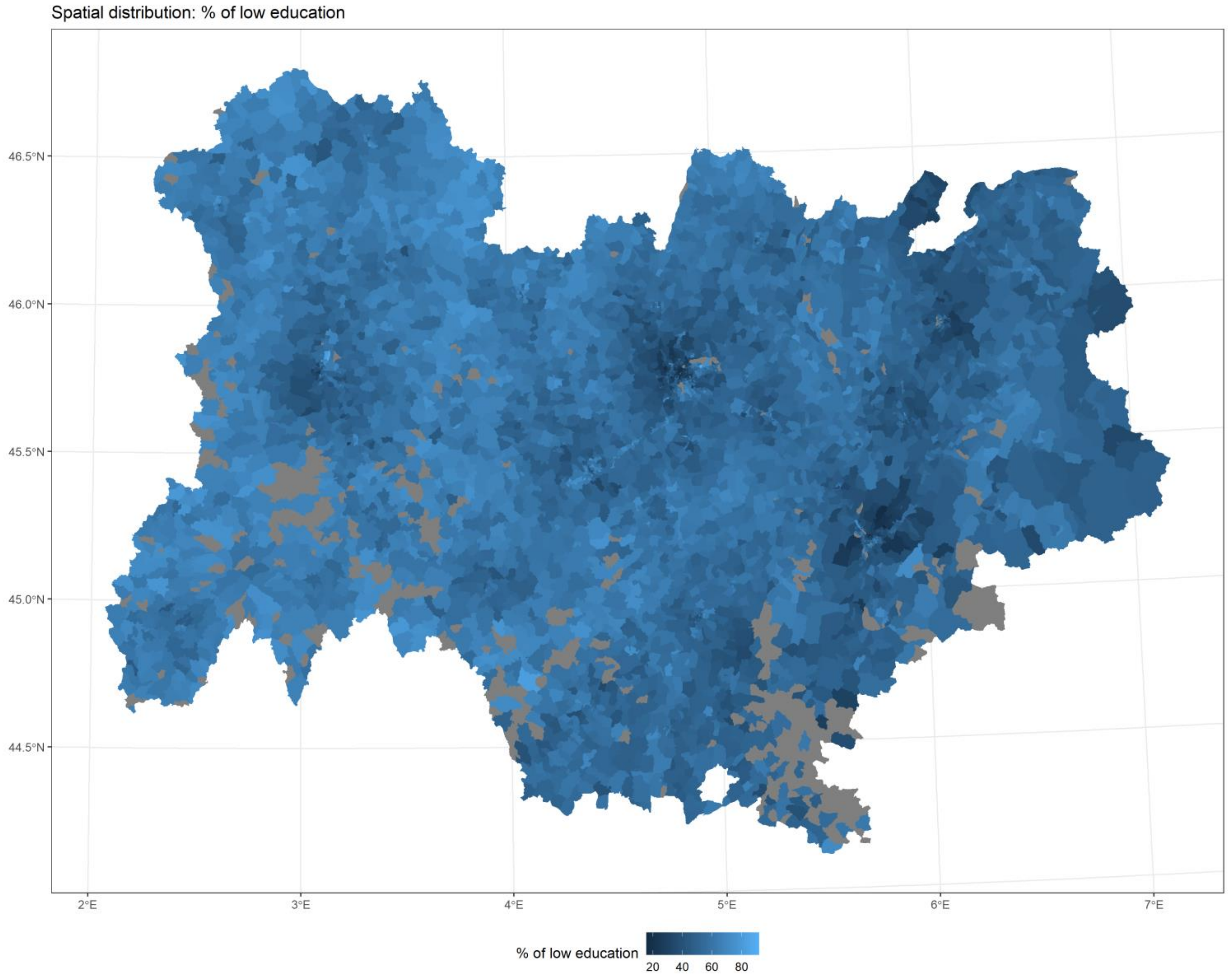


*Figure 10*

[Back to top](#)

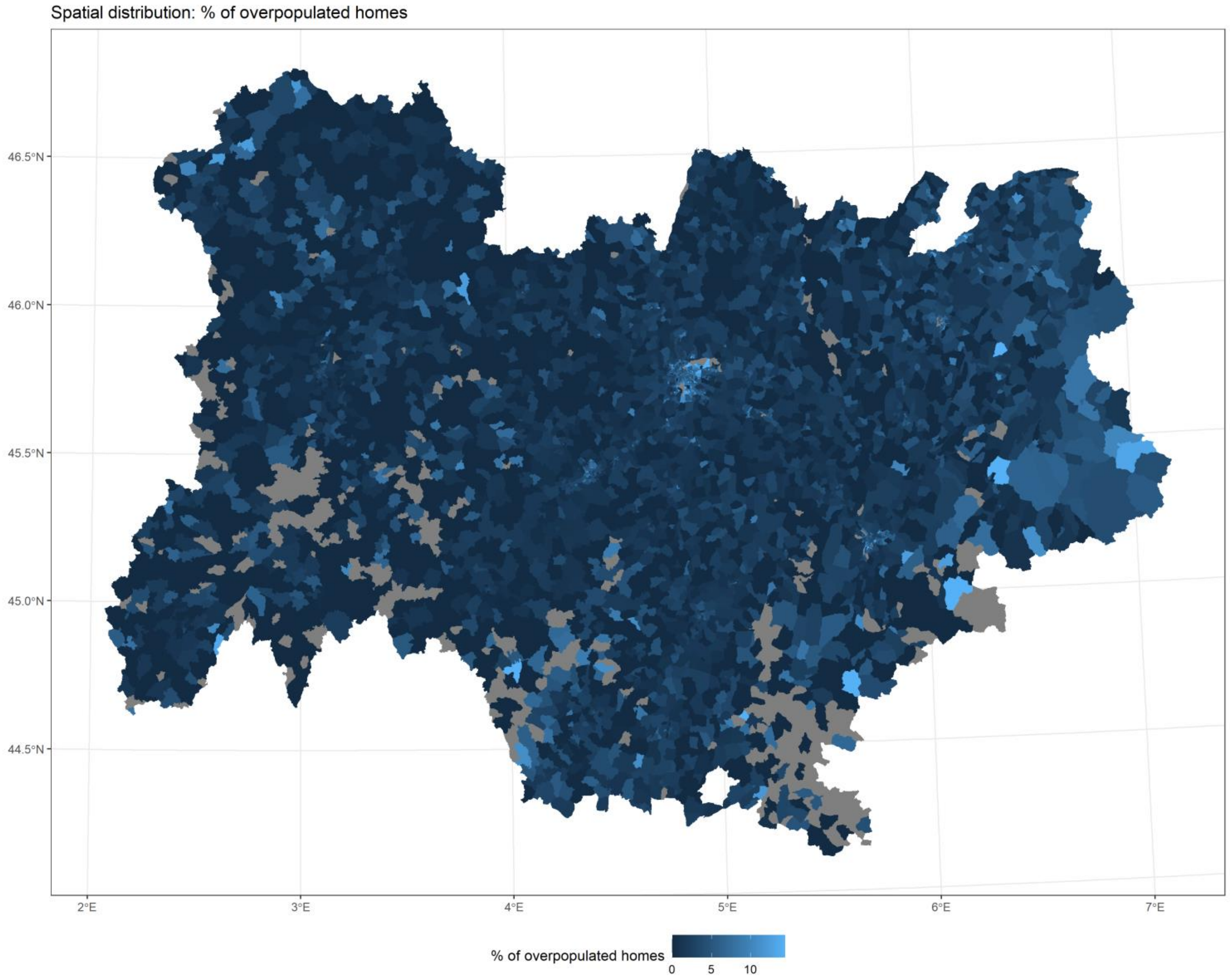


*Figure 11*

[Back to top](#)
