## Supplementary material for "Usefulness of ecological mobility and socio-economic indicators in SARS-CoV-2 infection modelling: a French case study": S2 - Statistical methods.pdf

### General model specification

In the present report, the following model was used to assess the relationship between the aggregated case count of SARS-CoV-2 infection and covariates, accounting for spatial effects:

$$Y_k \sim Pois(\lambda_k) \tag{1}$$

$$\log(\boldsymbol{\lambda}) = \boldsymbol{\mu} + \boldsymbol{X\beta} + \boldsymbol{\phi} \tag{2}$$

$$\boldsymbol{\phi}|\boldsymbol{W}, \tau^2, \rho \sim N(0; \tau^2[\rho \boldsymbol{W} + (1-\rho)\boldsymbol{I_K}]^{-1}) \tag{3}$$

With $\boldsymbol{\lambda}$ the vector of $\lambda_k$ the expected count for unit *k*, $\boldsymbol{\beta}$ the vector of covariate parameters of length $p$, $\boldsymbol{X}$ the covariate matrix of dimensions $p \times K$, $k \in \{1, \dots, K\}$ index of a given spatial unit, $\phi_k$ element *k* of $\boldsymbol{\phi}$ the vector of random spatial effect, $w_{kj}$ row of $\boldsymbol{W}$ the spatial weight matrix of dimensions $K \times K$, $\tau^2$ the variance weighting parameter, $\rho$ the spatial correlation coefficient, $\boldsymbol{I_K}$ the identity matrix. In the present report, we refer to the $\boldsymbol{X\beta}$ component as the covariate component, and $\boldsymbol{\phi}$ as the spatial effect component.

### Fixed effects formula

The $\boldsymbol{X\beta}$ component introduced in Equation 2 in the main body of this report can be detailed as such, for one spatial unit *k*:

$$\begin{aligned}\boldsymbol{X\beta} = {} & \beta_1\, ln(population) + \beta_2\, ln^2(population) \quad + \beta_3\, ln^3(population) \quad + \beta_4\, ln^4(population) \\ & + \beta_5\, ln^5(population) \quad + \beta_6\, I(population.density.index = 1) \\ & + \beta_7\, prop.migrant.indiv + \beta_8\, prop.unemployed.indiv \\ & + \beta_9\, prop.indiv.living.alone \\ & + \beta_{10}\, prop.indiv.working.outside.residency.town \\ & + \beta_{11}\, prop.indiv.going.work.by.car + \beta_{12}\, prop.families.without.child \\ & + \beta_{13}\, prop.car.ownership + \beta_{14}\, prop.indiv.under.high.school.diploma \\ & + \beta_{15}\, prop.overcrowded.homes\end{aligned}$$

The choice of a degree 5 polynomial link between the logarithm of the population-at-risk and the logarithm of the expected number of cases was to attain a complete fitting with no functional form remaining for this specific variable and improve the estimations regarding the other covariates.

### Conditional autoregressive spatial effect

The Equation 3 in the main body introduces the prior for the CAR random effect as a multinomial law with a variance-covariance matrix equal to $\tau^2[\rho \boldsymbol{W} + (1-\rho)\boldsymbol{I_K}]^{-1}$. This random effect can also be specified in a conditional form, as $\phi_k|\phi_{-k}$ with $j \in -k = \{0,1, \dots, k-1, k+1, \dots, K\}$ designating respectively a unit *j* which is not the unit *k* and –*k* the set of indices of all units except *k*.

$$\phi_k|\phi_{-k}, w_{kj}, \tau^2, \rho \sim N\left(\frac{\rho \sum_{j\in-k} \phi_j w_{kj}}{\rho \sum_{j\in-k} w_{kj} + 1 - \rho}; \frac{\tau^2}{\rho \sum_{j\in-k} w_{kj} + 1 - \rho}\right)$$

### Spatial weight matrix

Each cell $w_{kj}$ of the spatial weight matrix $\boldsymbol{W}$ of dimensions $K \times K$ represents the proximity between a unit k and all others units $j \in -k$ with regards to the random spatial effect. Each $w_{kj}$ was computed using an inverse travel-time metric as such:

$$w^*_{kj} = \begin{cases} \frac{1}{t_{kj}}, & t_{kj} < 60 \\ 0, & otherwise \end{cases}$$

$$w_{kj} = \frac{w^*_{kj}}{\sum_k \sum_j w^*_{kj}}$$

with $t_{kj}$ the travel time in minutes between the centroids of unit k and unit j. This provided a total normalized weight matrix $\boldsymbol{W}$ where all row sums are distributed around 1 and symmetry is preserved, which is a computational constraint of the package CARBayes. This also allows a moderately homogeneous interpretation of the parameter $\tau^2$ which must be interpreted for each spatial unit k with respect to the respective $\boldsymbol{W}$ row sum: $\sum_{j\in-k} w_{kj}$.

Travel time between centroids was computed using the Conveyal R5 routing engine (1–4), interfaced with R software through the r5r package (5). Three modes of travel were considered to assess a minimal travel time: bicycle, car, and public transit of all type. The OpenStreetMap geographic dataset was used to provide relevant network information to the travel time matrix computing (6).

### Performance metrics

The McFadden pseudo-$R^2$ is defined as:

$$R^2 = 1 - \frac{log\ \hat{L}(M_i)}{log\ \hat{L}(M_1)}$$

with $M_i$ the evaluated model, $M_1$ the reference model for computation, and $\hat{L}$ the estimated likelihood.

The Moran's I index quantifying the residual influence of units $j \in -k$ upon unit $k$ is computed following the equation:

$$I = \frac{K}{\sum_k \sum_j w_{kj}} \frac{\sum_k \sum_j w_{kj} (\epsilon_k - \bar{\epsilon})(\epsilon_j - \bar{\epsilon})}{\sum_k \sum_j (\epsilon_k - \bar{\epsilon})^2}$$

with $\epsilon_k$ the Pearson standardized residuals.

1. Conway MW, Byrd A, Eggermond M van. Accounting for uncertainty and variation in accessibility metrics for public transport sketch planning. J Transp Land Use [Internet]. 2018 Jul 23 [cited 2022 Jul 27];11(1). Available from: https://www.jtlu.org/index.php/jtlu/article/view/1074

2. Conway MW, Byrd A, van der Linden M. Evidence-Based Transit and Land Use Sketch Planning Using Interactive Accessibility Methods on Combined Schedule and Headway-Based Networks. 2017 [cited 2022 Jul 27]; Available from: https://keep.lib.asu.edu/items/127809

3. Conway MW, Stewart AF. Getting Charlie off the MTA: a multiobjective optimization method to account for cost constraints in public transit accessibility metrics. Int J Geogr Inf Sci. 2019 Sep 2;33(9):1759–87.

4. Conveyal R5 Routing Engine [Internet]. Conveyal; 2022 [cited 2022 Jul 27]. Available from: https://github.com/conveyal/r5

5. Rafael H. M. Pereira, Marcus Saraiva, Daniel Herszenhut, Carlos Kaue Vieira Braga, Matthew Wigginton Conway. r5r: Rapid Realistic Routing on Multimodal Transport Networks with R5 in R. Findings. 2021;

6. OpenStreetMap contributors. France dump retrieved from https://download.geofabrik.de/europe/france.html. 2022.
