## Supplementary material for "Usefulness of ecological mobility and socio-economic indicators in SARS-CoV-2 infection modelling: a French case study": S3 - Models performance.pdf

### Models diagnostics

Residuals: Pearson’s residuals for each spatial unit, model (null model M1, covariates only M2, spatial effects only M3, full model M4) and period (low incidence, growth, peak and decrease, stabilization).

Predictive check: position of the observed incidence for a given spatial unit compared to the predicted values (computed from each chain iteration), for each spatial unit, model, and period. The observed value for the spatial units in red or blue are lower than the 5th percentile or greater than the 95th percentile of the predicted values, indicating an inadequate prediction.

Back to title

Back to title

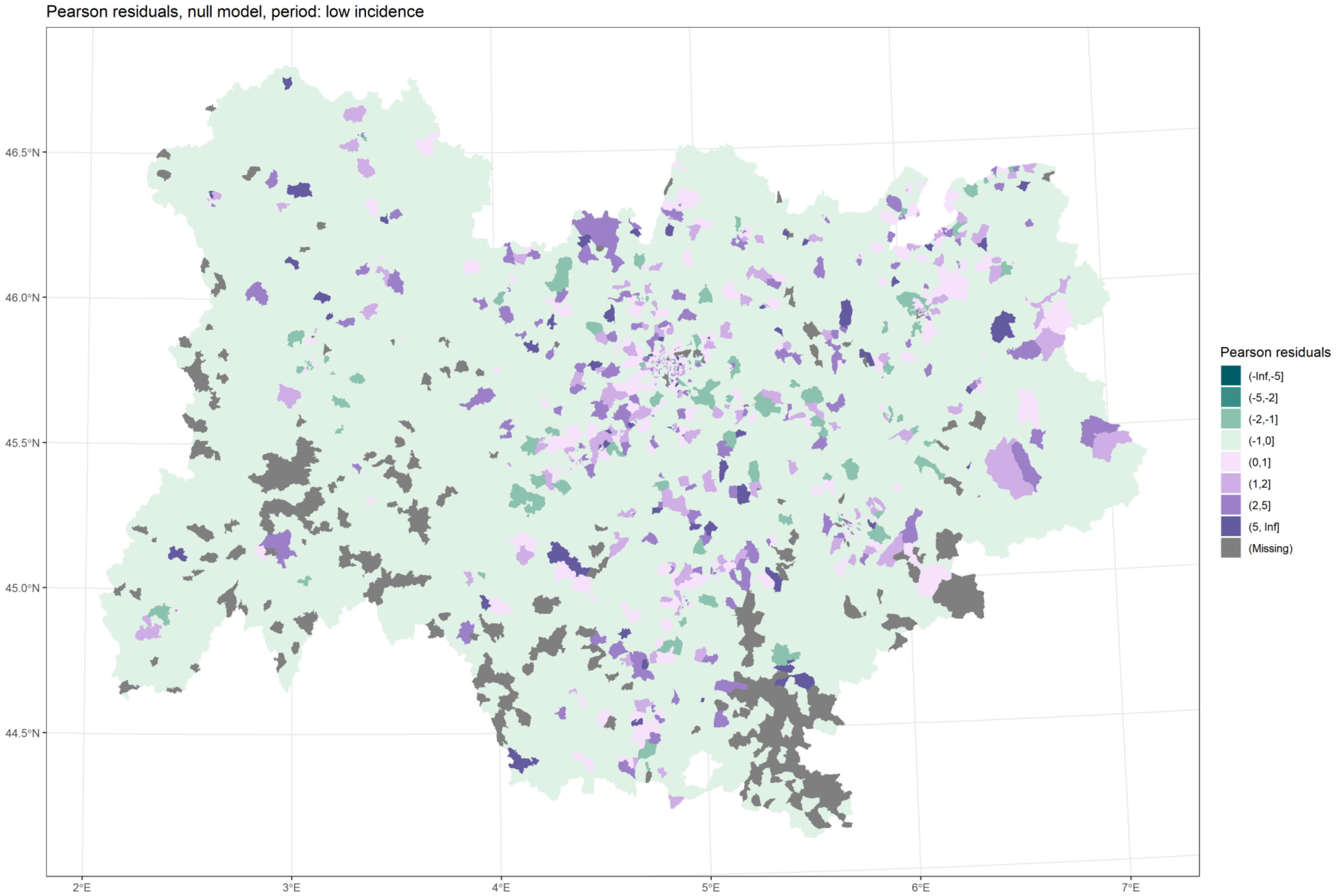

*Figure 1 Residuals, M1, low incidence*

Back to title

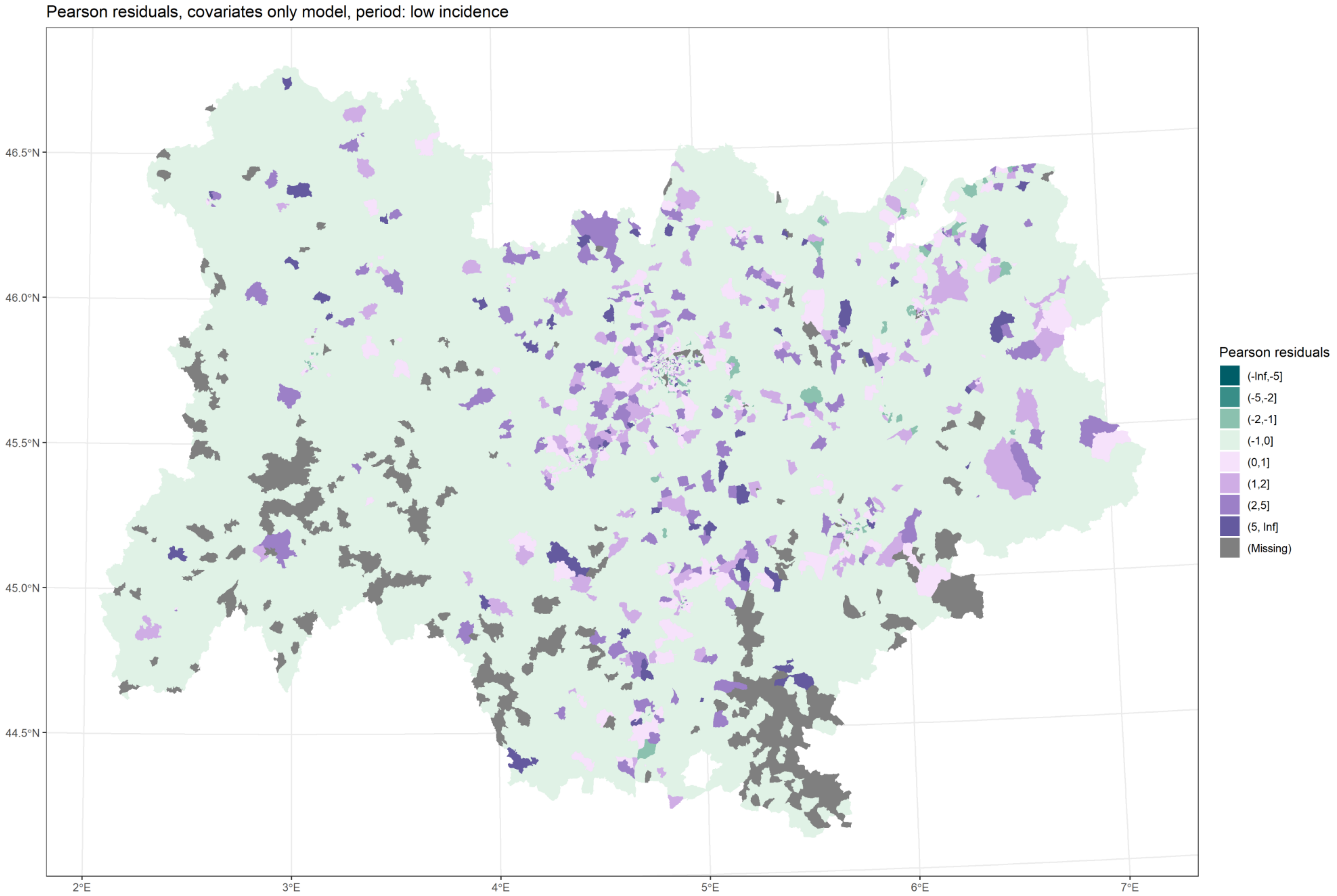

*Figure 2 Residuals, M2, low incidence*

Back to title

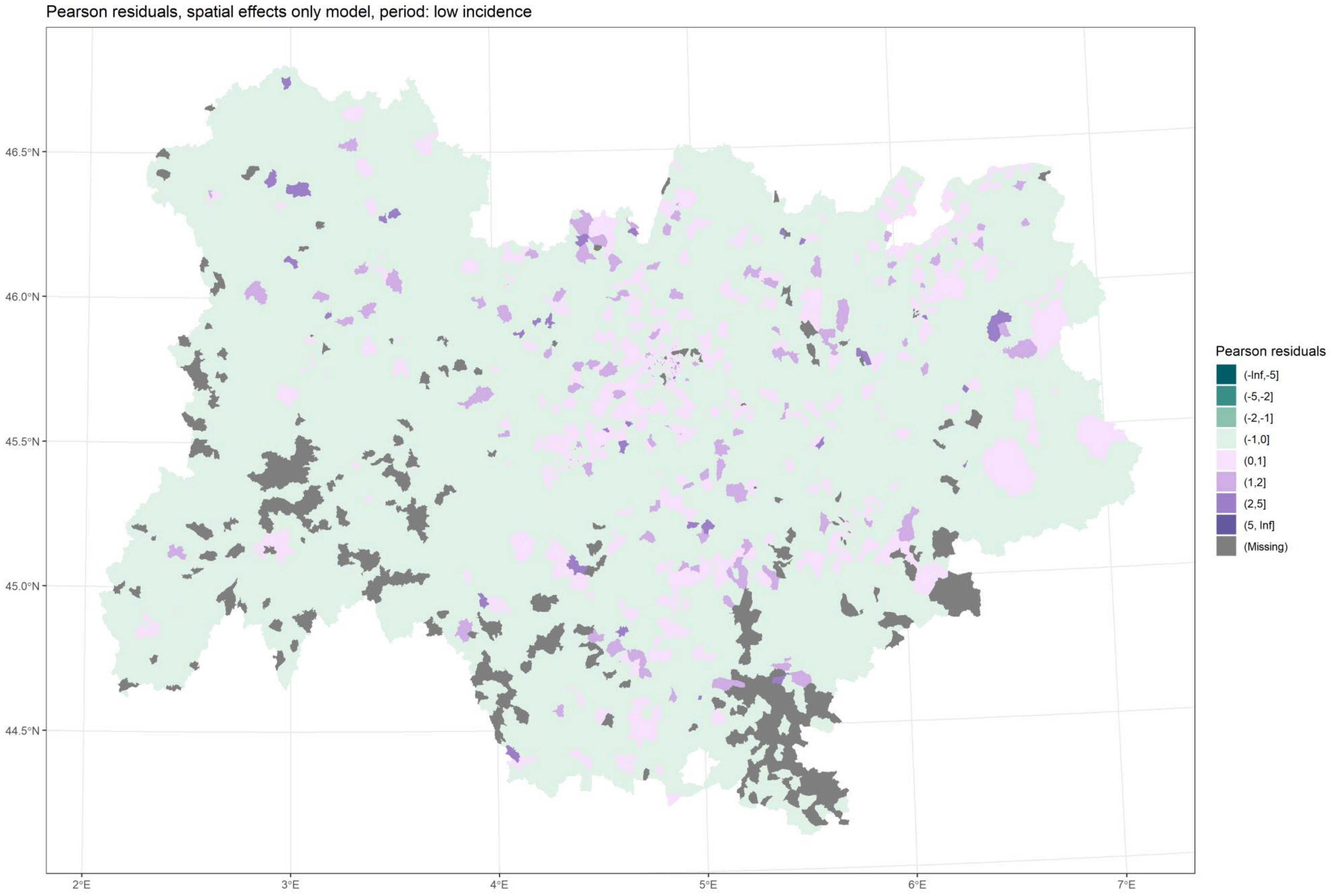

*Figure 3 Residuals, M3, low incidence*

Back to title

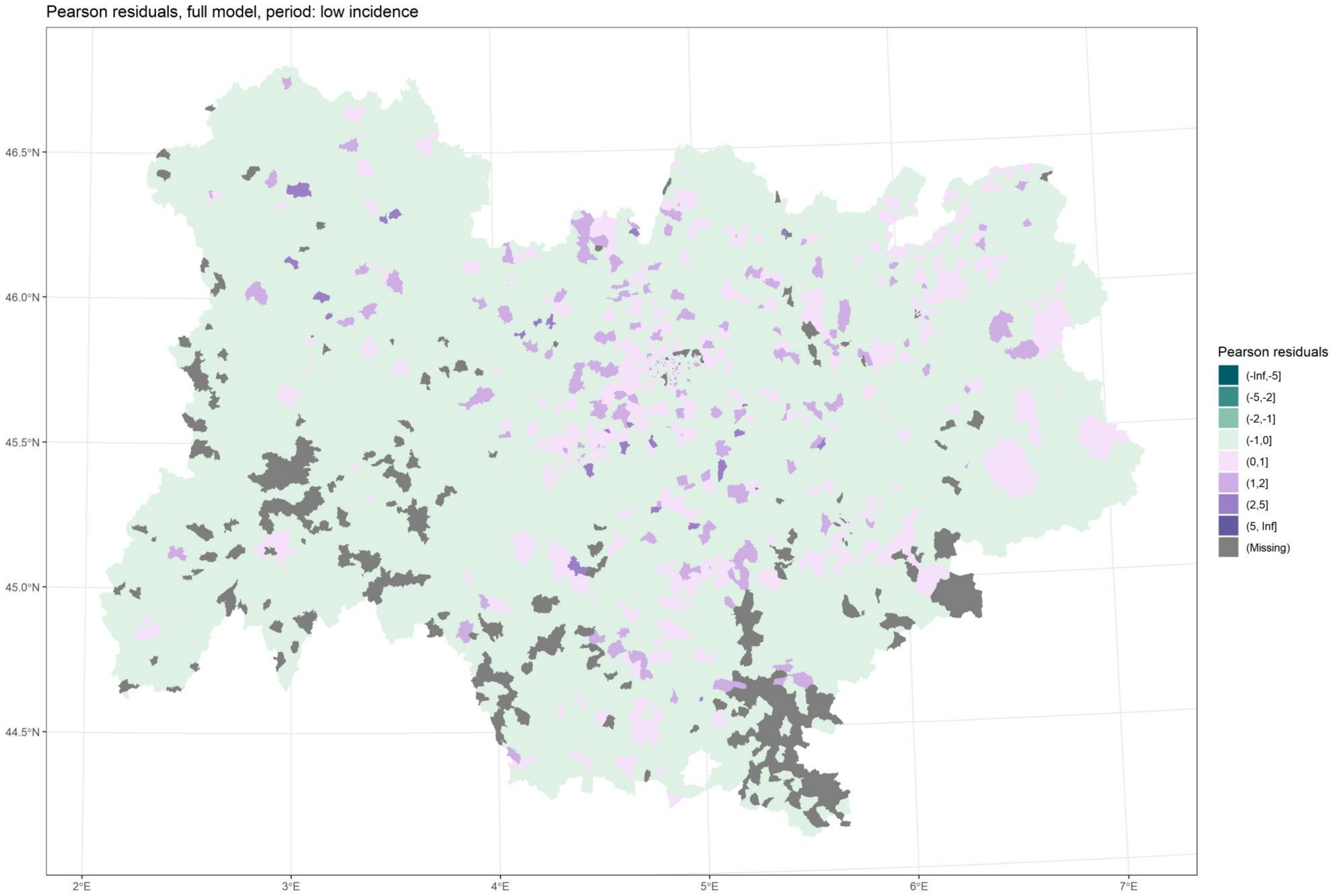

*Figure 4 Residuals, M4, low incidence*

Back to title

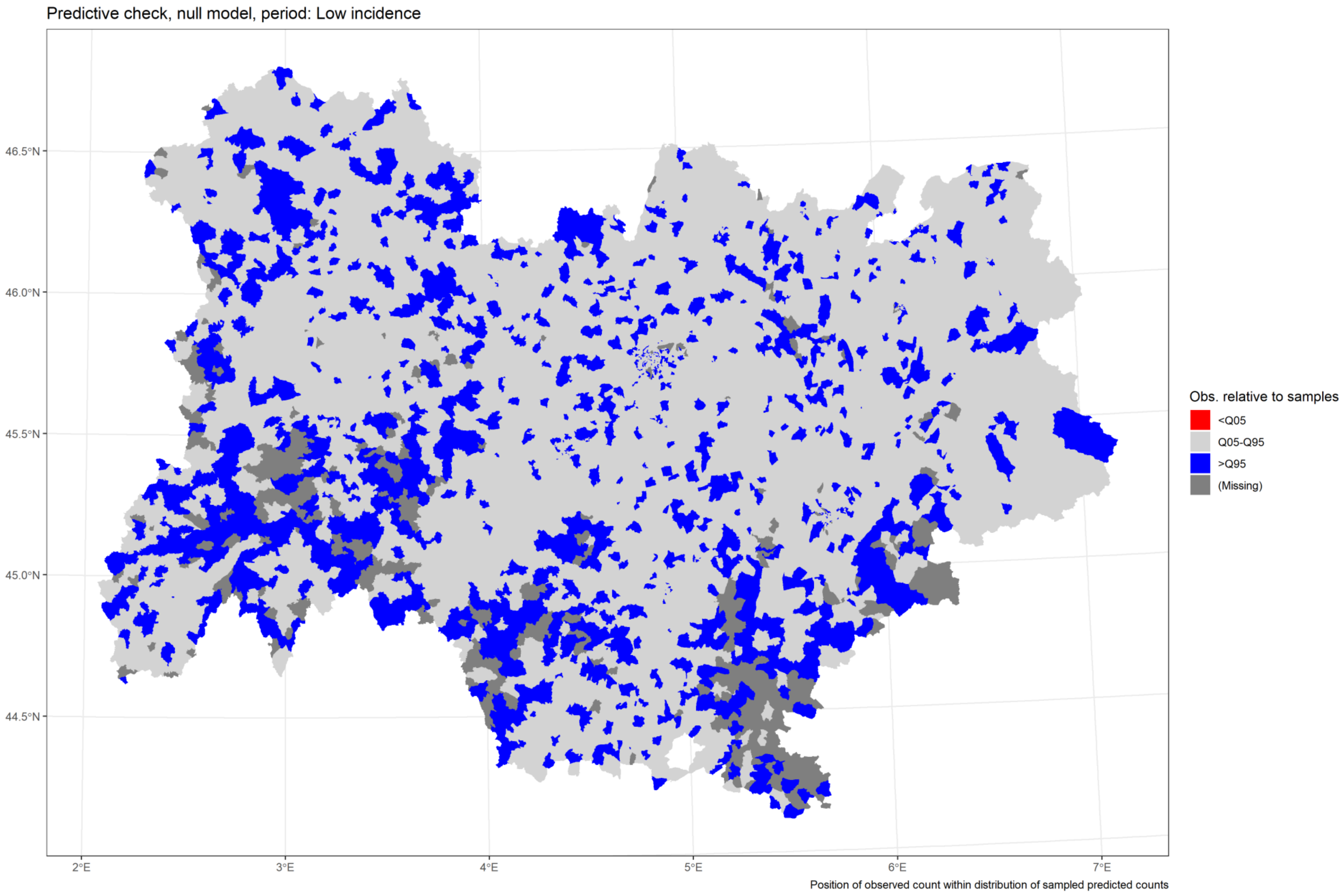

*Figure 5 Predictive check, M1, low incidence*

[Back to title](#)

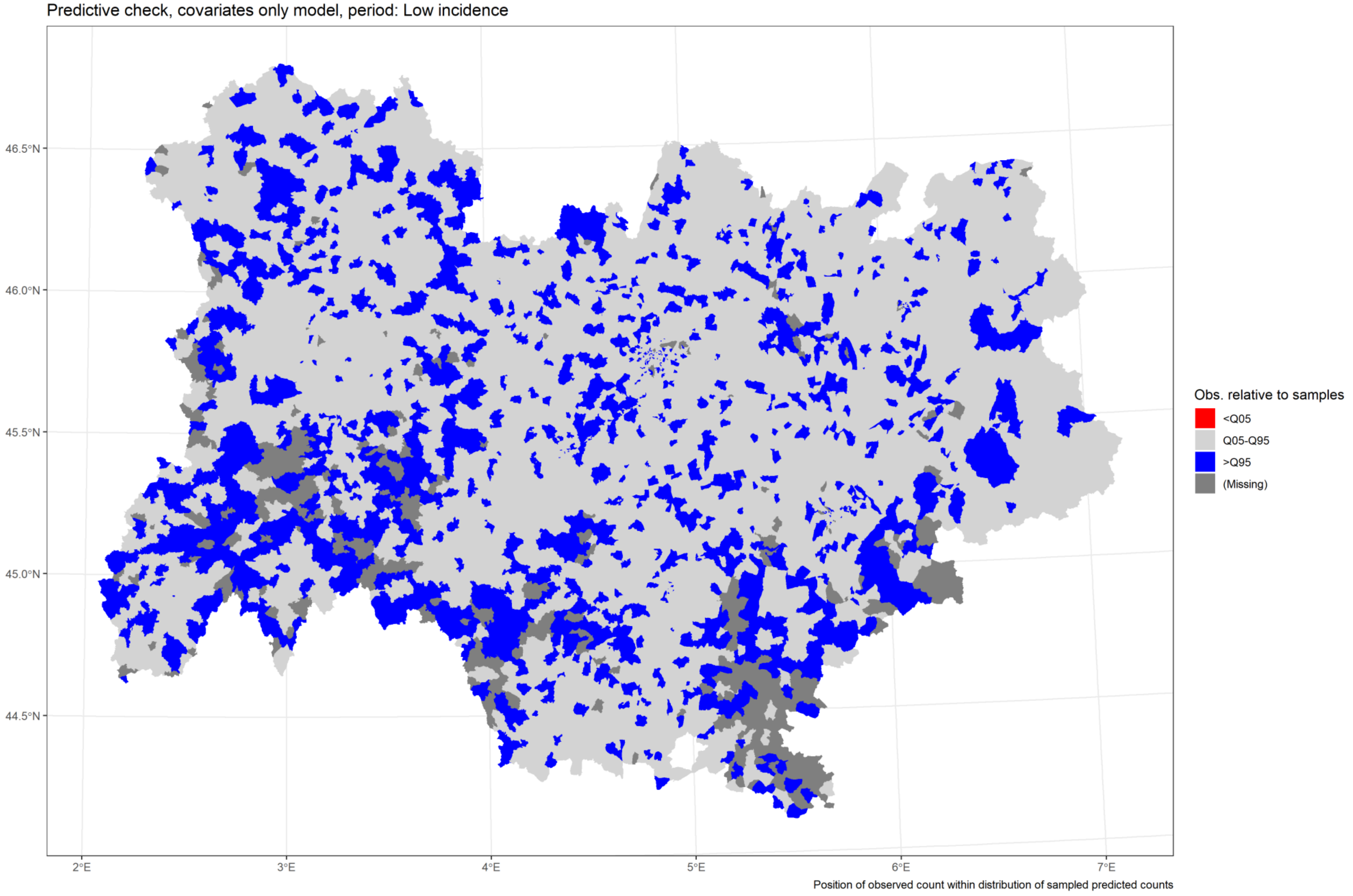

*Figure 6 Predictive check, M2, low incidence*

[Back to title](#)

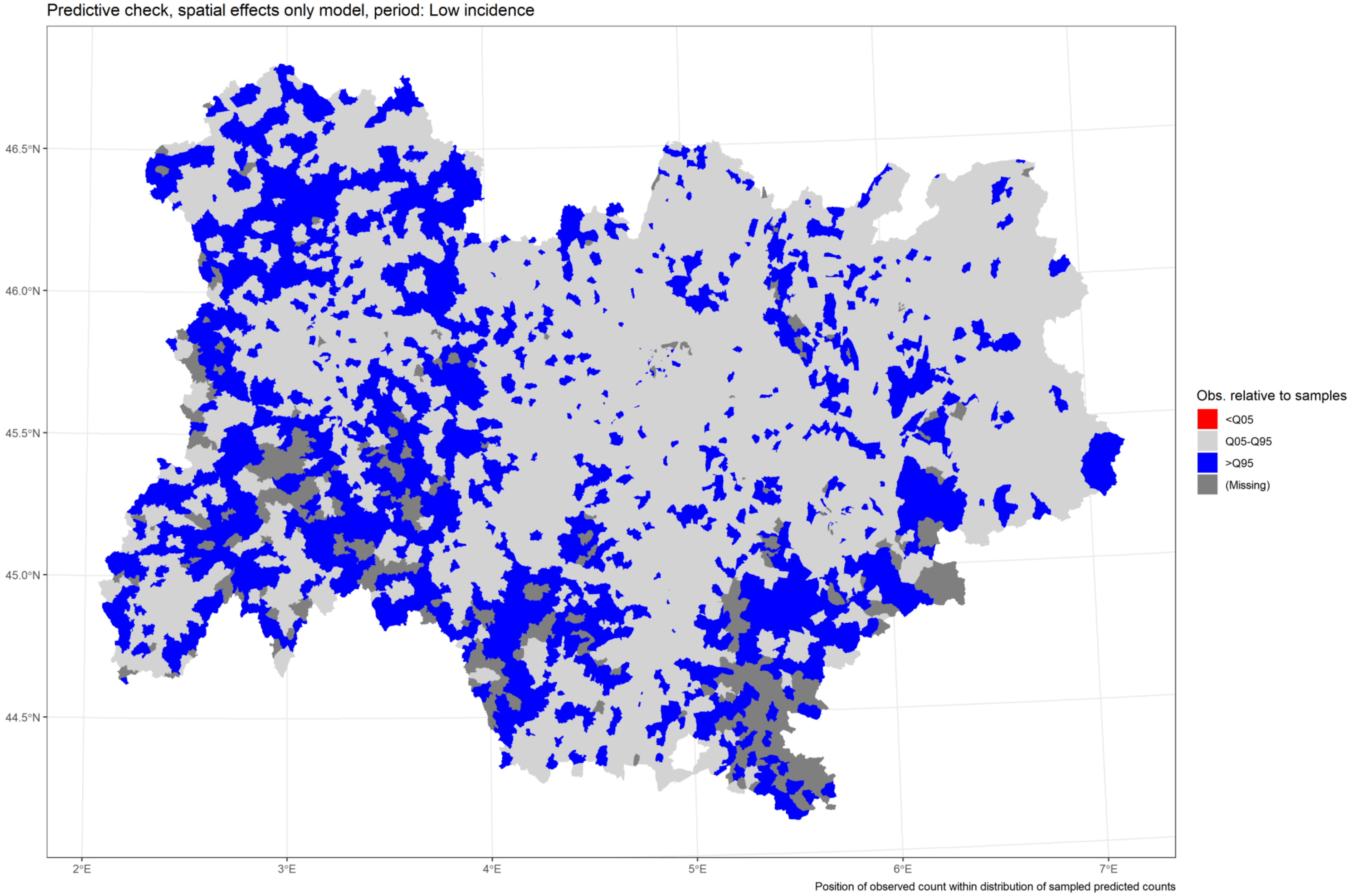

*Figure 7 Predictive check, M3, low incidence*

[Back to title](#)

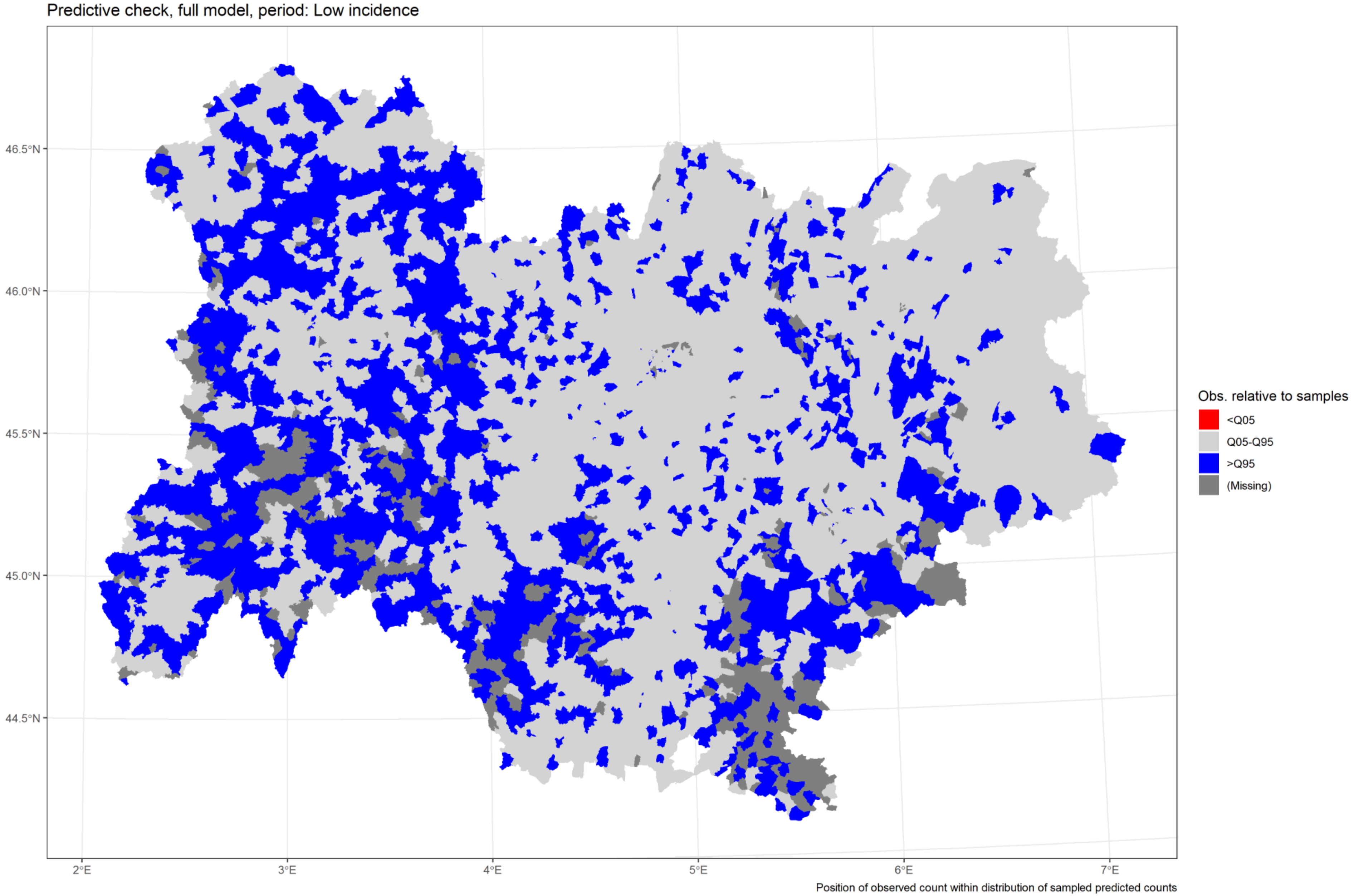

*Figure 8 Predictive check, M4, low incidence*

Back to title

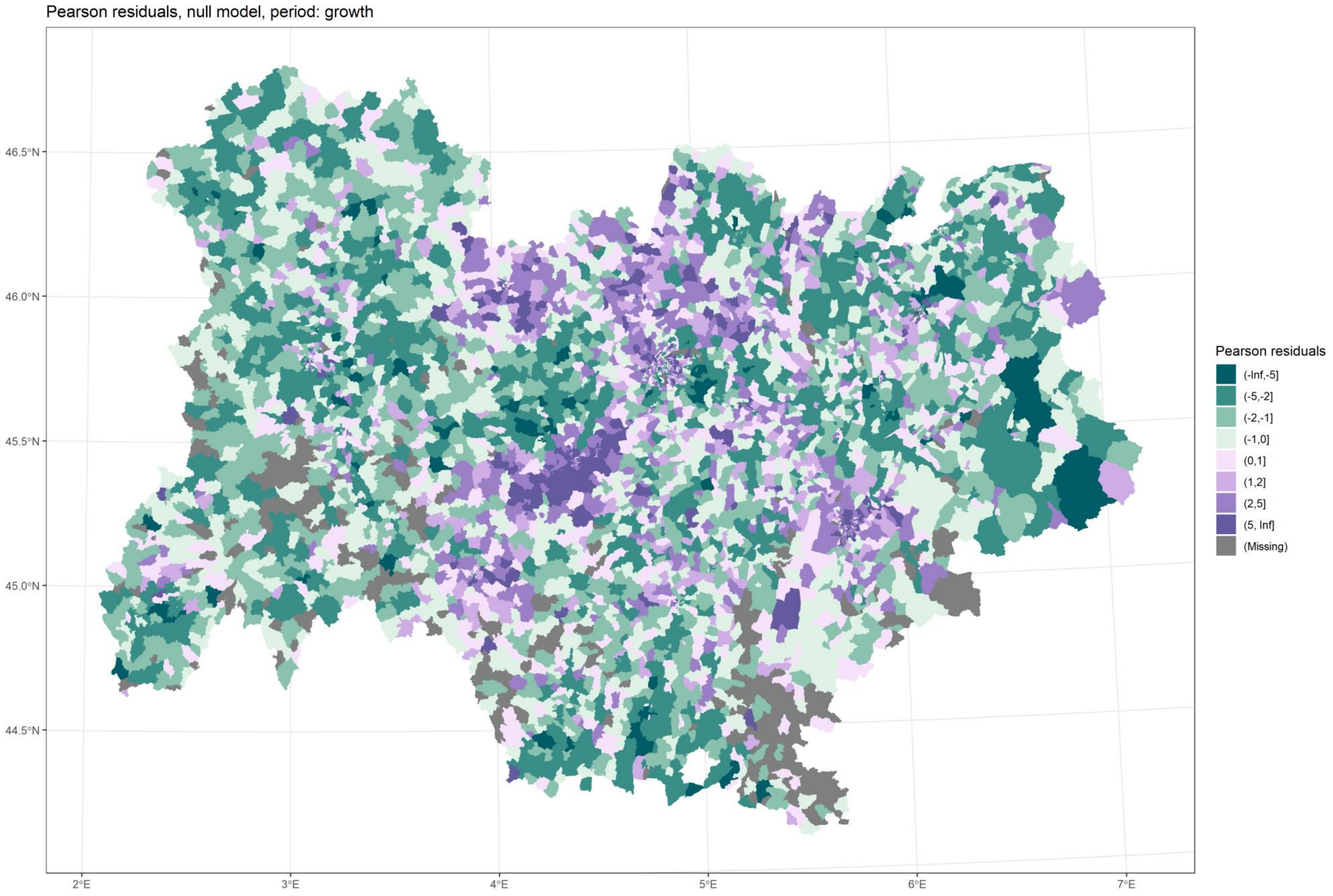

*Figure 9 Residuals, M1, growth*

Back to title

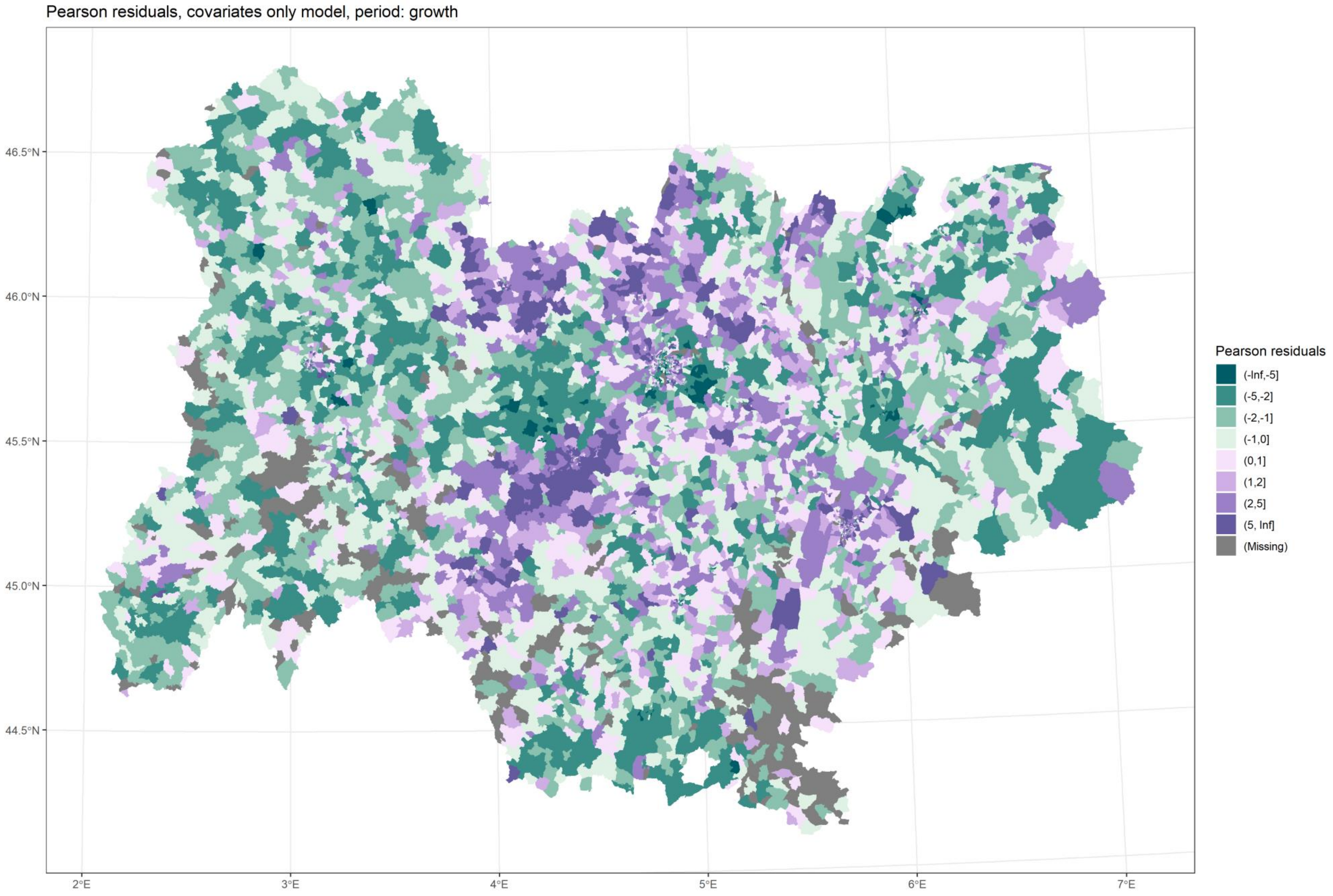

*Figure 10 Residuals, M2, growth*

[Back to title](#)

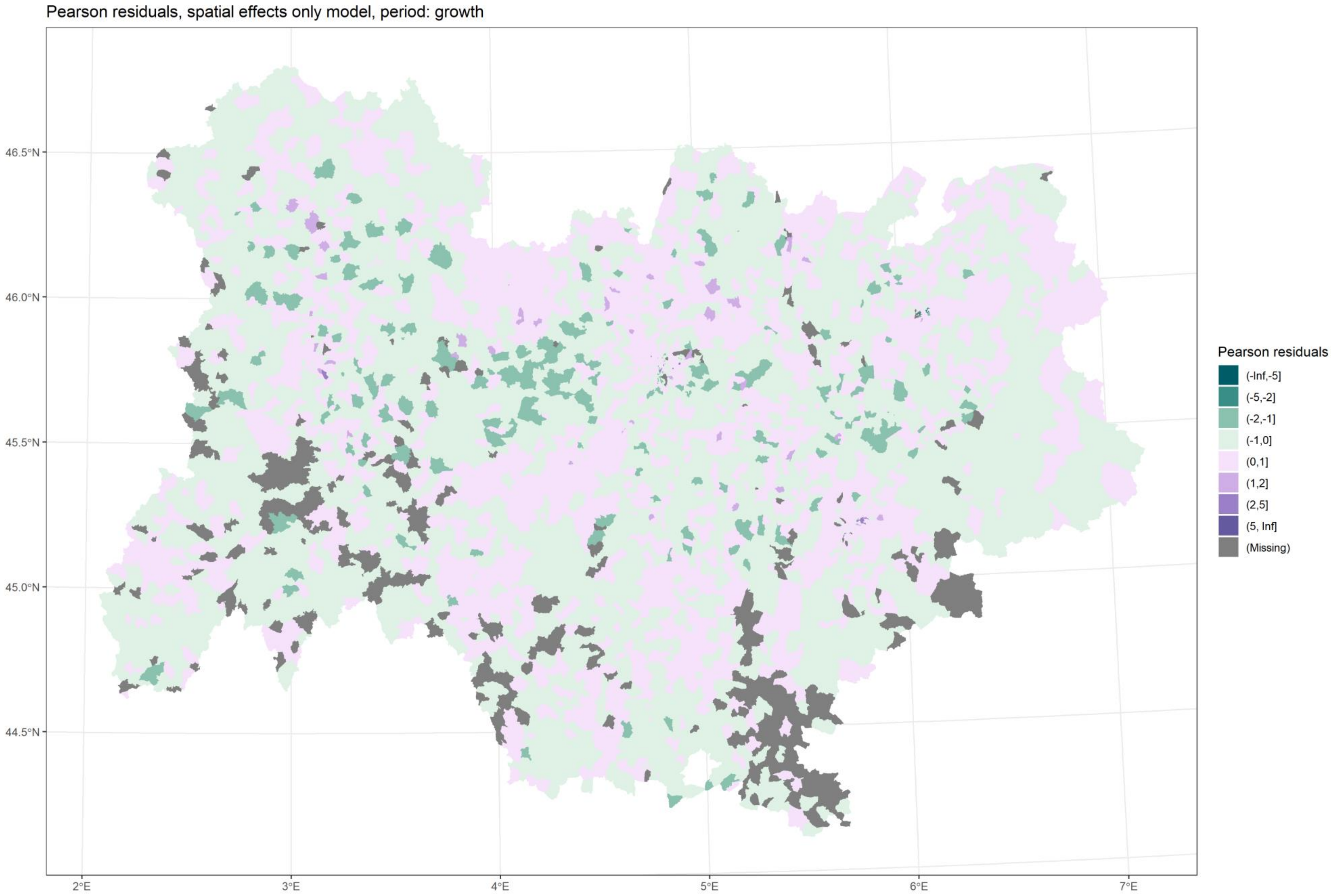

*Figure 11 Residuals, M3, growth*

[Back to title](#)

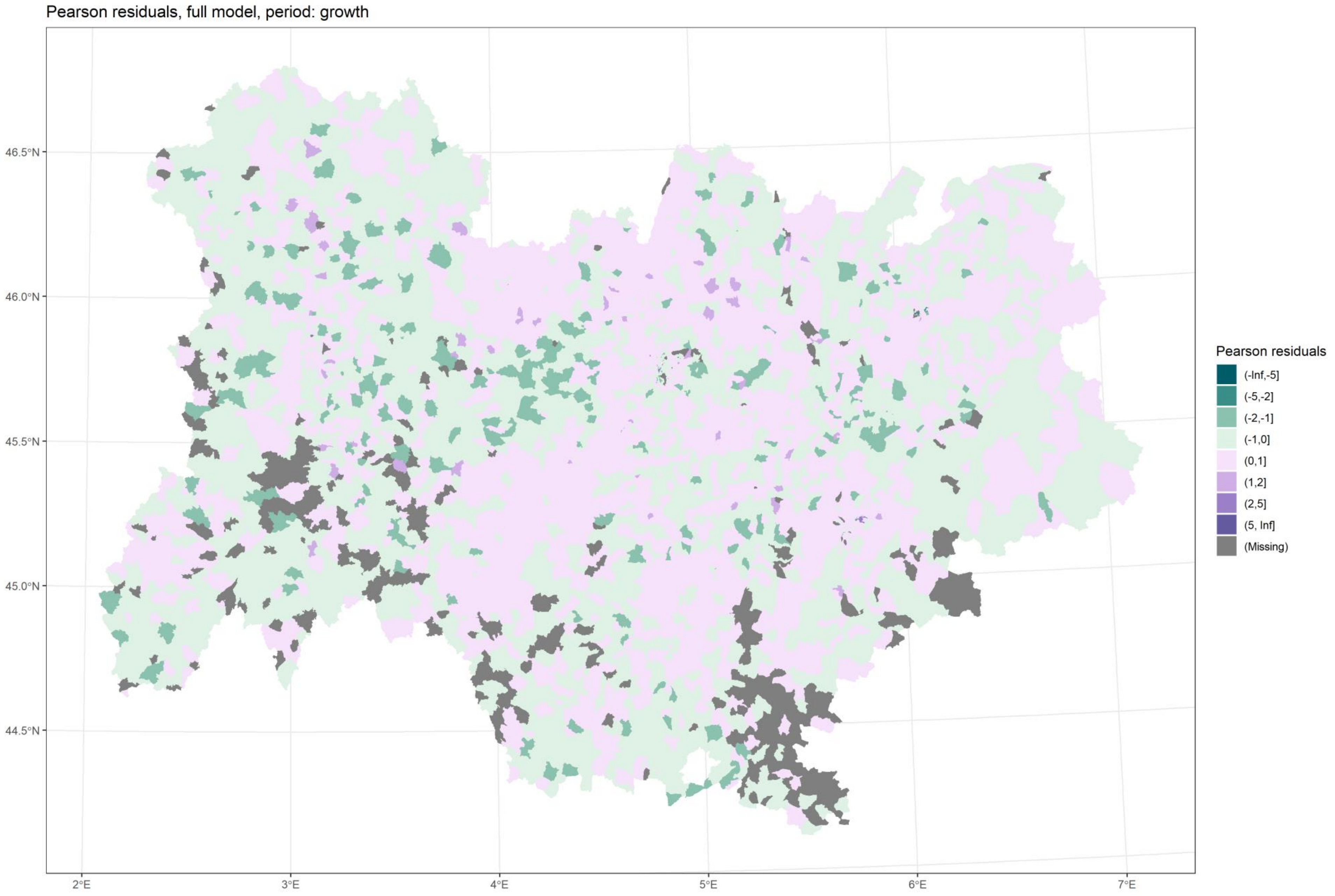

*Figure 12 Residuals, M4, growth*

Back to title

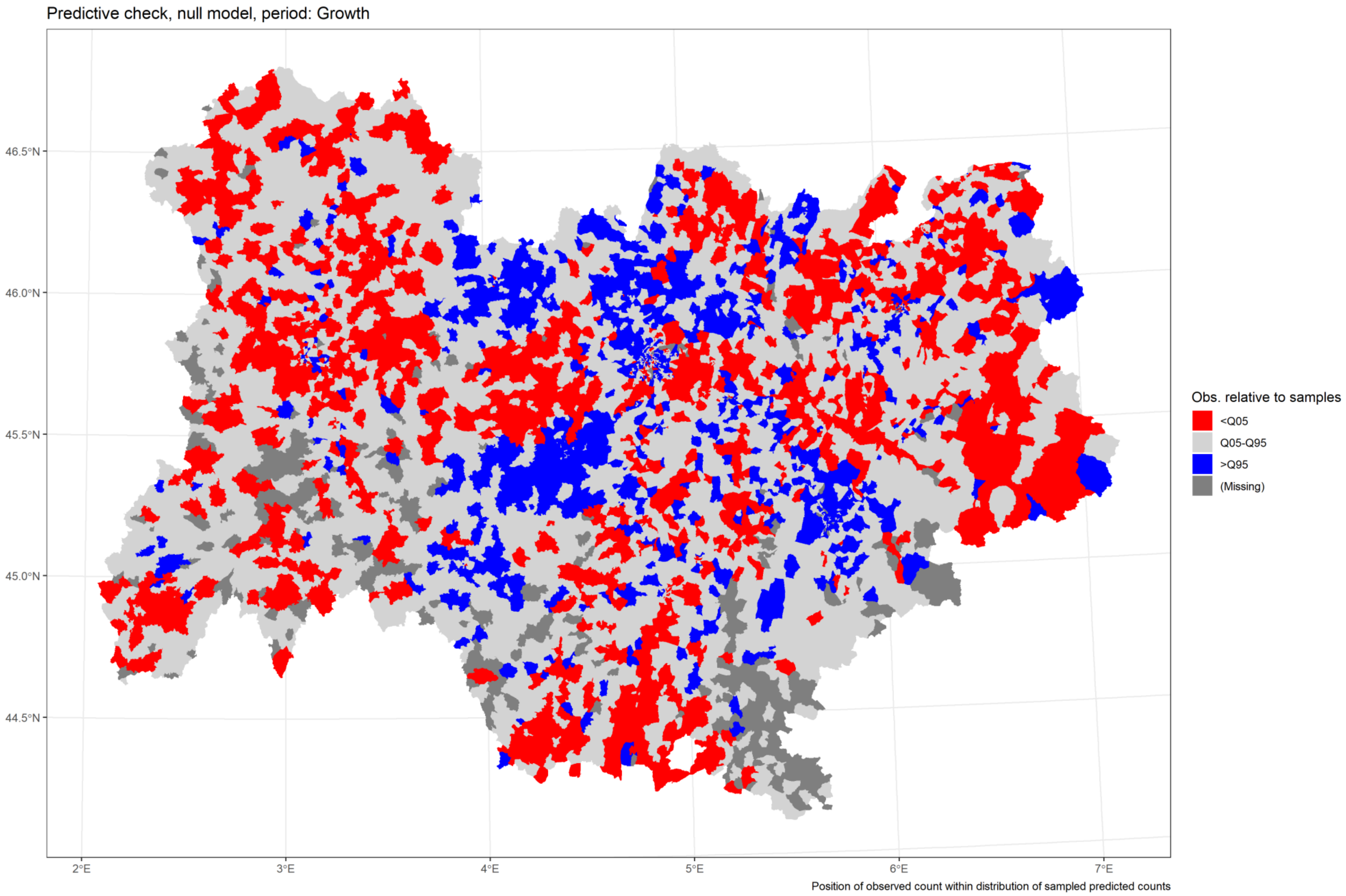

*Figure 13 Predictive check, M1, growth*

Back to title

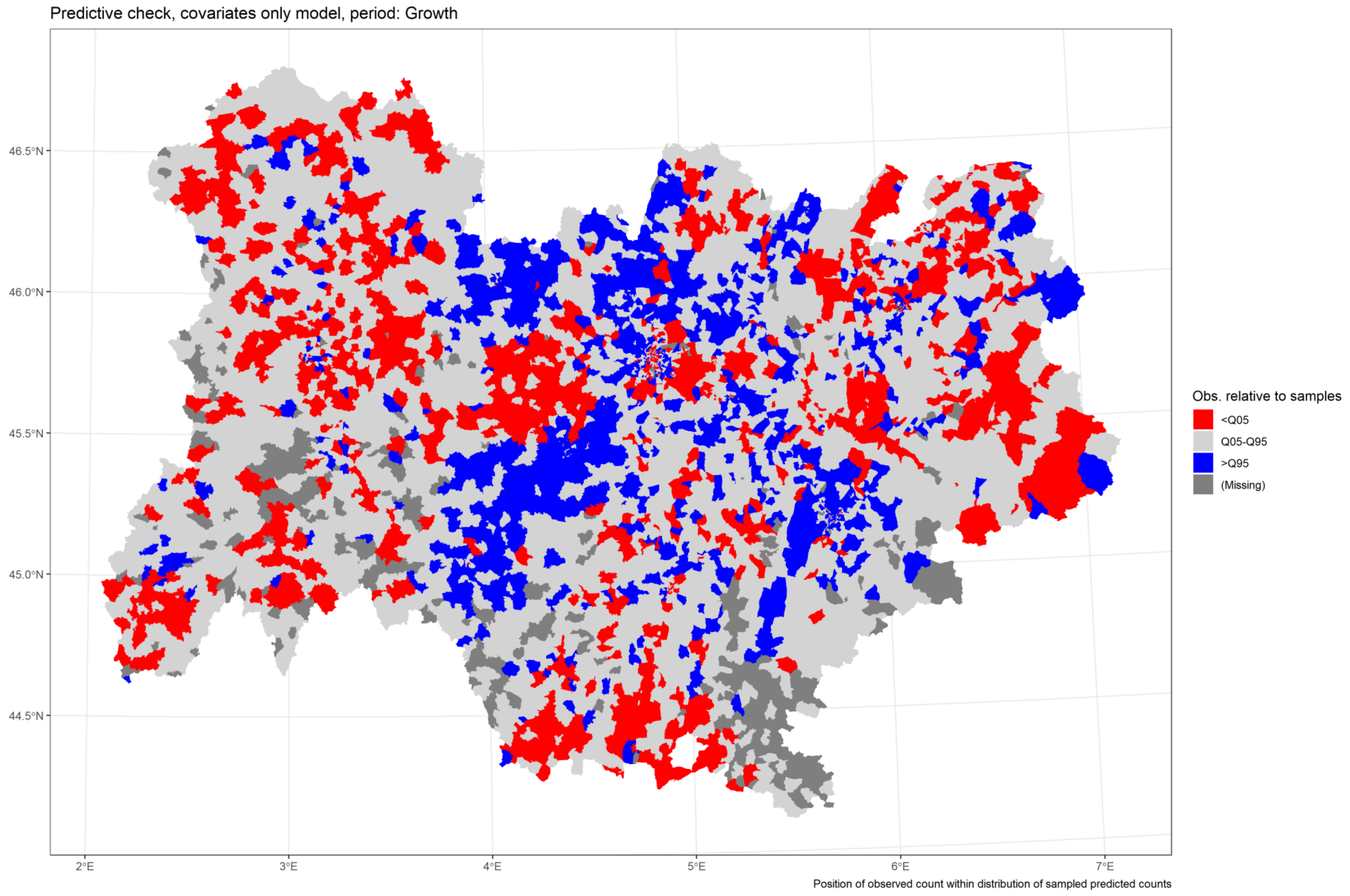

*Figure 14 Predictive check, M2, growth*

Back to title

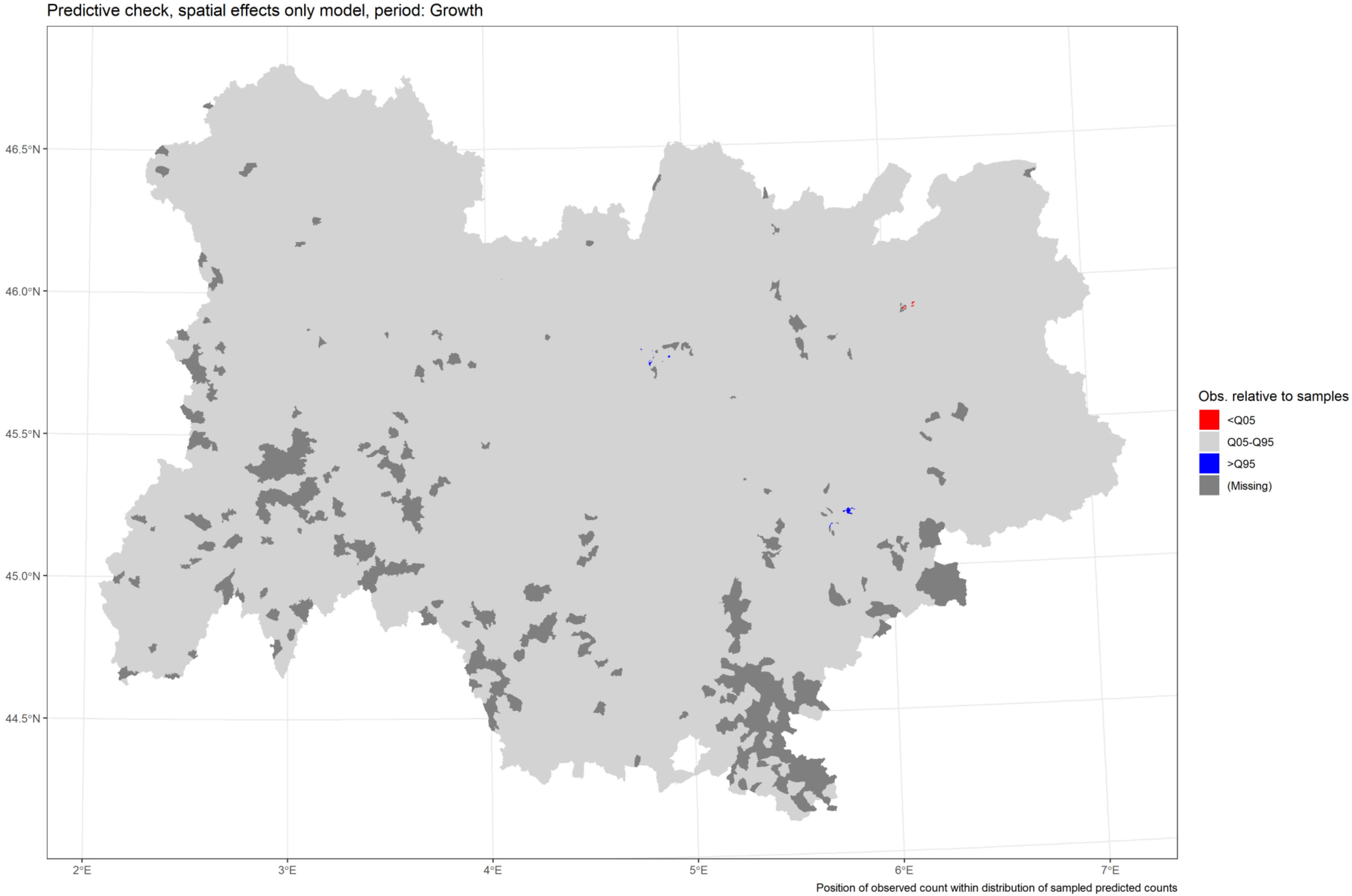

*Figure 15 Predictive check, M3, growth*

[Back to title](#)

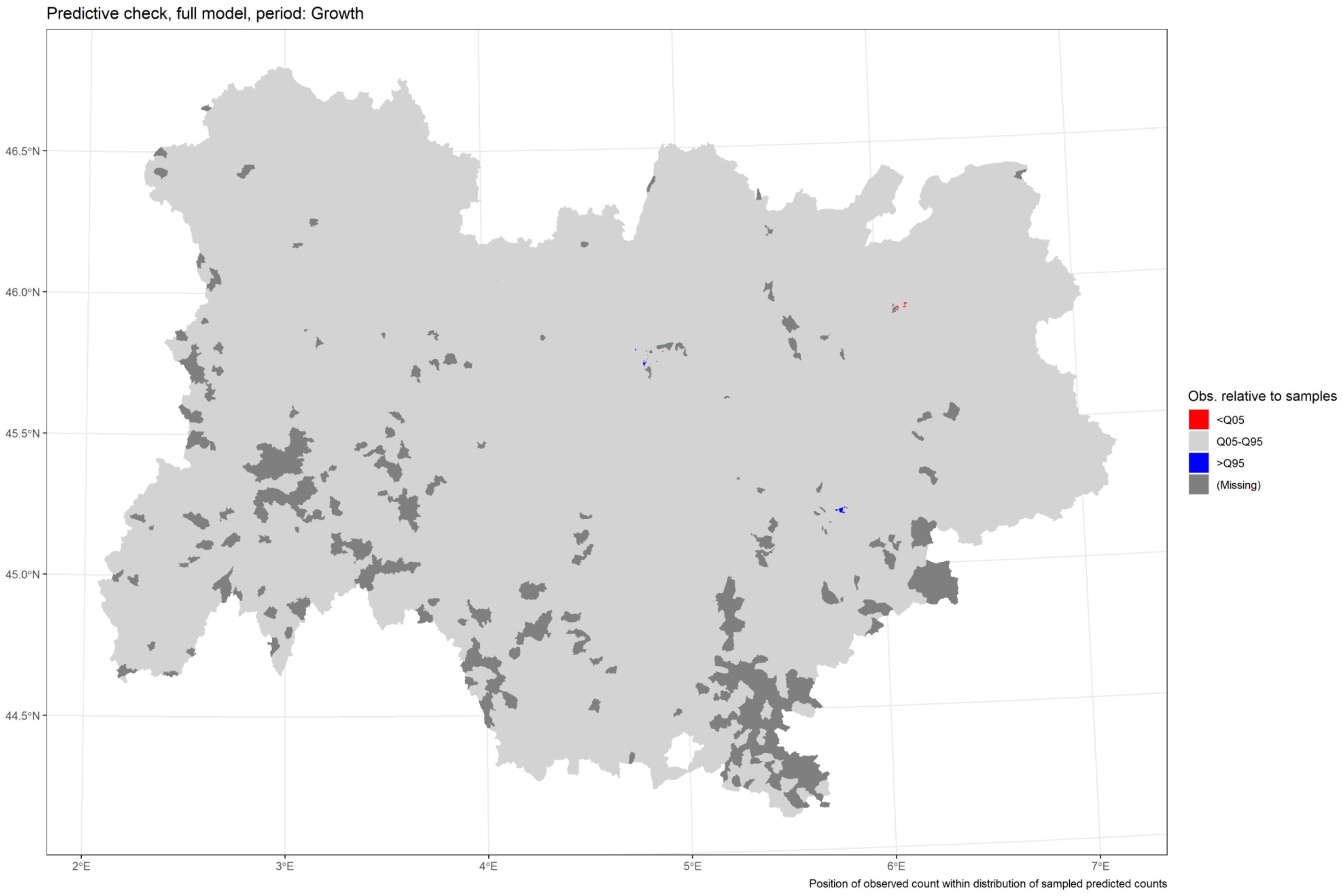

*Figure 16 Predictive check, M4, growth*

[Back to title](#)

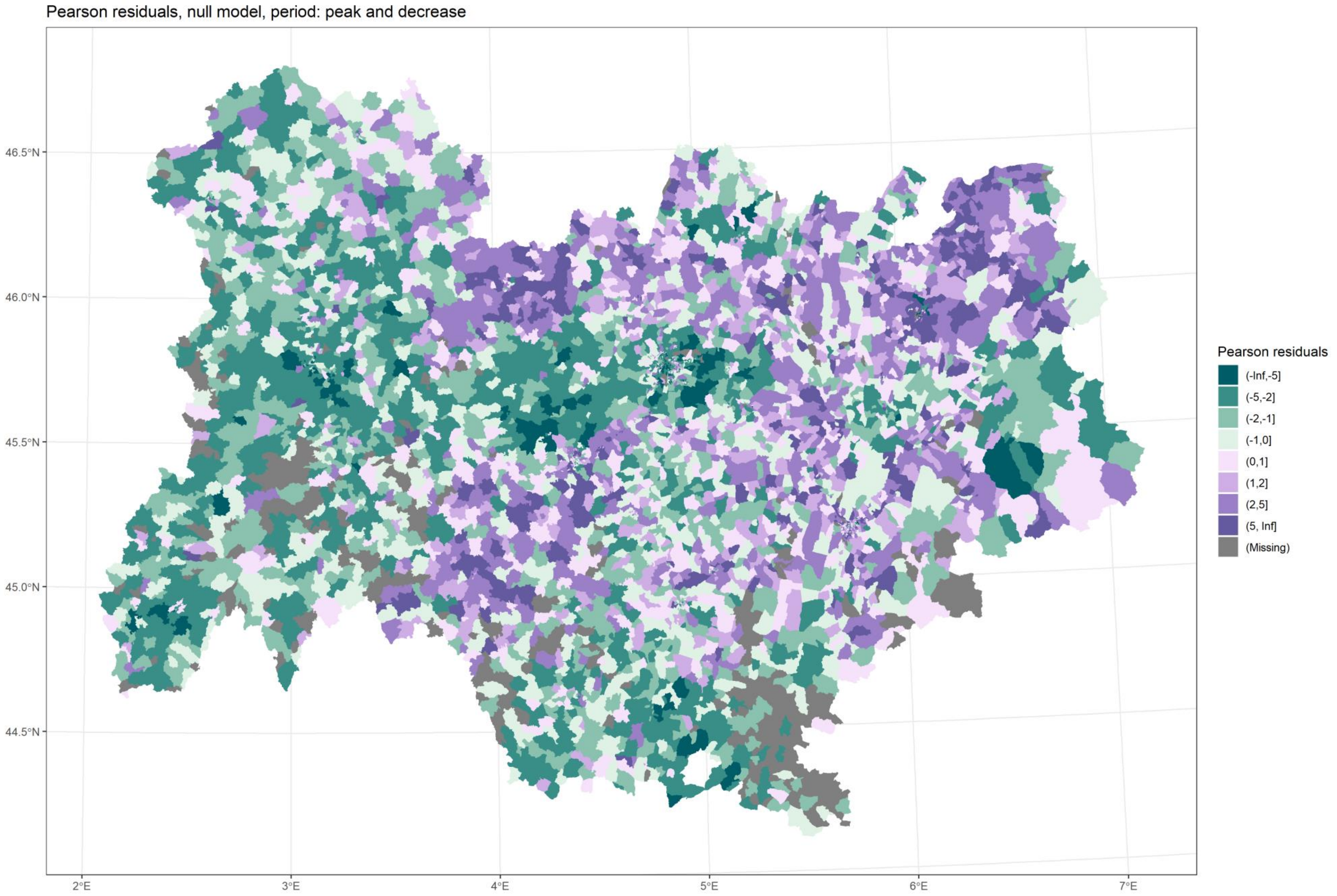

*Figure 17 Residuals, M1, peak and decrease*

[Back to title](#)

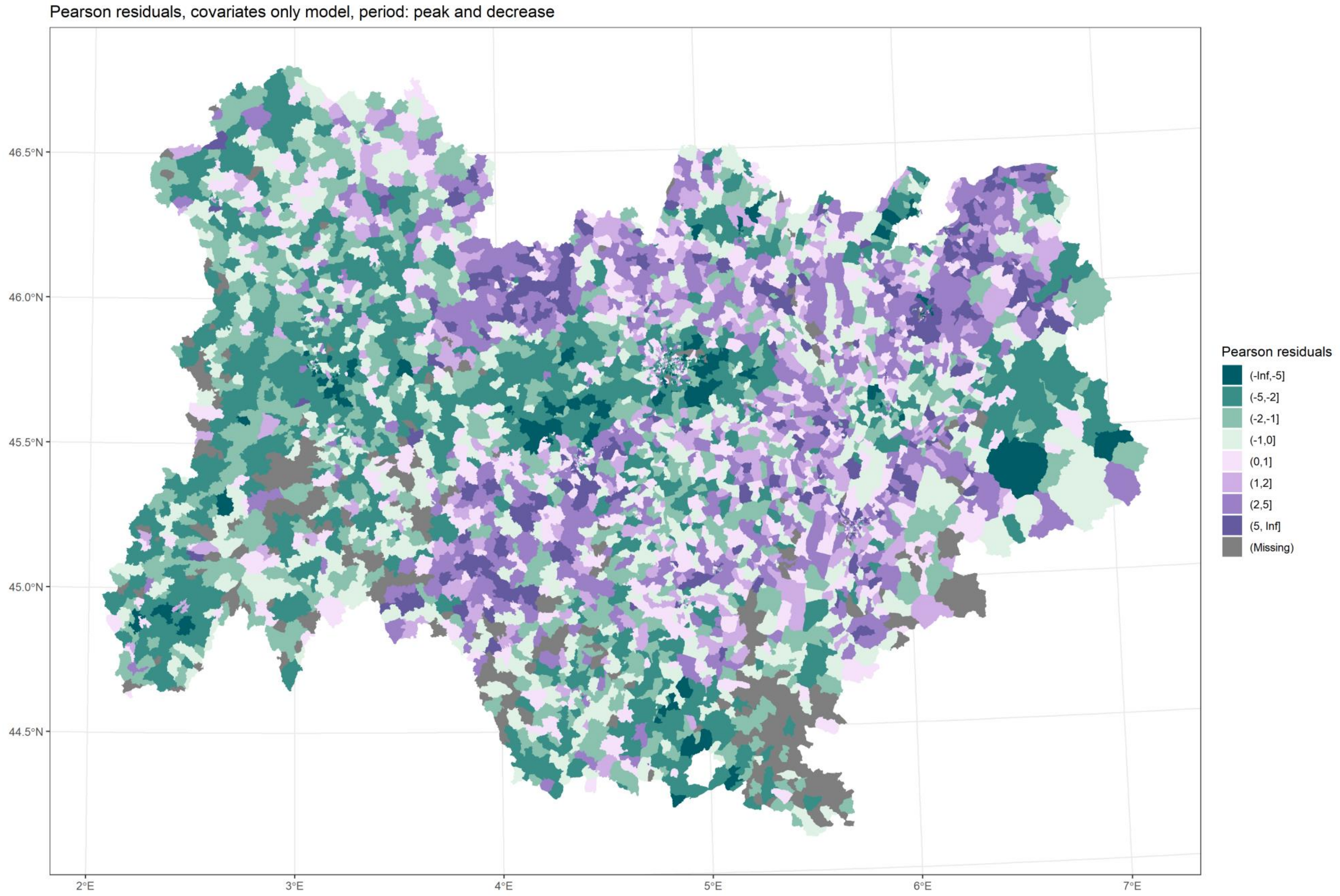

*Figure 18 Residuals, M2, peak and decrease*

Back to title

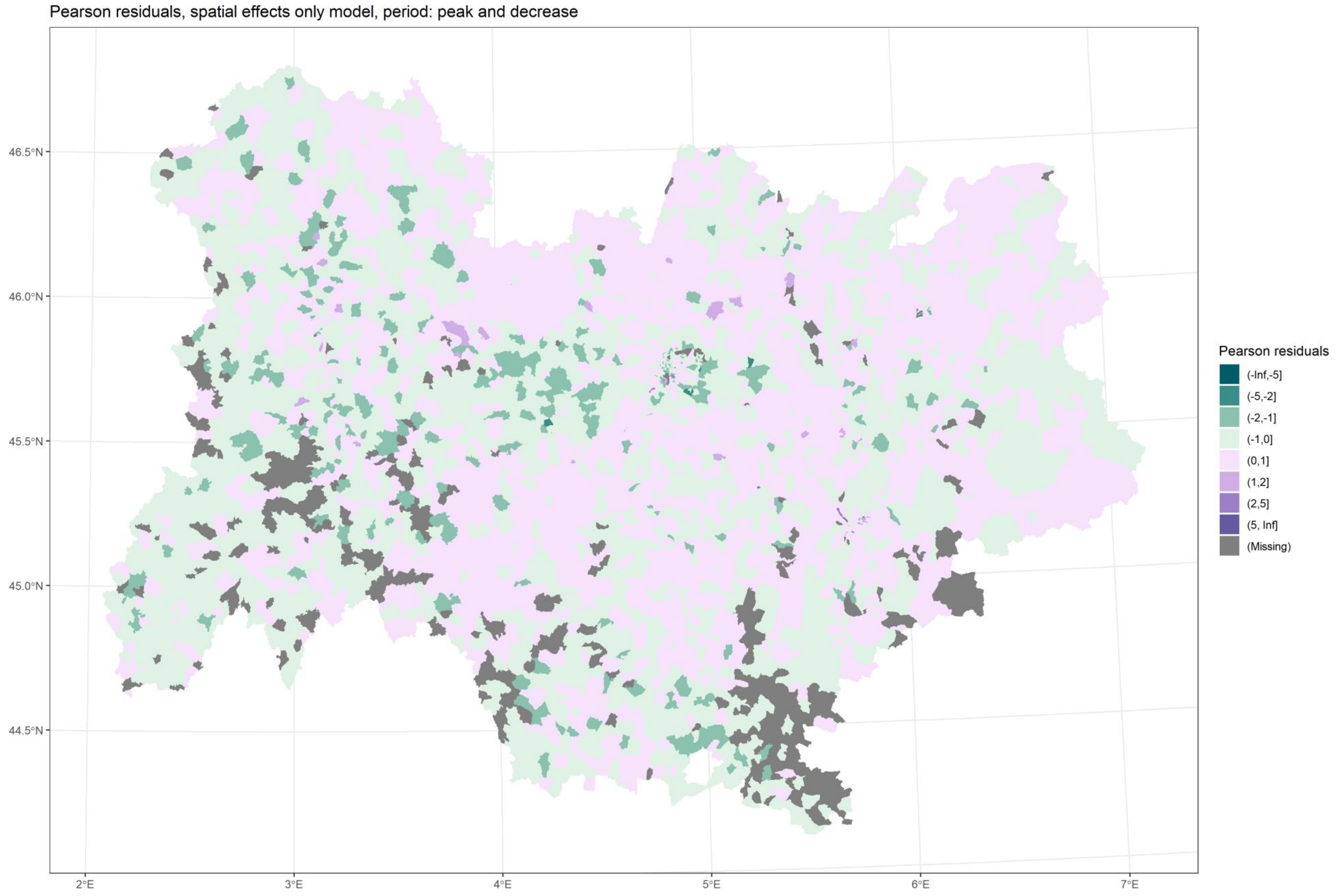

*Figure 19 Residuals, M3, peak and decrease*

Back to title

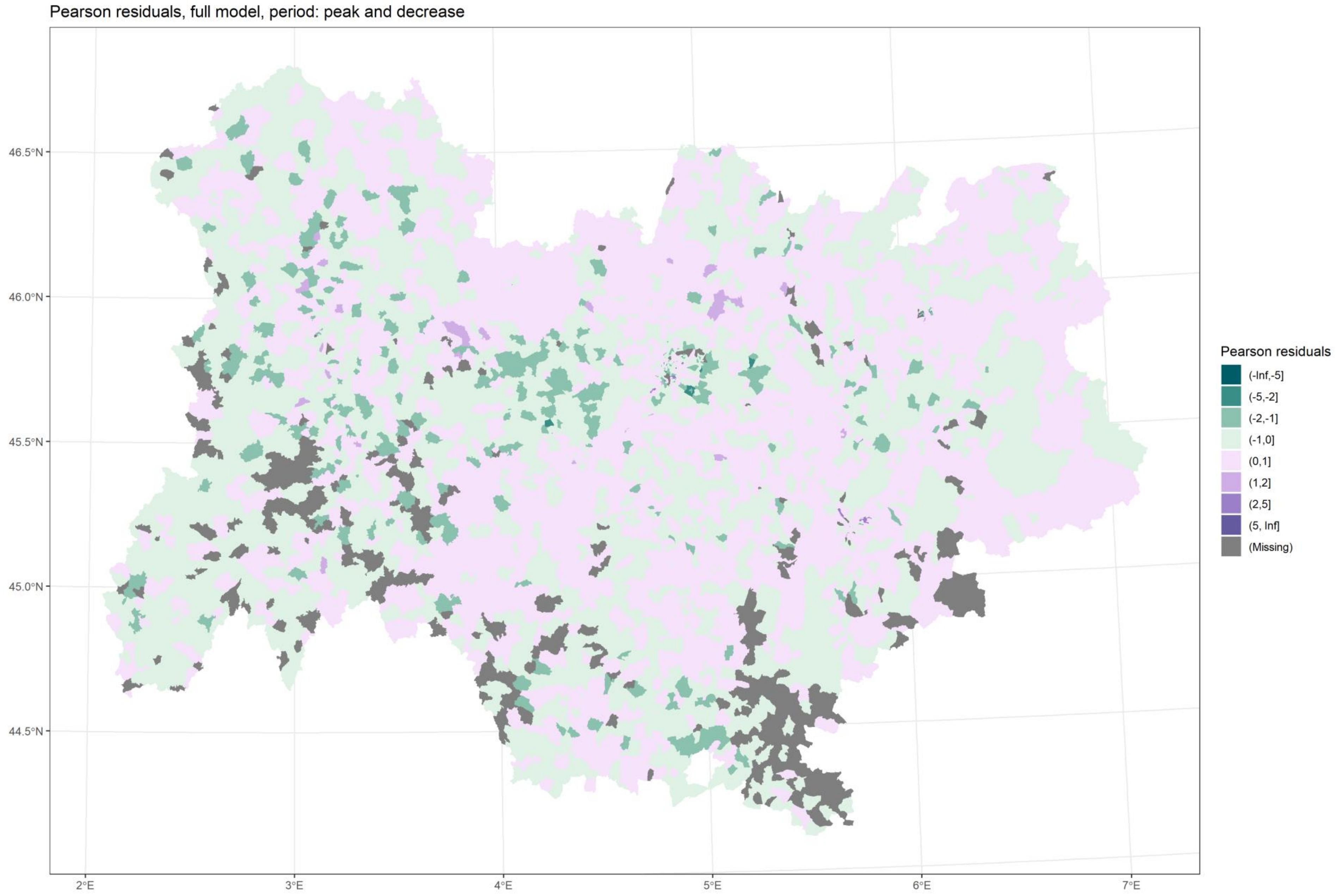

*Figure 20 Residuals, M4, peak and decrease*

Back to title

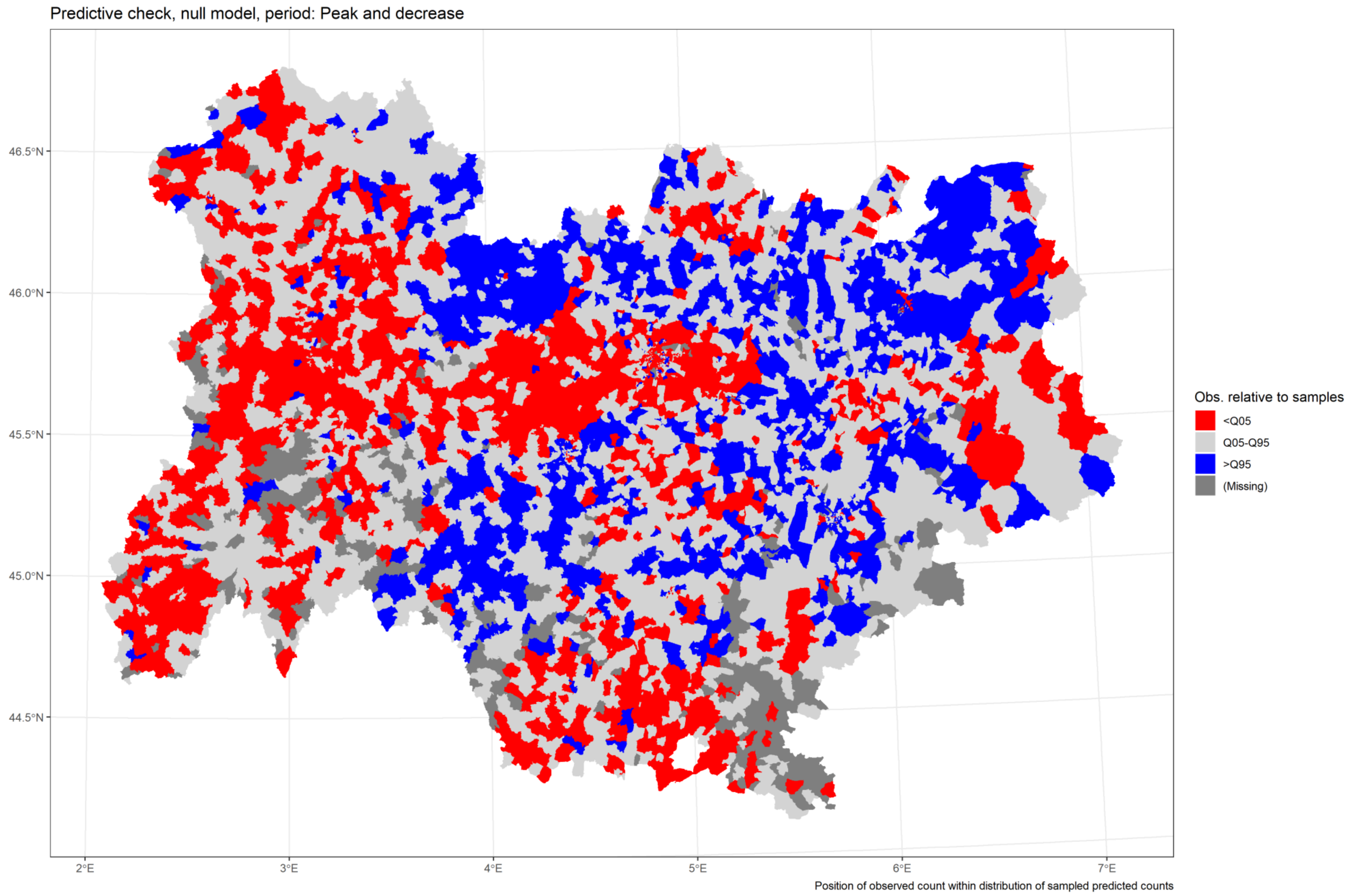

*Figure 21 Predictive check, M1, peak and decrease*

Back to title

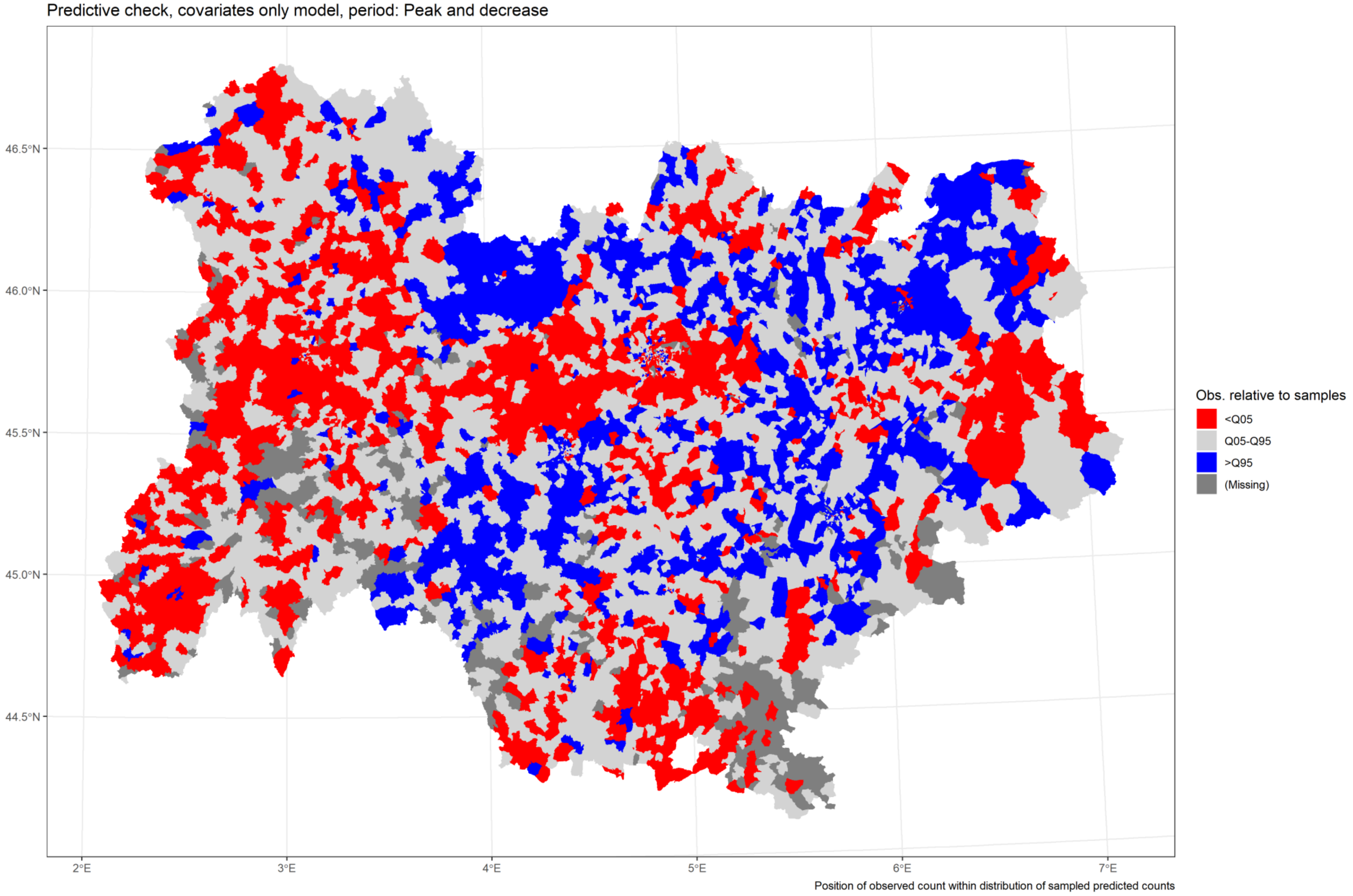

*Figure 22 Predictive check, M2, peak and decrease*

Back to title

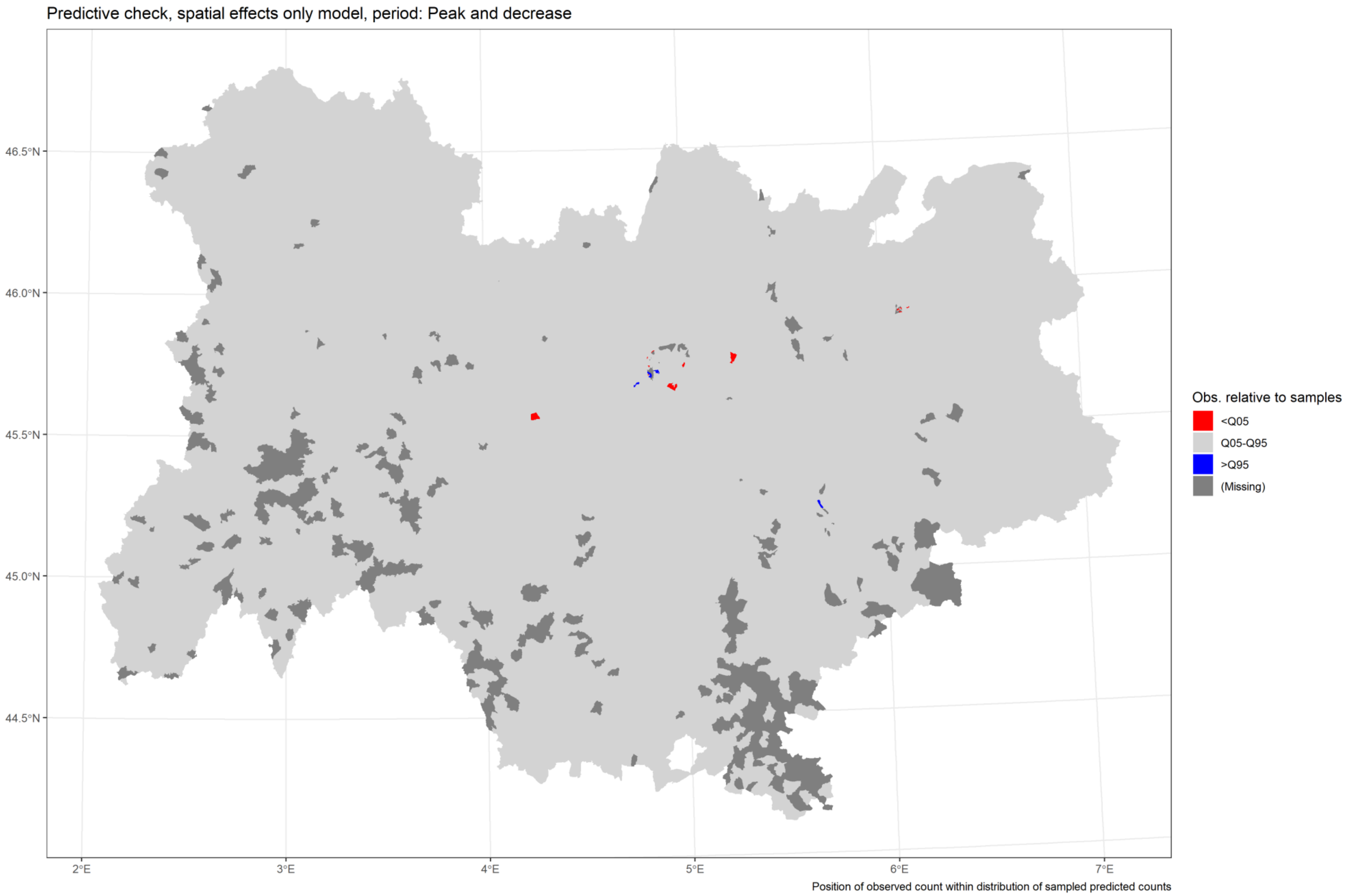

*Figure 23 Predictive check, M3, peak and decrease*

Back to title

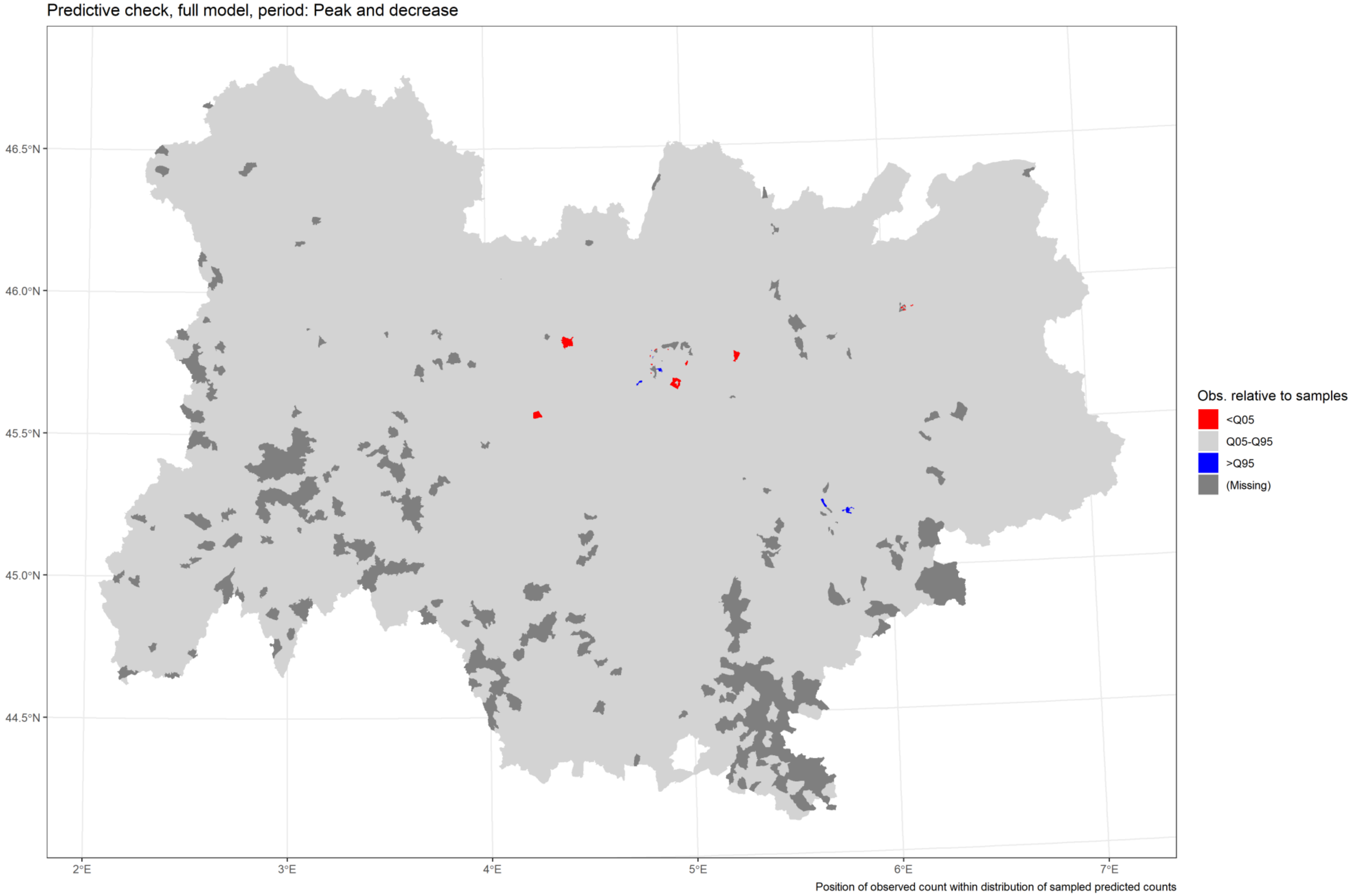

*Figure 24 Predictive check, M4, peak and decrease*

Back to title

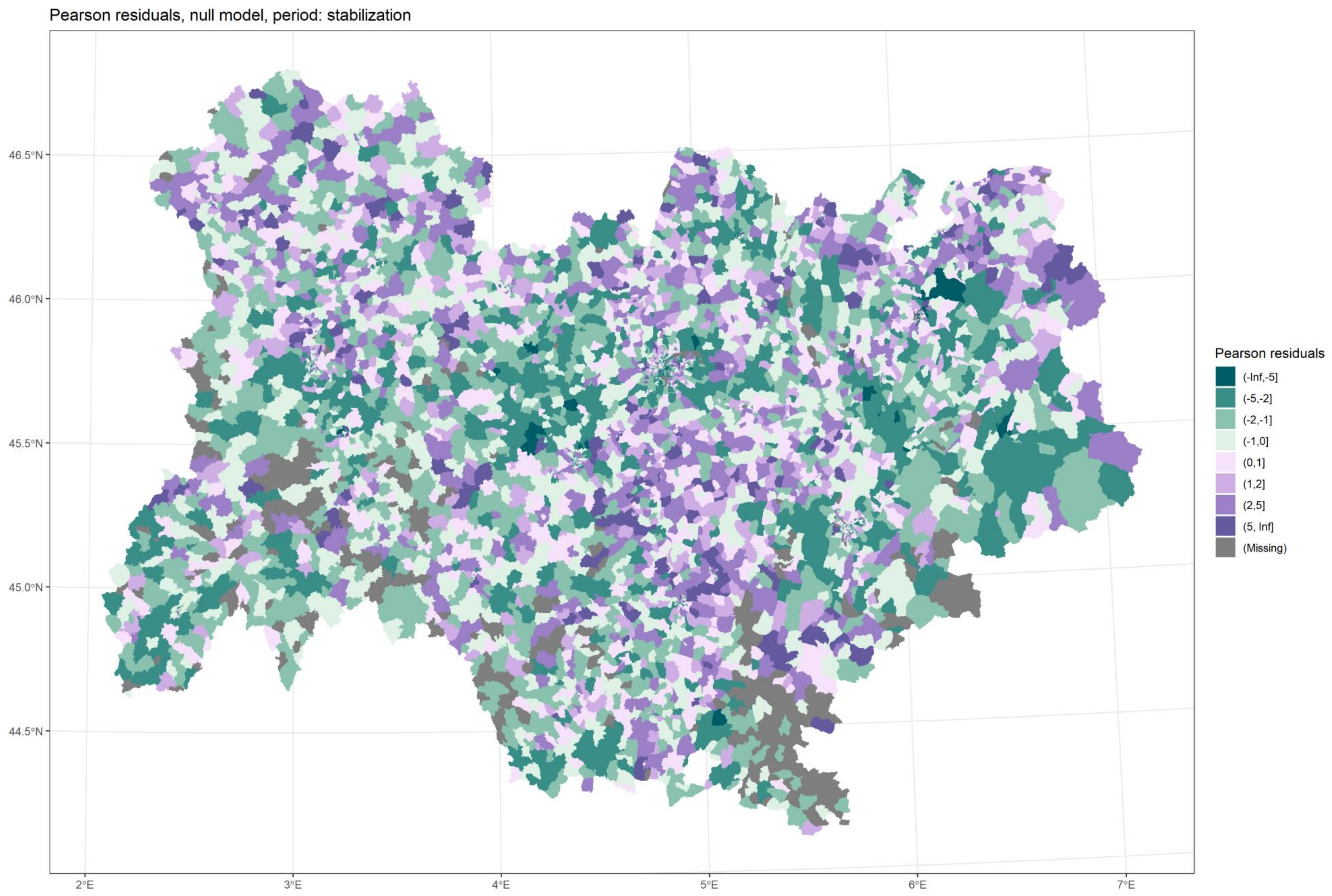

*Figure 25 Residuals, M1, stabilization*

Back to title

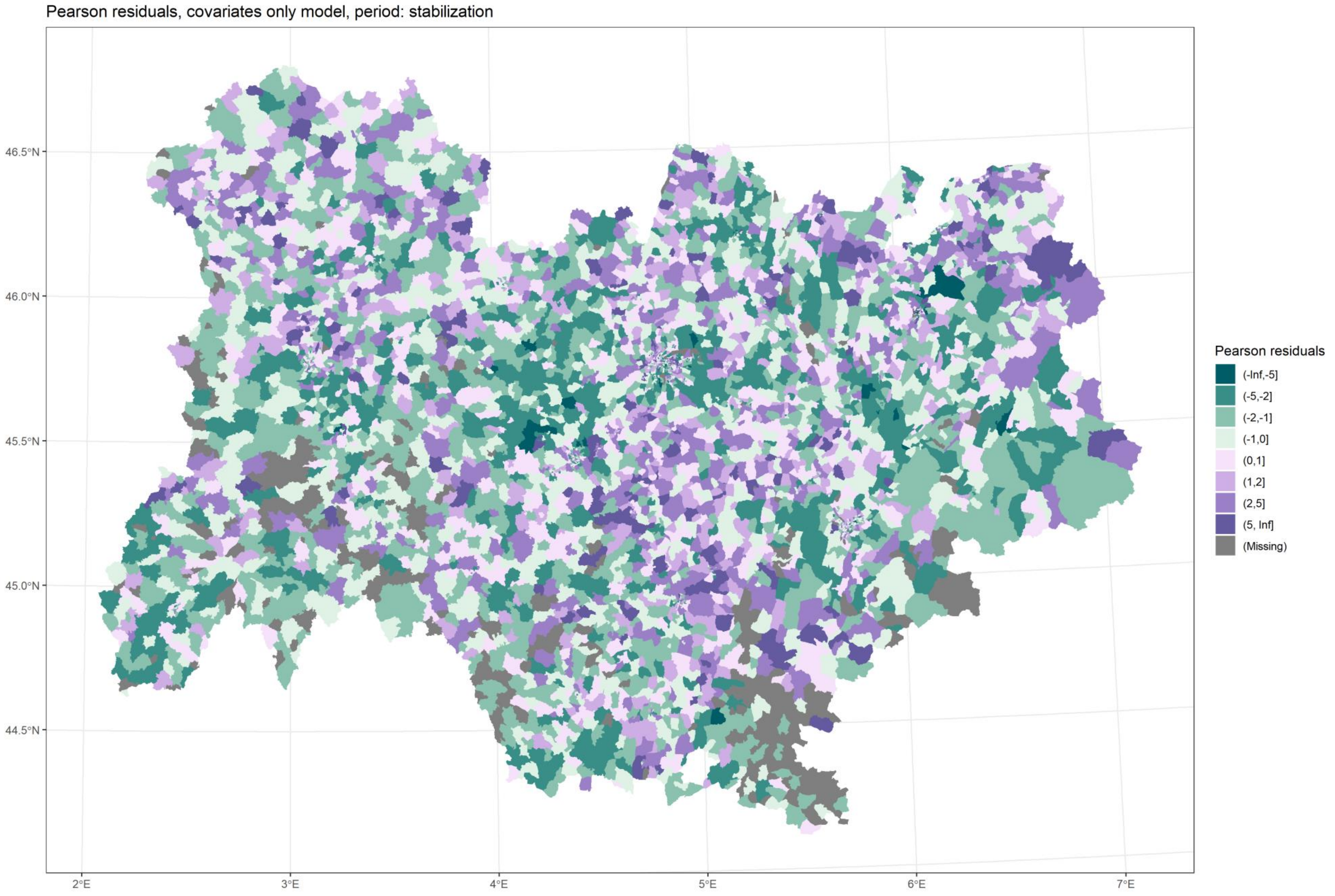

*Figure 26 Residuals, M2, stabilization*

Back to title

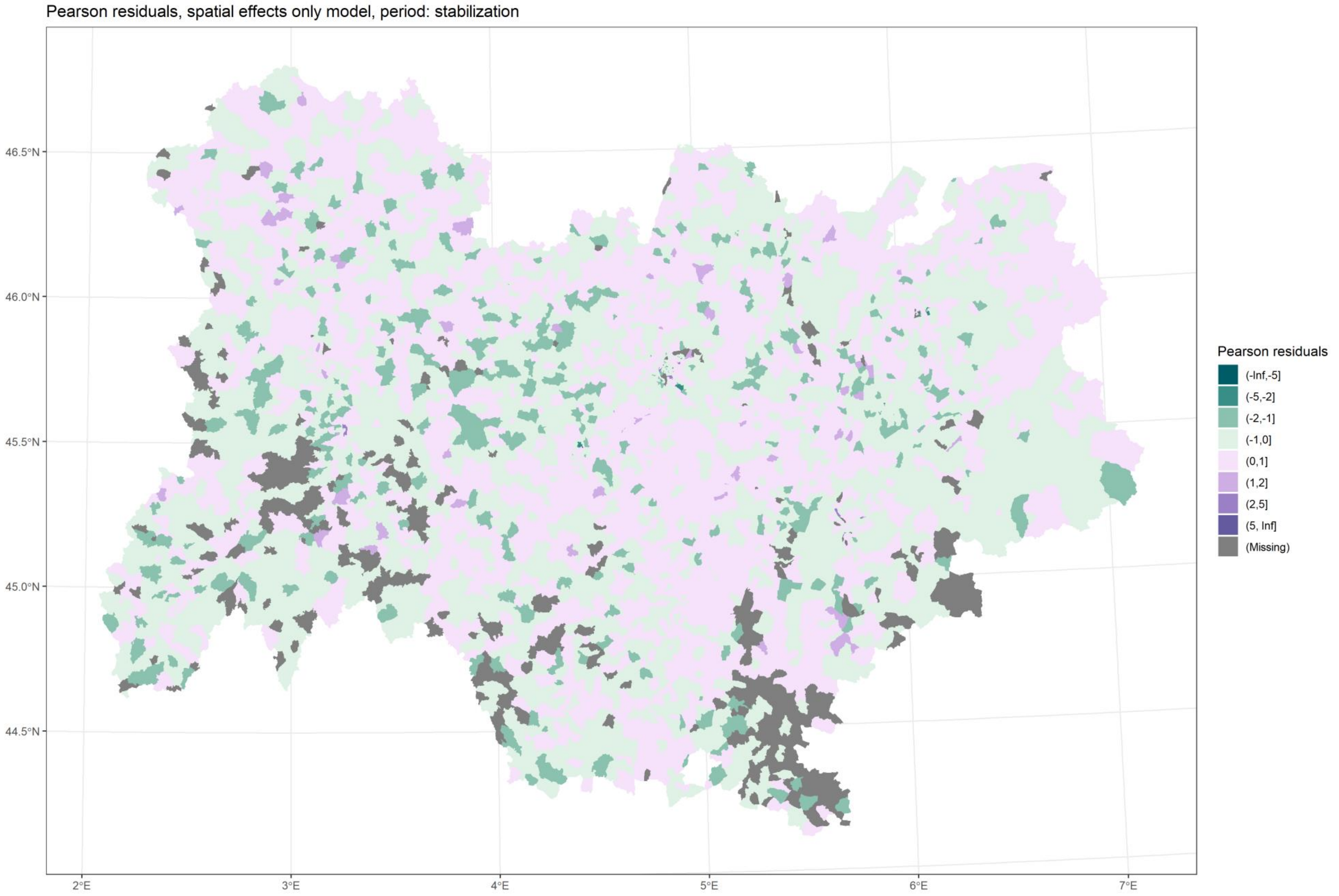

*Figure 27 Residuals, M3, stabilization*

Back to title

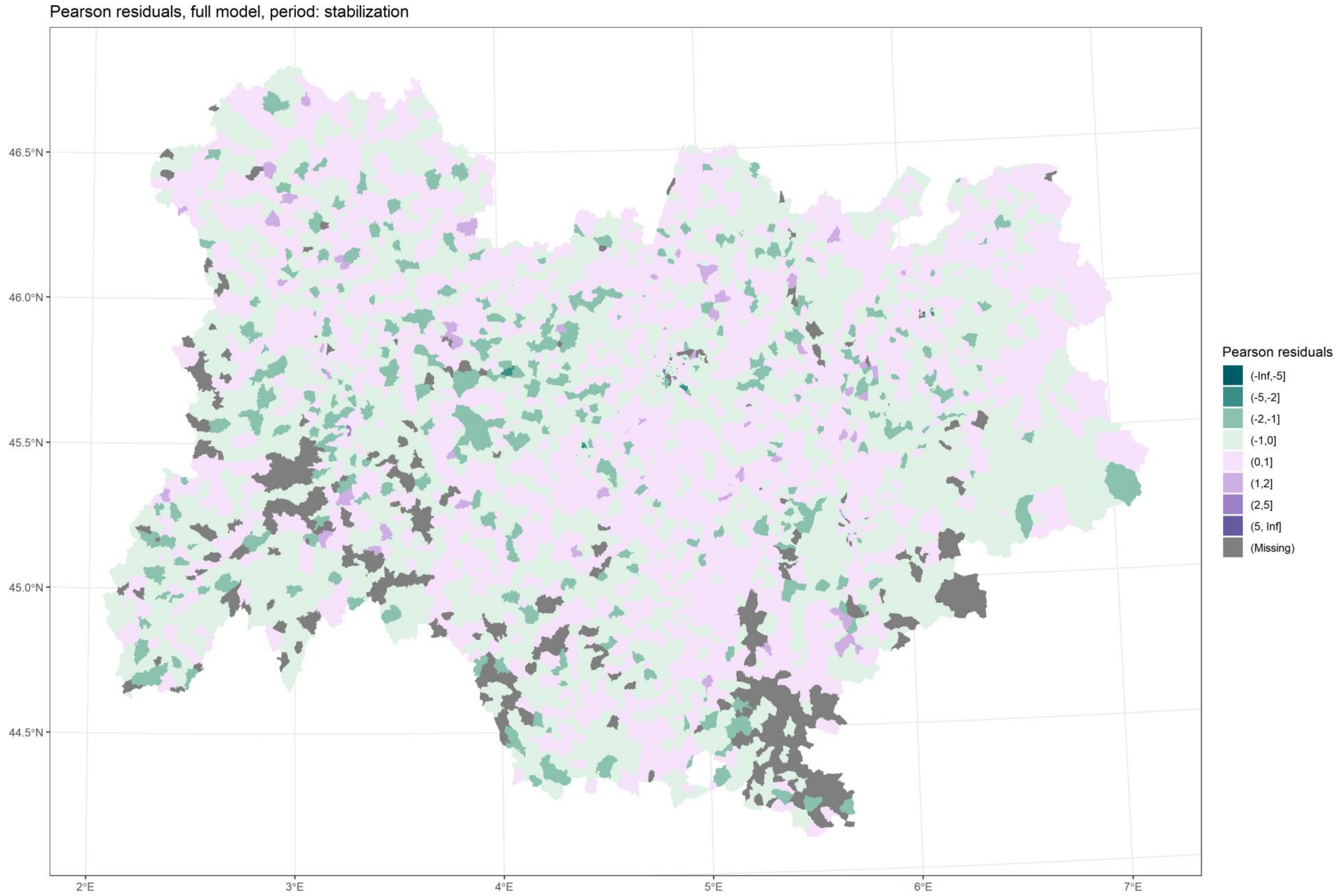

*Figure 28 Residuals, M4, stabilization*

Back to title

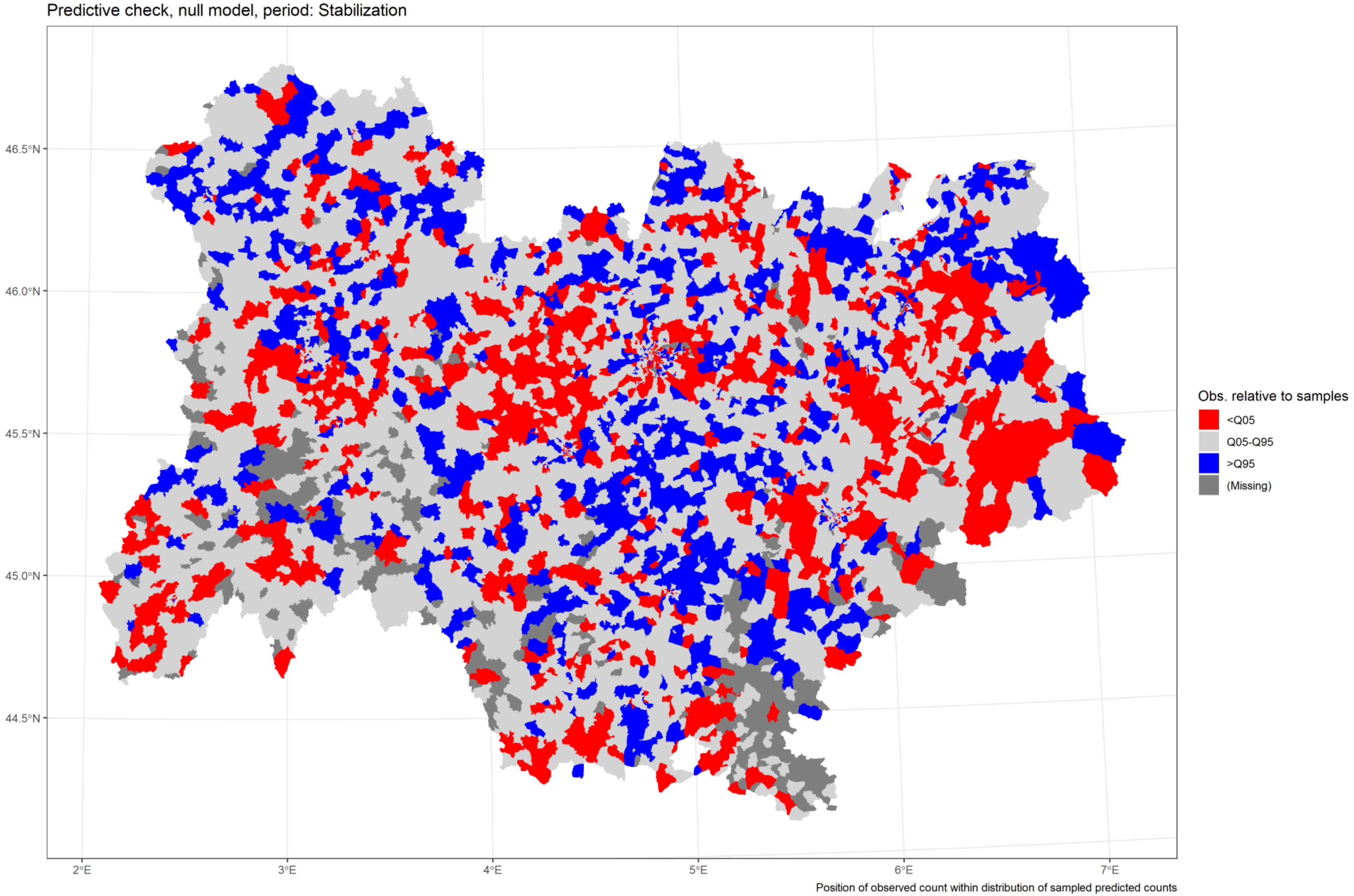

*Figure 29 Predictive check, M1, stabilization*

Back to title

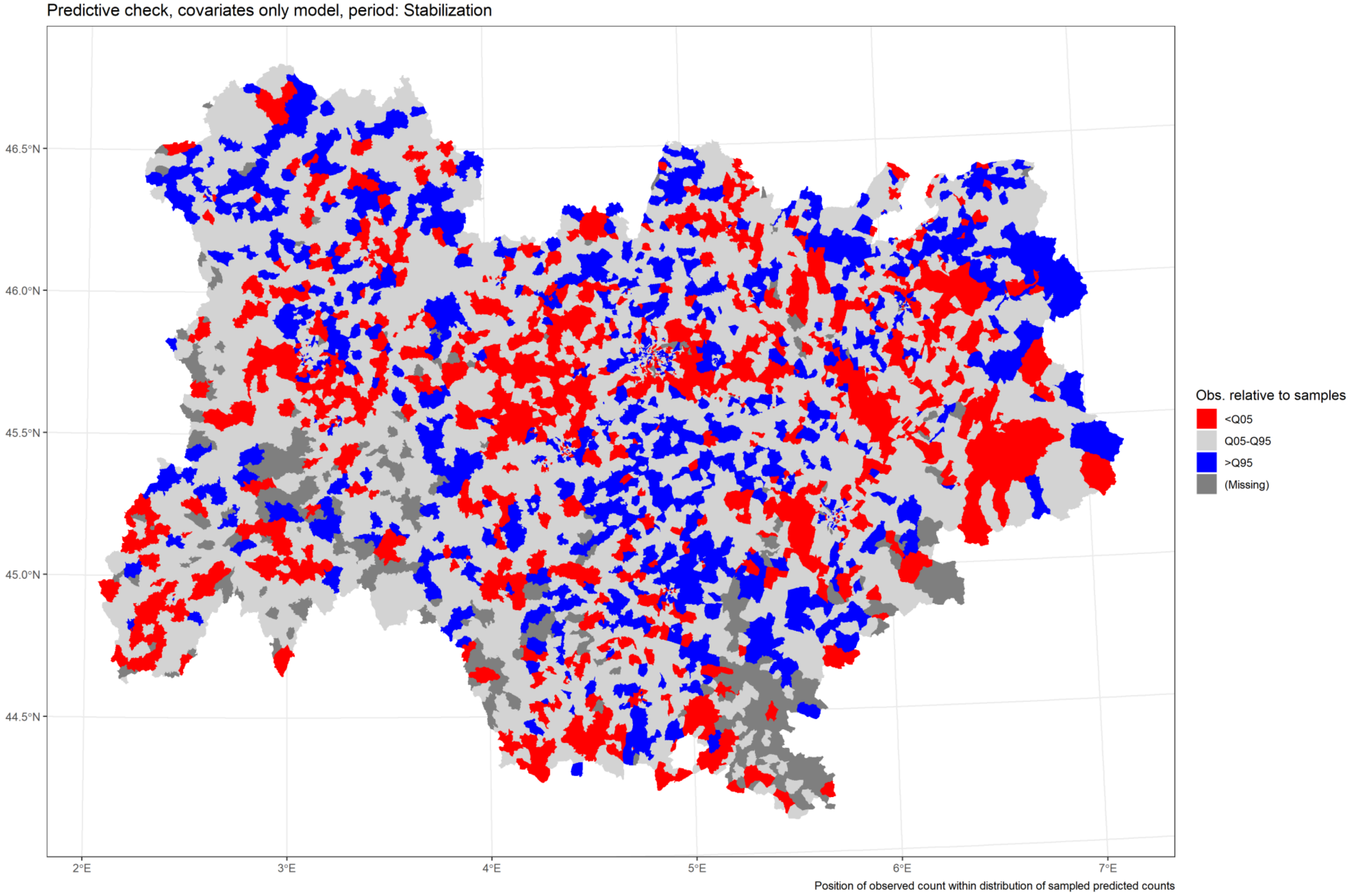

*Figure 30 Predictive check, M2, stabilization*

[Back to title](#)

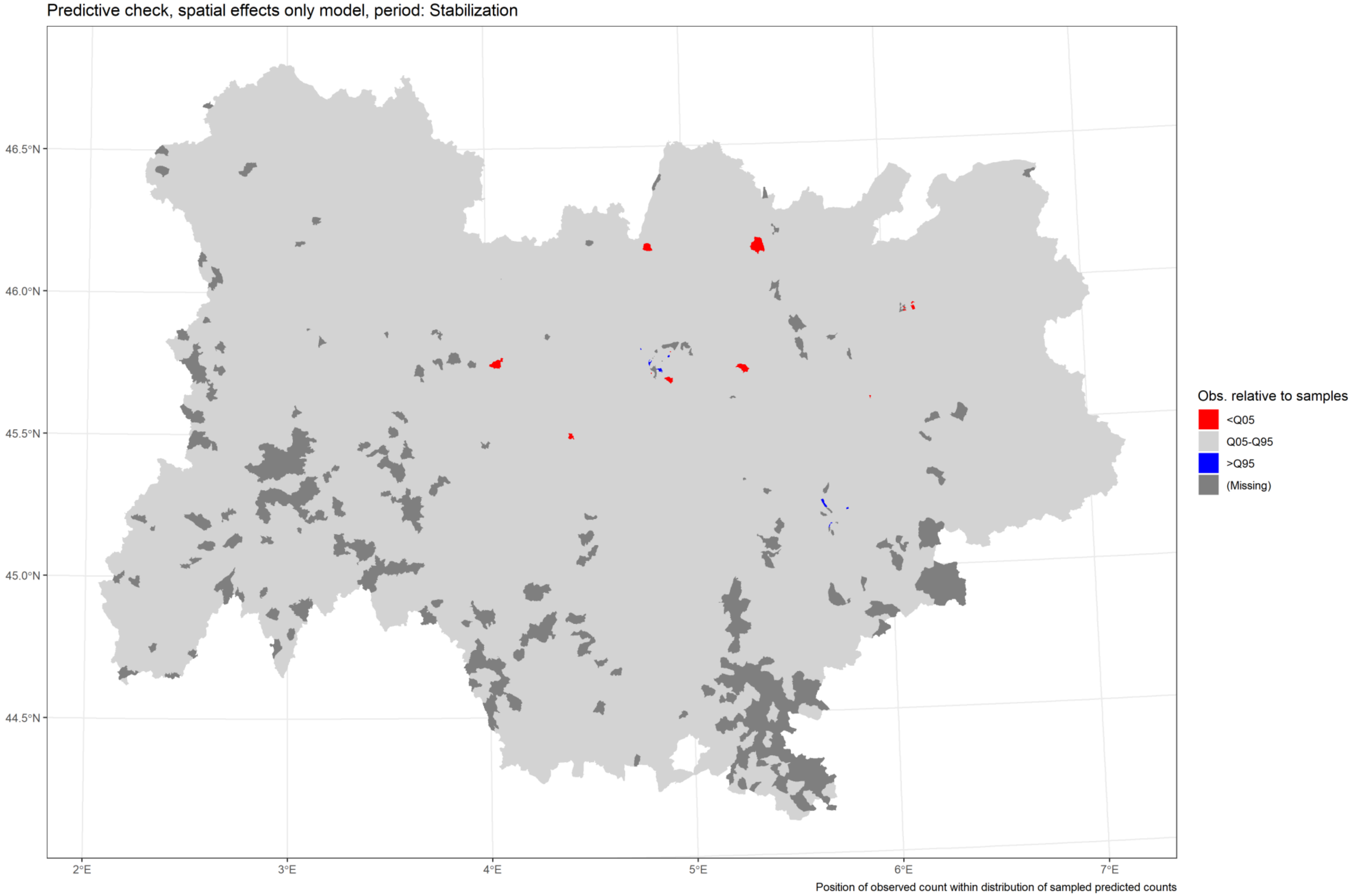

*Figure 31 Predictive check, M3, stabilization*

Back to title

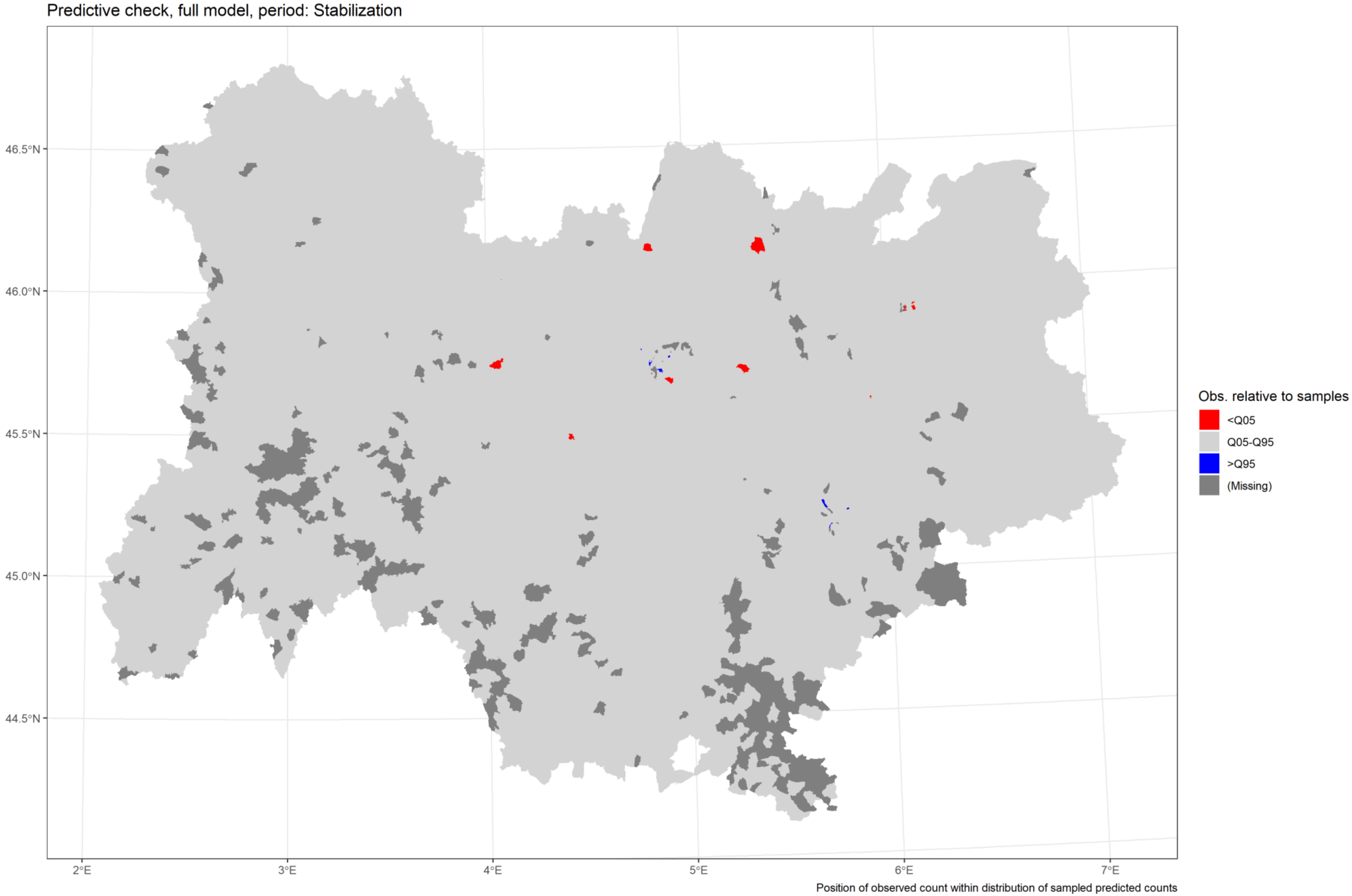

*Figure 32 Predictive check, M4, stabilization*

Back to title
