## Supplementary material for "Usefulness of ecological mobility and socio-economic indicators in SARS-CoV-2 infection modelling: a French case study": S4 - Metropolitan areas maps.pdf

### Predicted incidence rates in metropolitan areas

The following maps are focuses on the fourth metropolitan areas within the study region: Clermont-Ferrand, Grenoble, Lyon, Saint-Etienne. These a zoomed in versions of the Figure 1 as displayed in the main article.

Figure 1 Study region map (in the main article) .......... 3
Figure 2 Clermont-Ferrand (west) .......... 4
Figure 3 Grenoble (southeast) .......... 5
Figure 4 Lyon (regional capital, centre-east) .......... 6
Figure 5 Saint-Etienne (southwest of Lyon) .......... 7

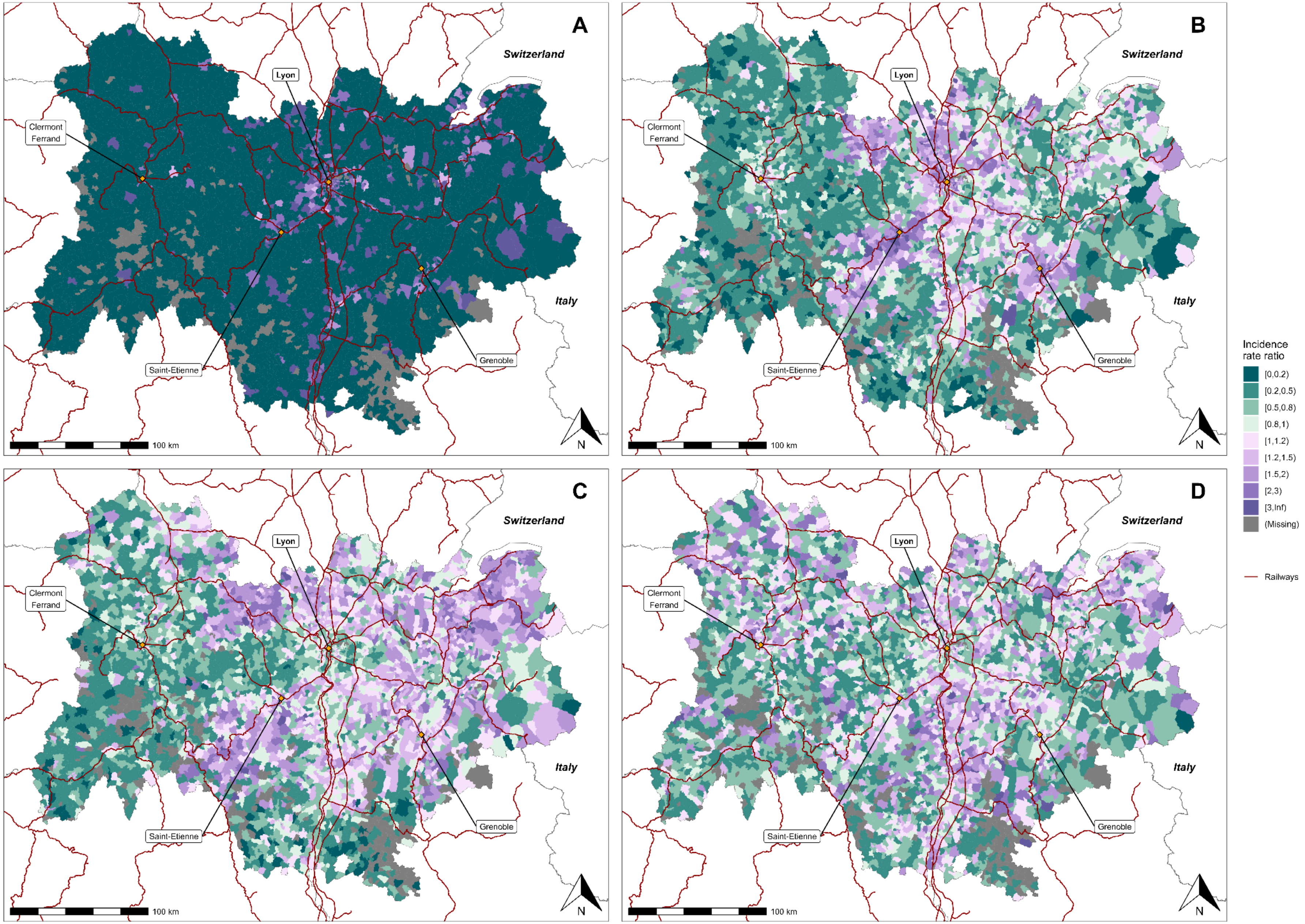


*Figure 1 predicted incidence rate ratio of SARS-CoV-2 infection for the IRIS (infra-municipal spatial unit) of the French region Auvergne-Rhône-Alpes during the second epidemic wave: model with covariates and spatial effect (M4). From A to D: Low incidence, Growth, Peak and decrease, and Stabilization periods (in the main article)*

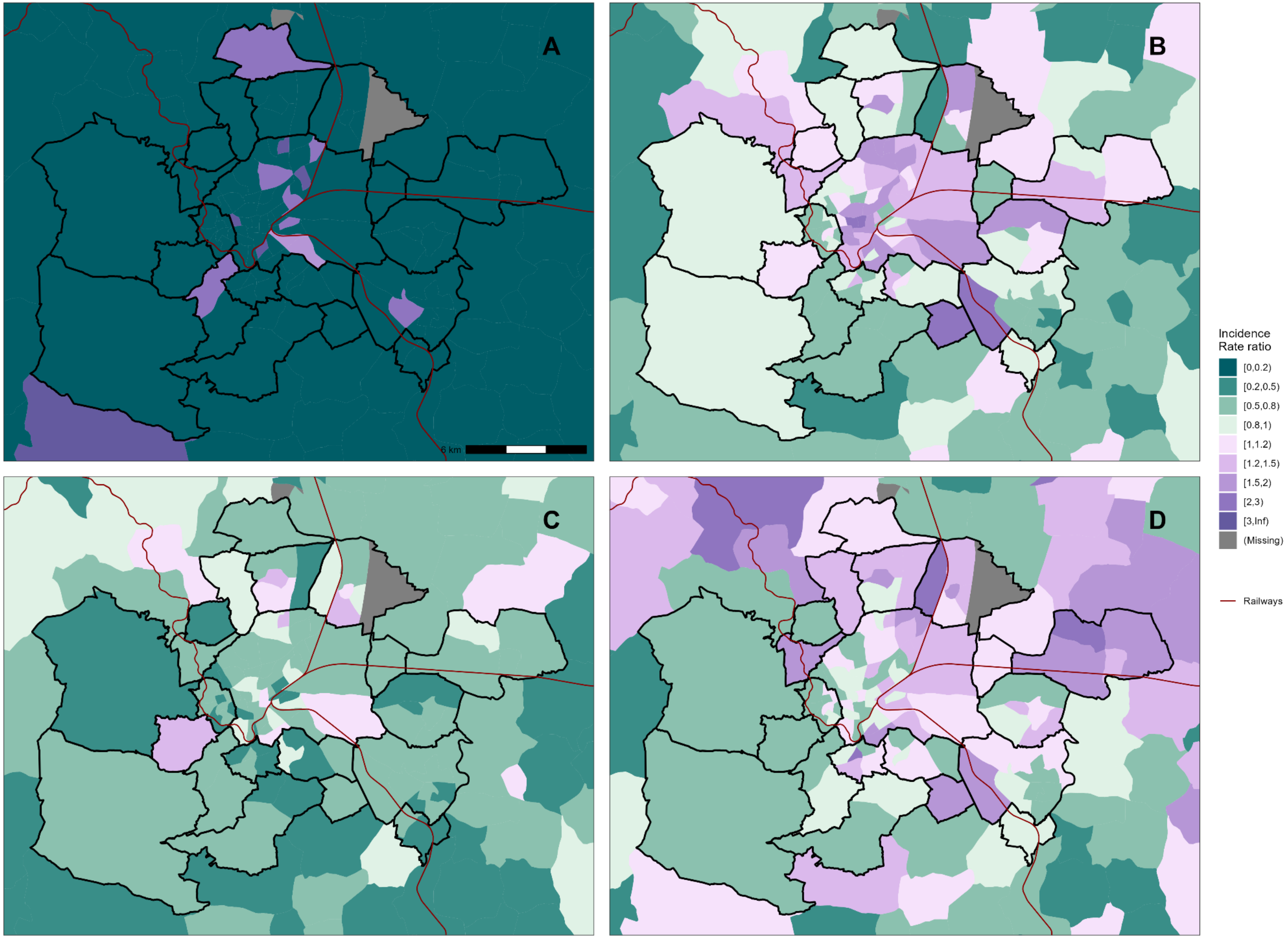


*Figure 2 predicted incidence rate ratio of SARS-CoV-2 infection for the IRIS (infra-municipal spatial unit) of the metropolitan area of Clermont-Ferrand during the second epidemic wave: model with covariates and spatial effect (M4). From A to D: Low incidence, Growth, Peak and decrease, and Stabilization periods*



Back to title

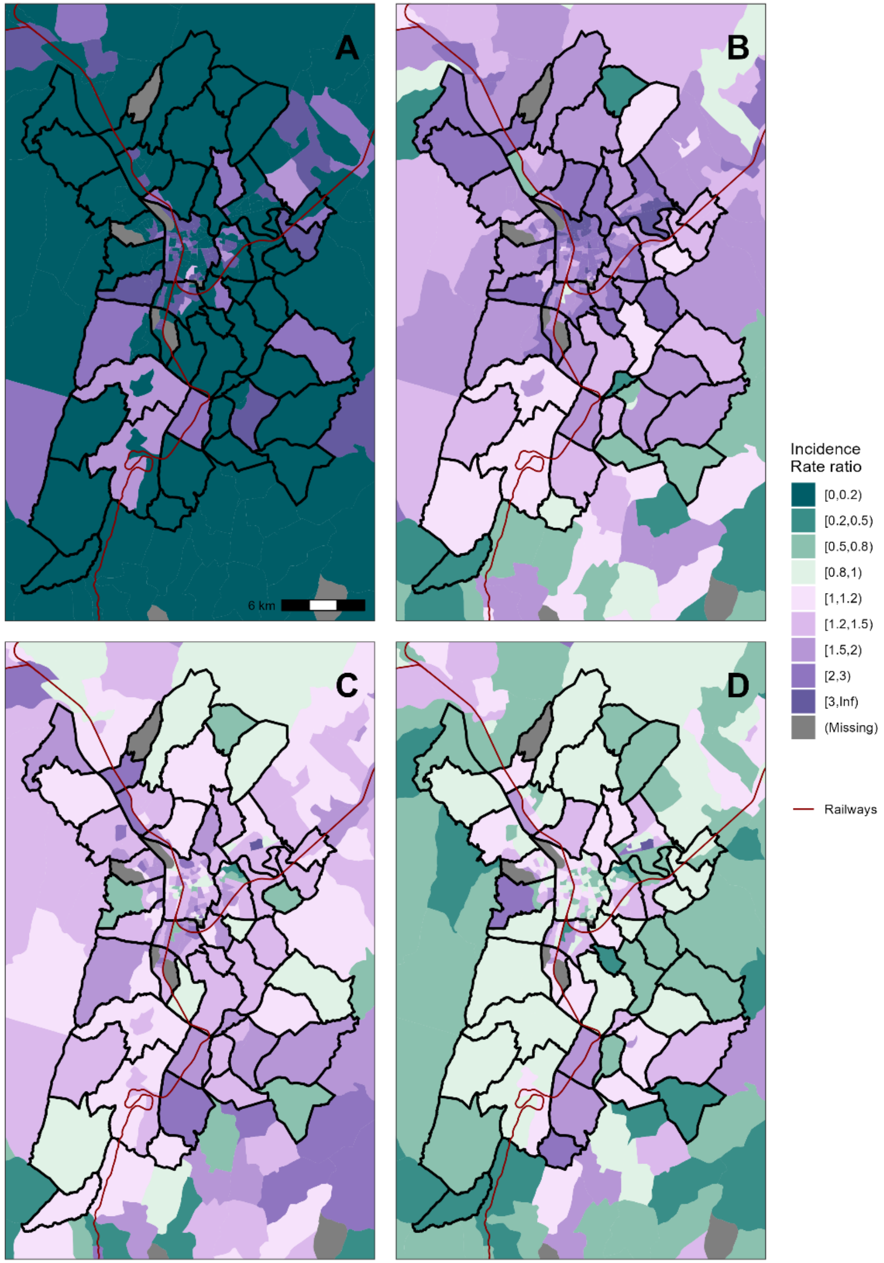


*Figure 3 predicted incidence rate ratio of SARS-CoV-2 infection for the IRIS (infra-municipal spatial unit) of the metropolitan area of Grenoble during the second epidemic wave: model with covariates and spatial effect (M4). From A to D: Low incidence, Growth, Peak and decrease, and Stabilization periods*



Back to title

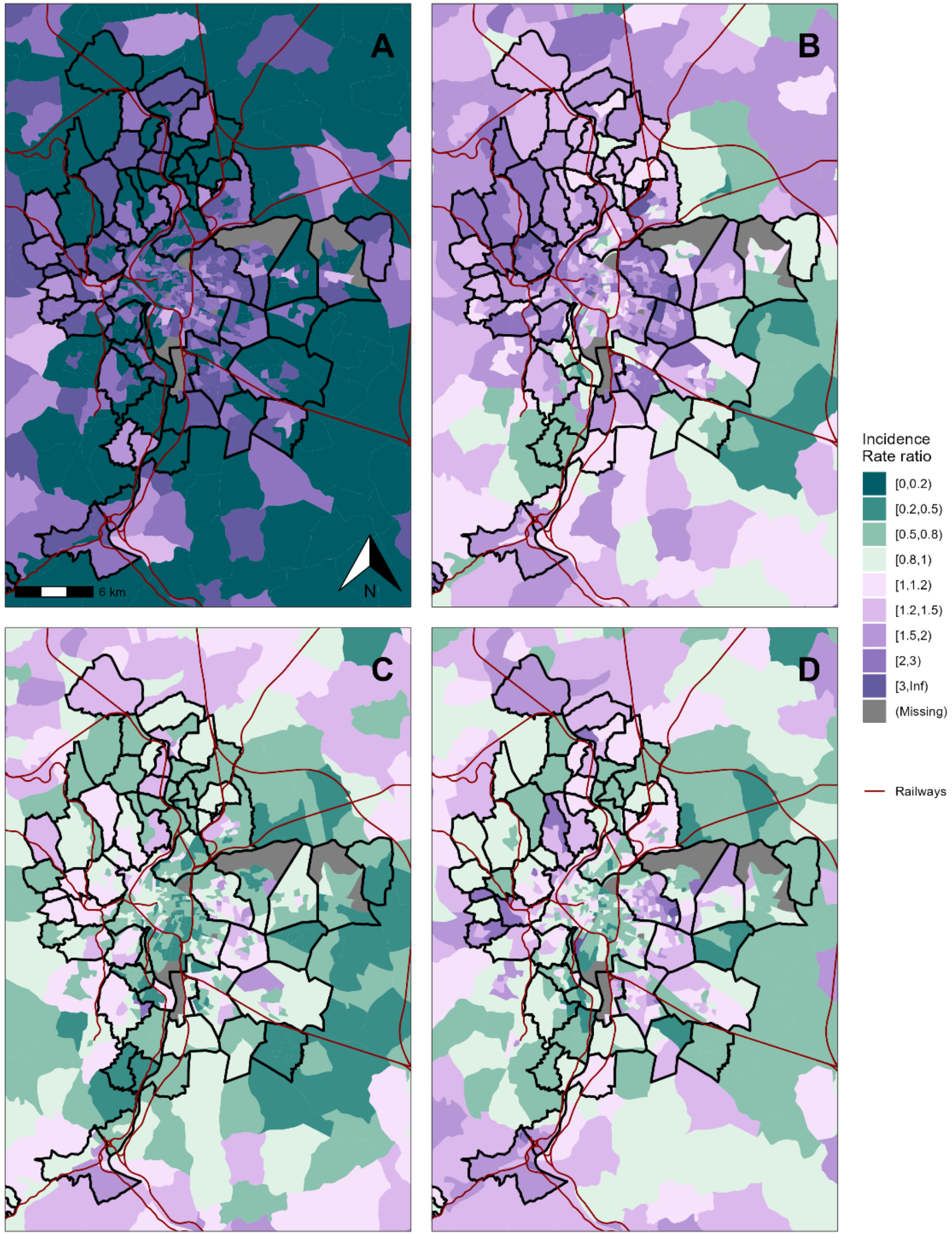


*Figure 4 predicted incidence rate ratio of SARS-CoV-2 infection for the IRIS (infra-municipal spatial unit) of the metropolitan area of Lyon during the second epidemic wave: model with covariates and spatial effect (M4). From A to D: Low incidence, Growth, Peak and decrease, and Stabilization periods*



Back to title

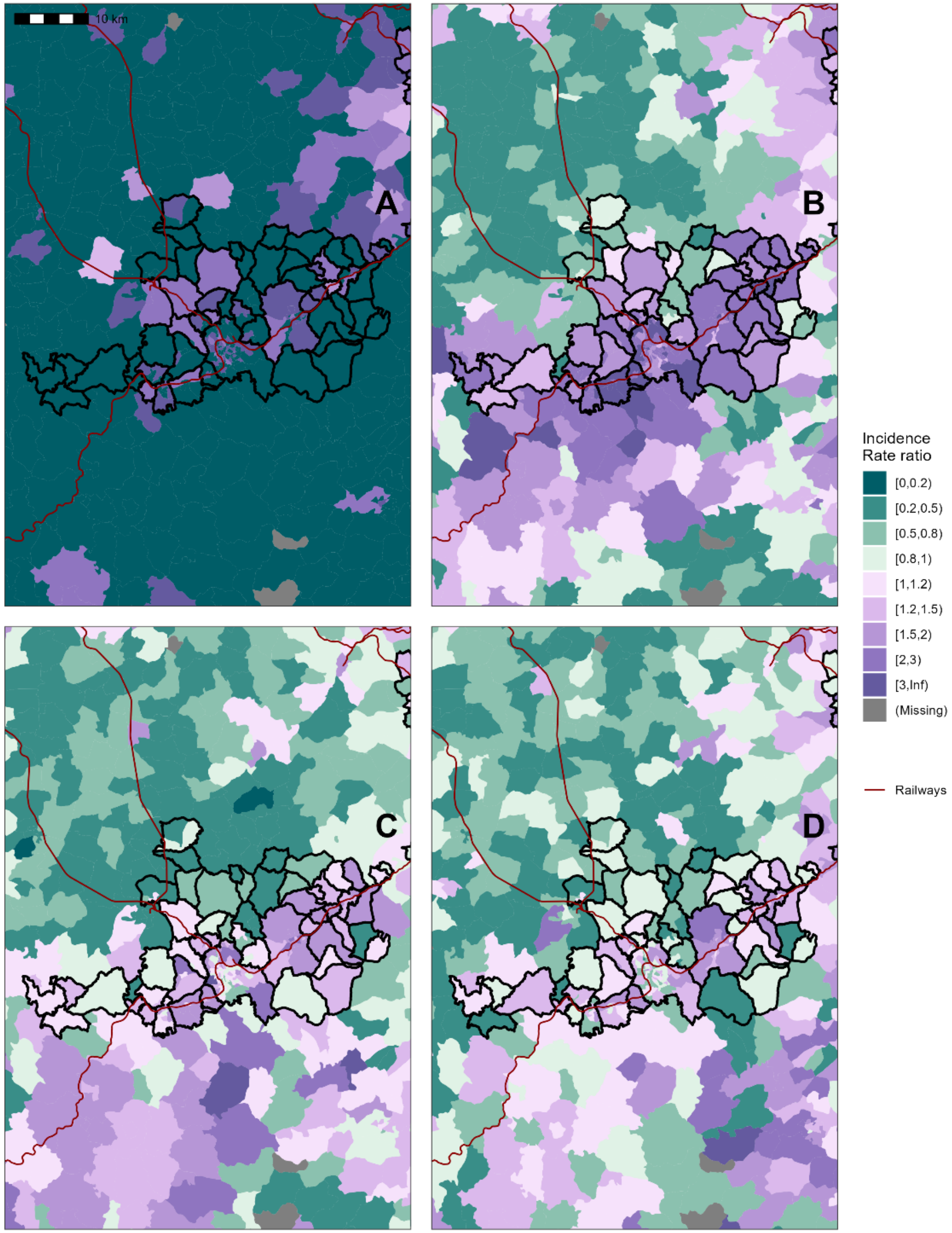


*Figure 5 predicted incidence rate ratio of SARS-CoV-2 infection for the IRIS (infra-municipal spatial unit) of the metropolitan area of Saint-Etienne during the second epidemic wave: model with covariates and spatial effect (M4). From A to D: Low incidence, Growth, Peak and decrease, and Stabilization periods*



[Back to title](#)
