## Supplementary material for "Usefulness of ecological mobility and socio-economic indicators in SARS-CoV-2 infection modelling: a French case study": S5 - Sensitivity analysis results.pdf

The following figures display the estimated relative risks and CAR parameters for the sensitivity analysis. For each subunit of the study region (12 *Départements*), we estimated the parameters of the full model. We evaluate whether the local parameters are congruent with the global estimates, and whether they are congruent amongst themselves. Estimations for the Low incidence period for each subunit are unstable and should not be interpreted as accurate results.



Back to title

Figure 1 Administrative subdivisions of the Auvergne-Rhône-Alpes region. ML denotes the Lyon Metropole. .......... 3
Figure 3 Rate ratios, Low incidence period .......... 4
Figure 4 Rate ratios, Growth period .......... 5
Figure 5 Rate ratios, Peak and decrease period .......... 6
Figure 6 Rate ratios, Stabilization period .......... 7
Figure 7 Spatial effect parameters, Low incidence period .......... 8
Figure 8 Spatial effect parameters, Growth period .......... 9
Figure 9 Spatial effect parameters, Peak and decrease period .......... 10
Figure 10 Spatial effect parameters, Stabilization period .......... 11

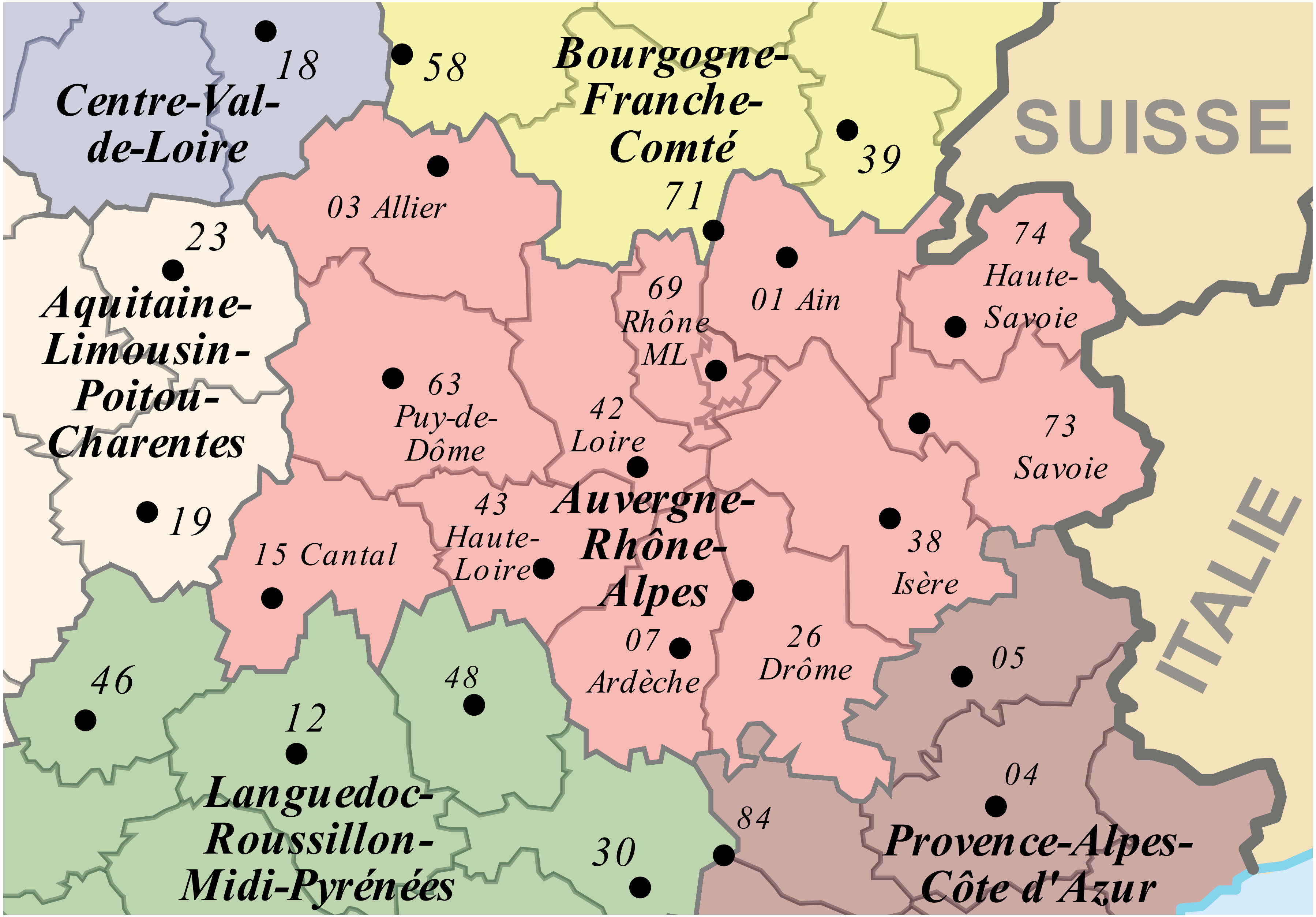


*Figure 1 Administrative subdivisions of the Auvergne-Rhône-Alpes region. ML denotes the Lyon Metropole. Source: https://commons.wikimedia.org/wiki/File:Auvergne-Rh%C3%B4ne-Alpes_with_names_and_numeros_2016.svg, Wikimedia Commons*



[Back to title](#)

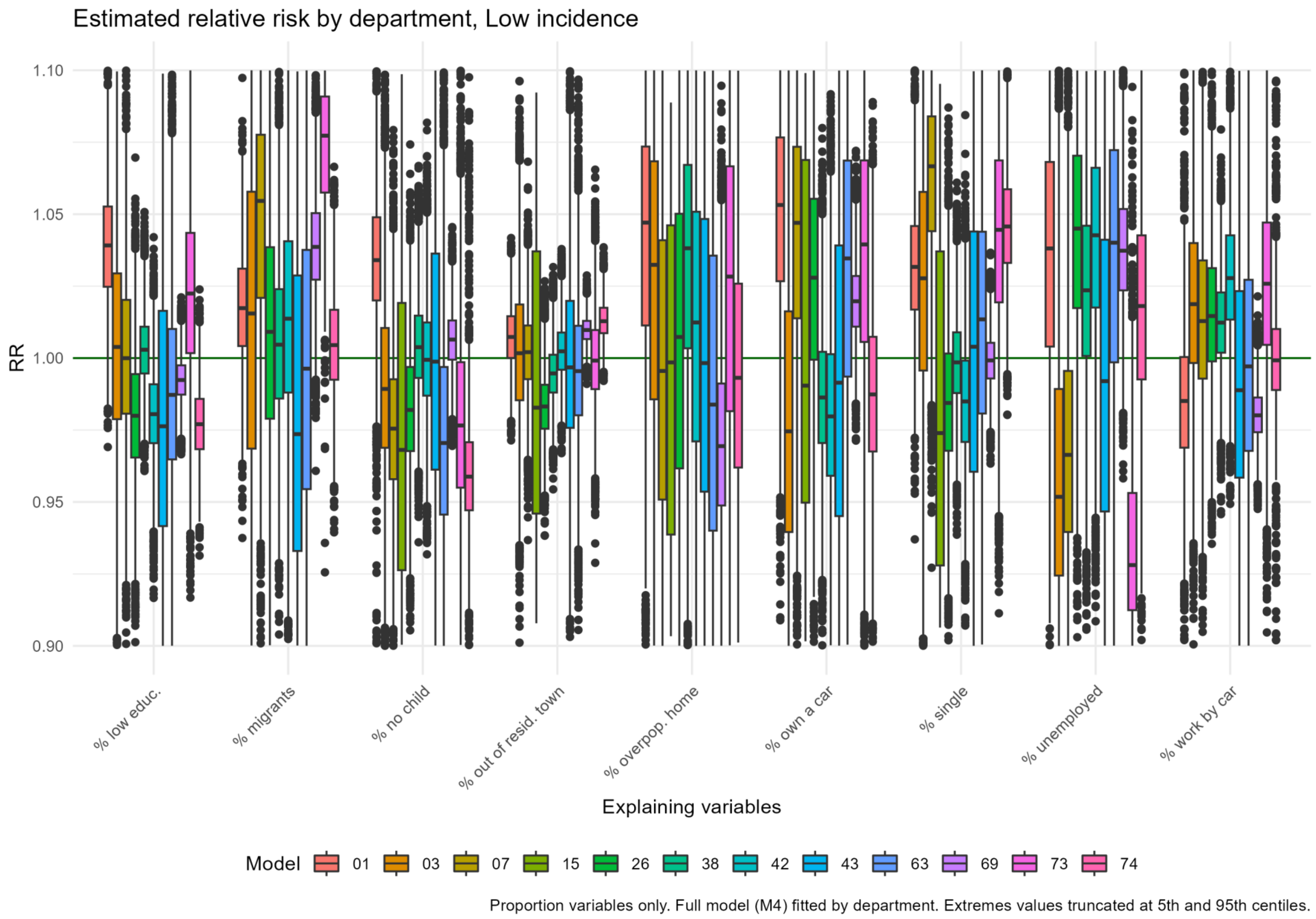


*Figure 2 Posterior distribution of the rate ratio estimates, quantifying the effect of the socio-economic covariates on the SARS-CoV-2 infection rate in the IRIS (infra-municipal spatial units) of the 12 Départements of the French region Auvergne-Rhône-Alpes, during the Low incidence period of the second epidemic wave: results of the model with covariates and spatial effect (M4)*



Back to title

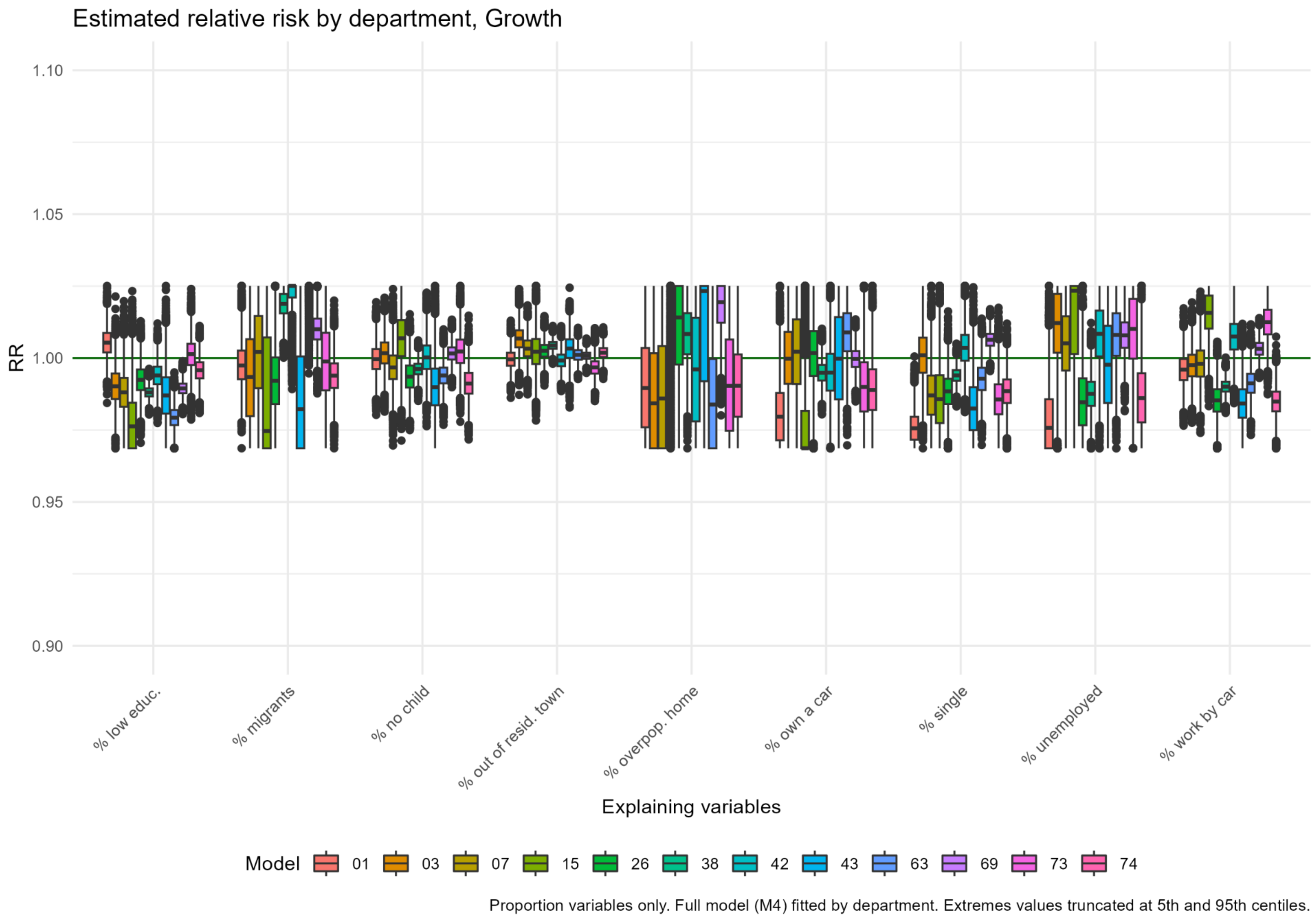


*Figure 3 Posterior distribution of the rate ratio estimates, quantifying the effect of the socio-economic covariates on the SARS-CoV-2 infection rate in the IRIS (infra-municipal spatial units) of the 12 Départements of the French region Auvergne-Rhône-Alpes, during the Growth period of the second epidemic wave: results of the model with covariates and spatial effect (M4)*



Back to title

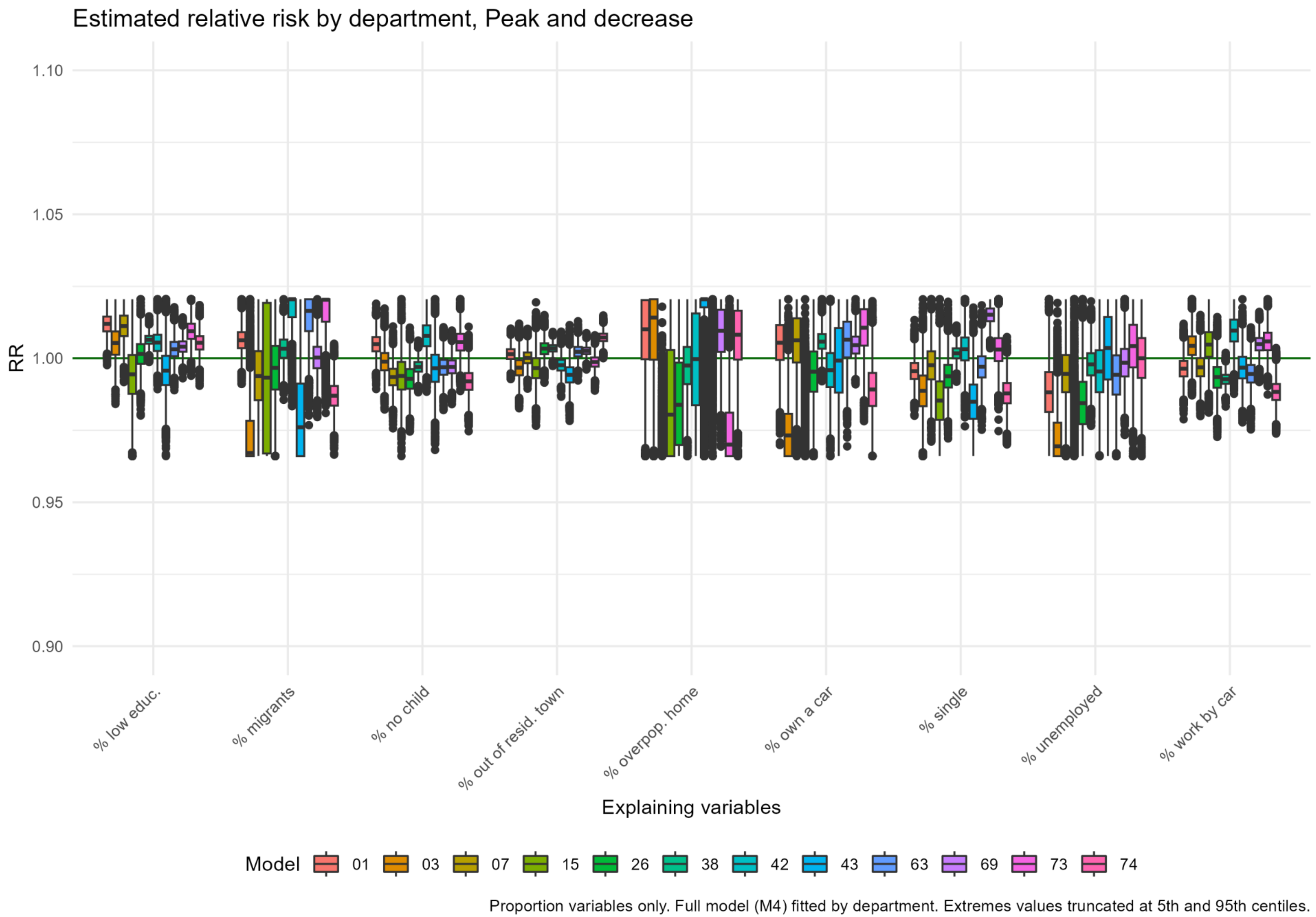


*Figure 4 Posterior distribution of the rate ratio estimates, quantifying the effect of the socio-economic covariates on the SARS-CoV-2 infection rate in the IRIS (infra-municipal spatial units) of the 12 Départements of the French region Auvergne-Rhône-Alpes, during the Peak and decrease period of the second epidemic wave: results of the model with covariates and spatial effect (M4)*



Back to title

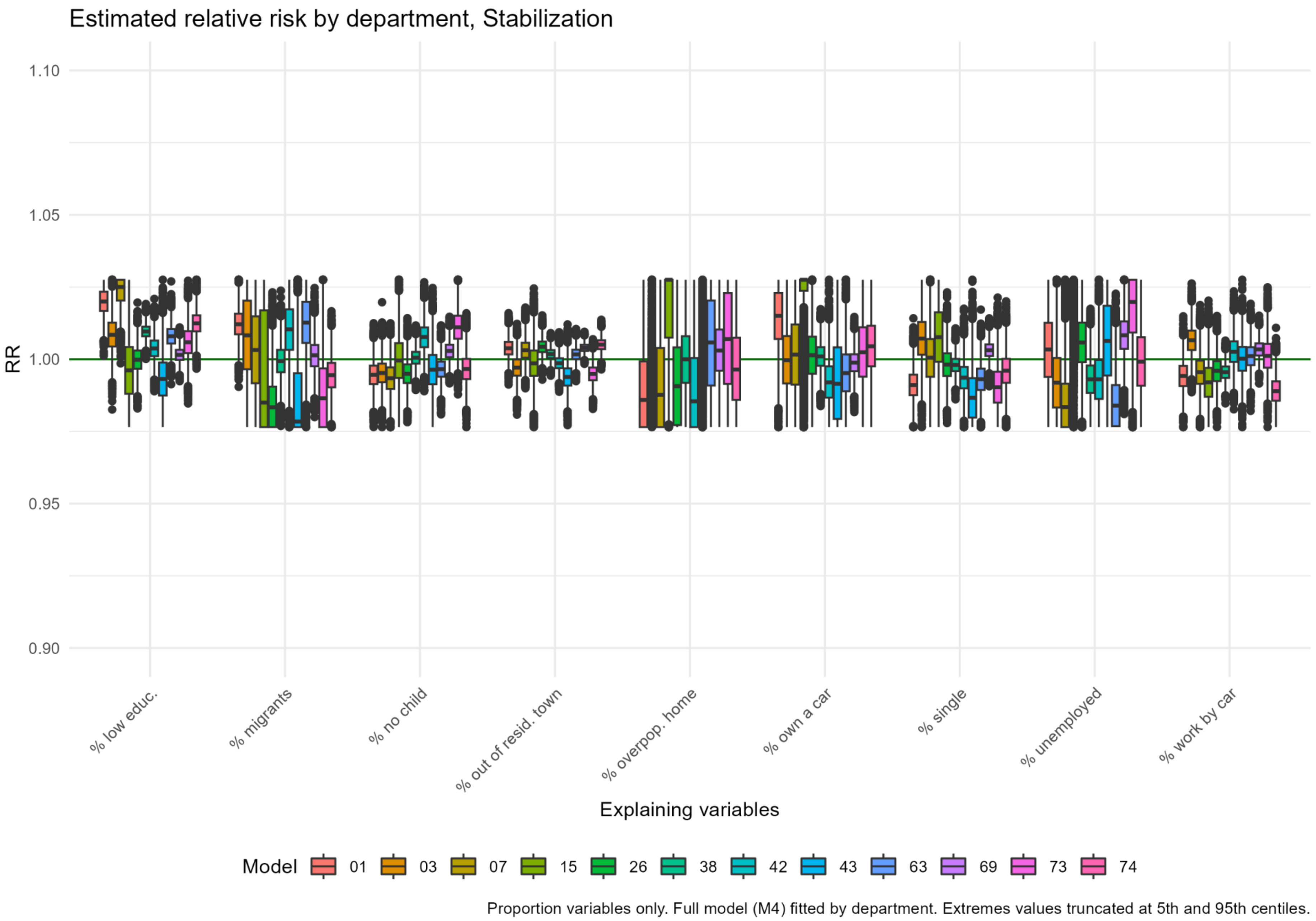


*Figure 5 Posterior distribution of the rate ratio estimates, quantifying the effect of the socio-economic covariates on the SARS-CoV-2 infection rate in the IRIS (infra-municipal spatial units) of the 12 Départements of the French region Auvergne-Rhône-Alpes, during the Stabilization period of the second epidemic wave: results of the model with covariates and spatial effect (M4)*



Back to title

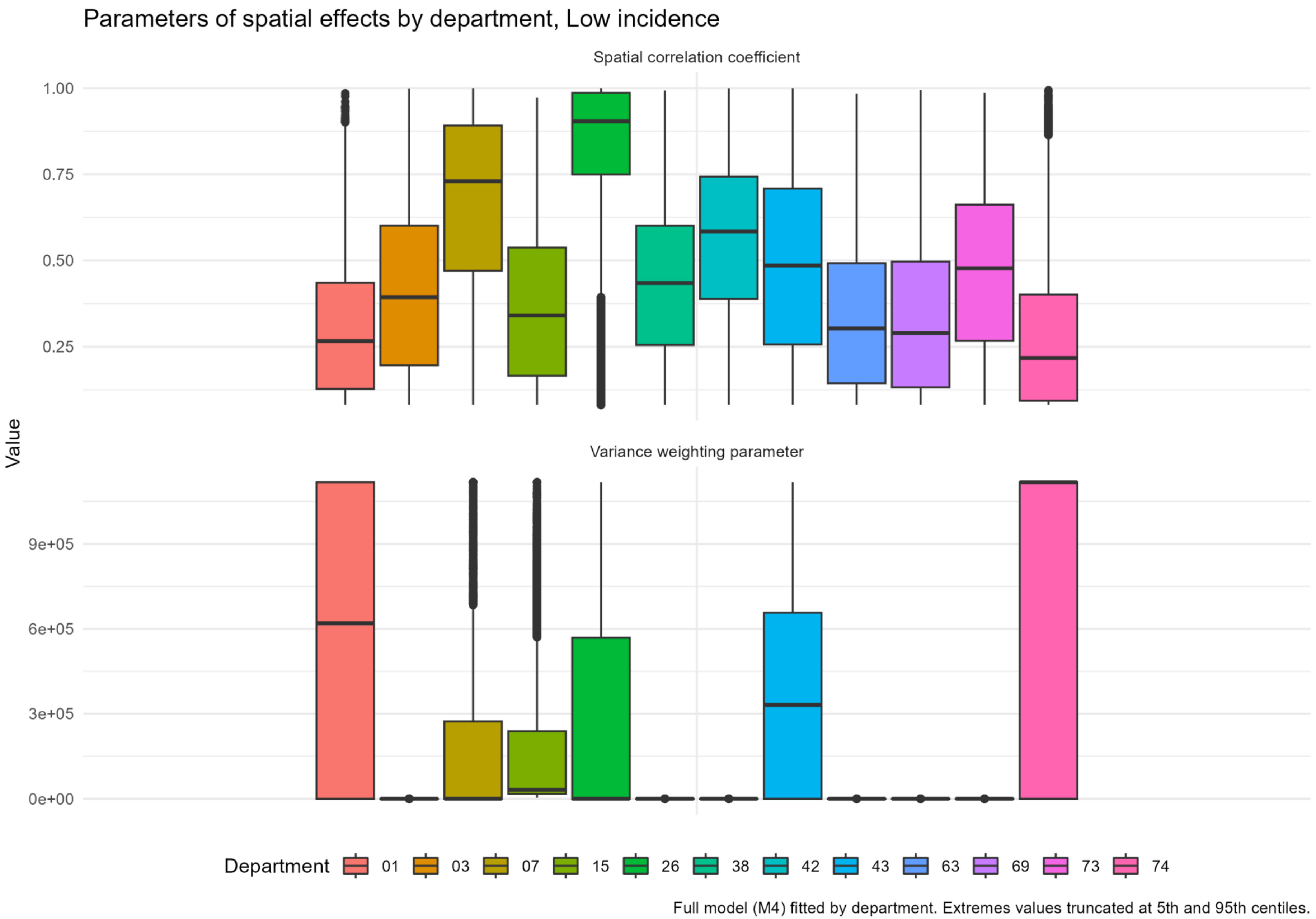


*Figure 6 Posterior distribution of the spatial effect parameters' estimates in the 12 Départements of the French region Auvergne-Rhône-Alpes, during the Low incidence period of the second epidemic wave: results of the model with covariates and spatial effect (M4)*



Back to title

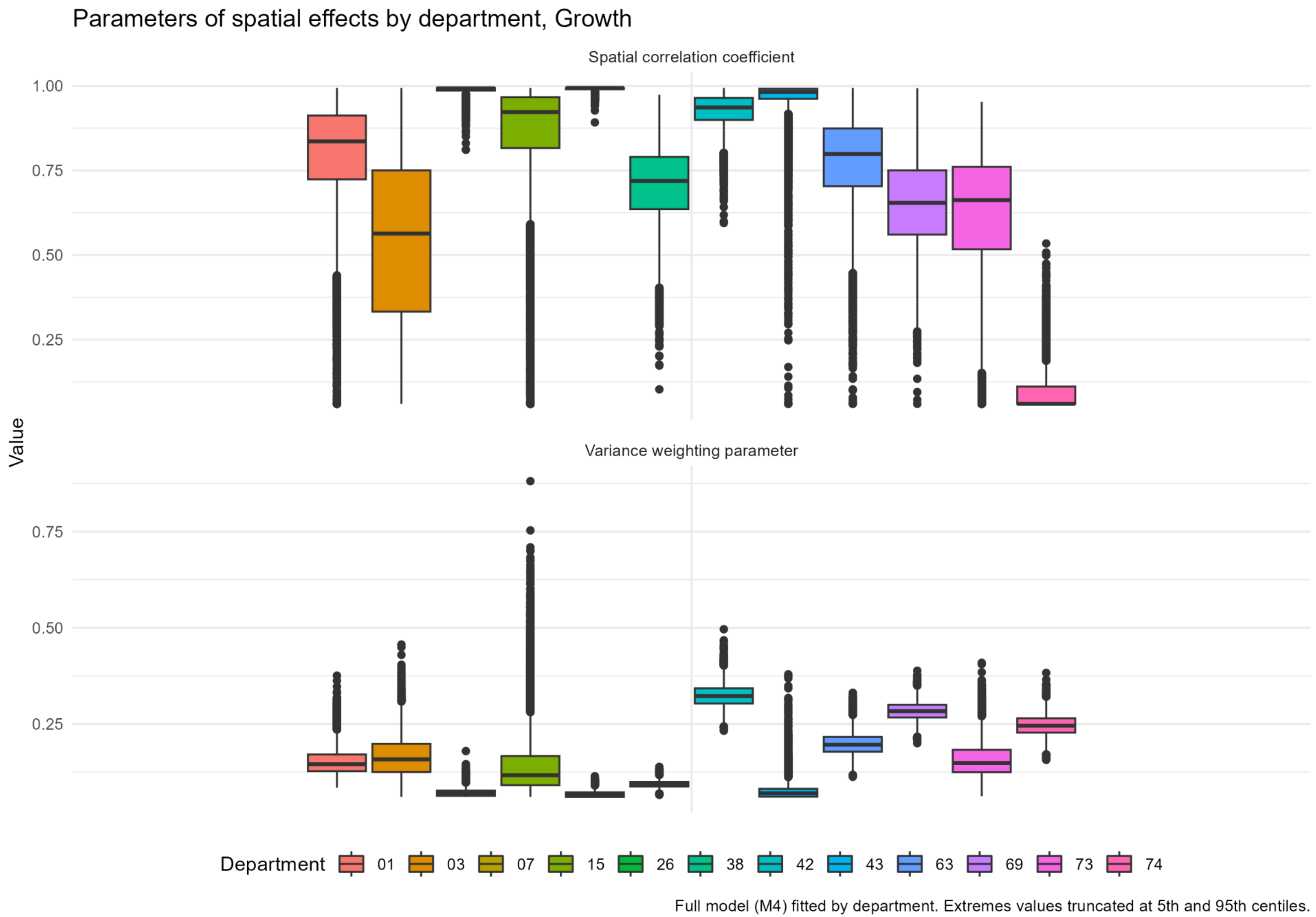


*Figure 7 Posterior distribution of the spatial effect parameters' estimates in the 12 Départements of the French region Auvergne-Rhône-Alpes, during the Growth period of the second epidemic wave: results of the model with covariates and spatial effect (M4)*



Back to title

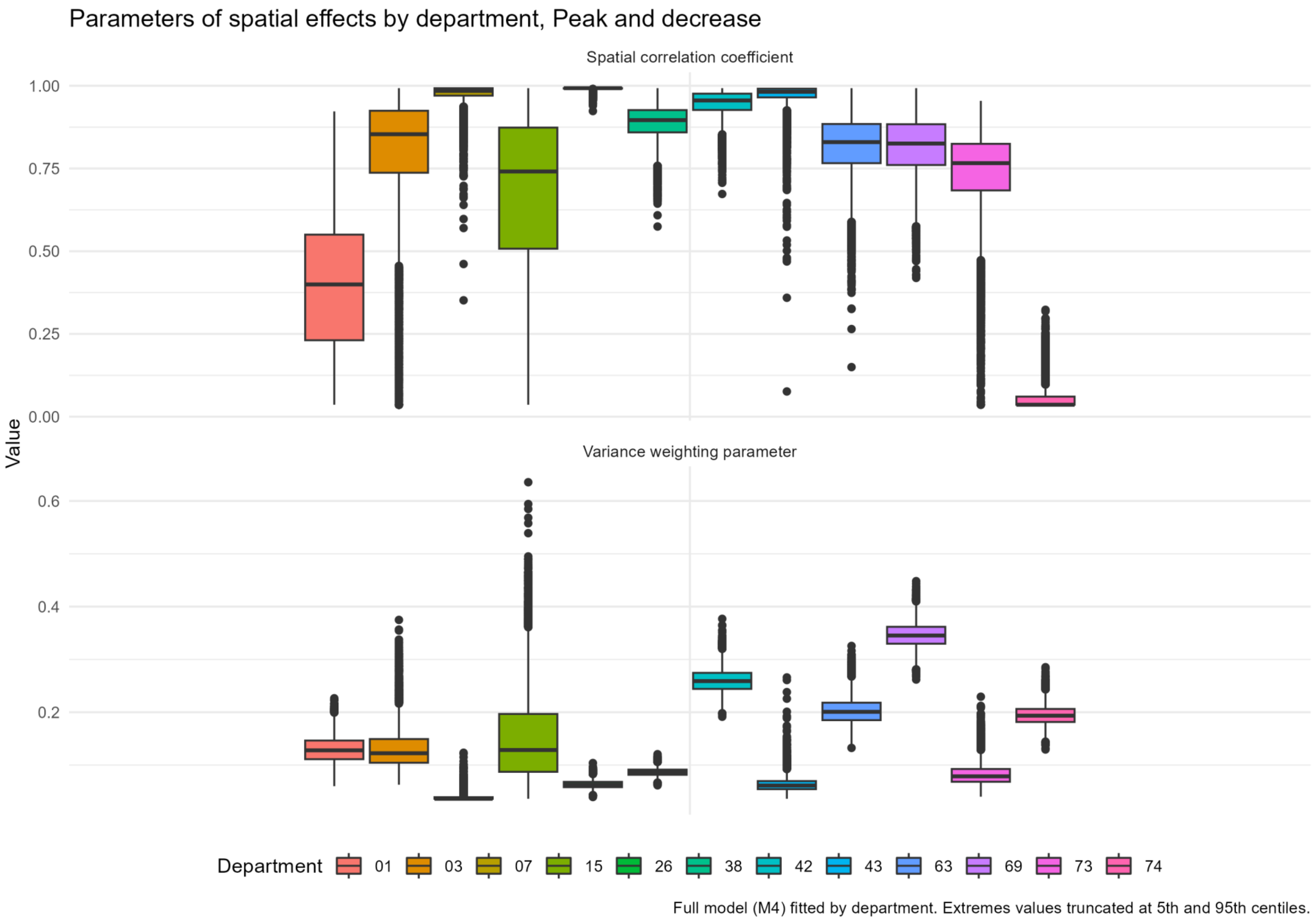


*Figure 8 Posterior distribution of the spatial effect parameters' estimates in the 12 Départements of the French region Auvergne-Rhône-Alpes, during the Peak and decrease period of the second epidemic wave: results of the model with covariates and spatial effect (M4)*



Back to title

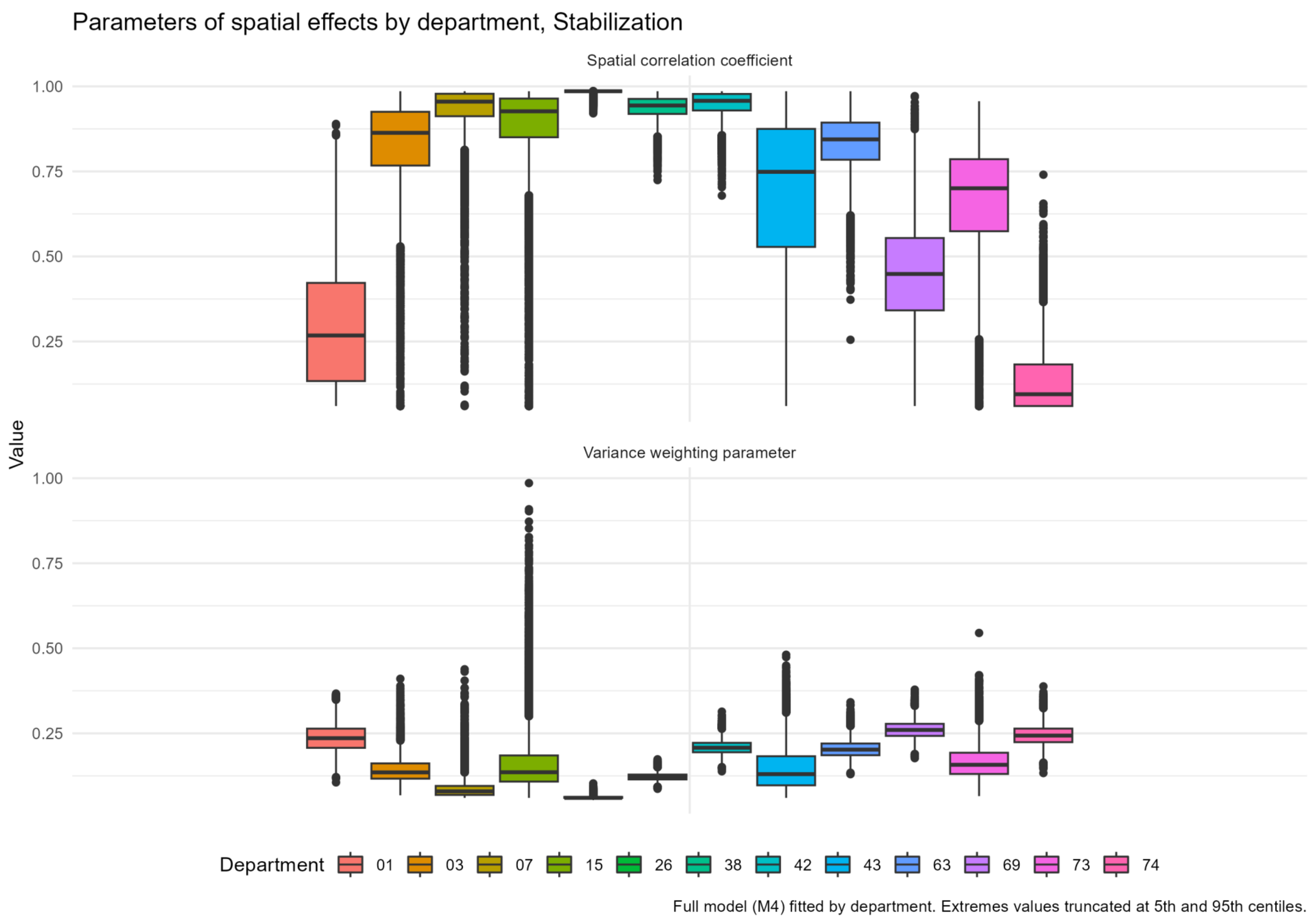


*Figure 9 Posterior distribution of the spatial effect parameters' estimates in the 12 Départements of the French region Auvergne-Rhône-Alpes, during the Stabilization period of the second epidemic wave: results of the model with covariates and spatial effect (M4)*



Back to title
